# Circulating polyunsaturated fatty acids and COVID-19: a prospective cohort study and Mendelian randomization analysis

**DOI:** 10.1101/2022.02.06.22270562

**Authors:** Yitang Sun, Radhika Chatterjee, Akash Ronanki, Kaixiong Ye

## Abstract

**Background:** Higher circulating polyunsaturated fatty acids (PUFAs), especially omega-3 ones, have been linked to a better prognosis in patients of coronavirus disease 2019 (COVID-19). However, the effects and causality of pre-infection PUFA levels remain unclear.

**Objective:** To investigate the observational and causal associations of circulating PUFAs with COVID-19 susceptibility and severity.

**Design:** We first performed a prospective cohort study in UK Biobank, with 20,626 controls who were tested negative and 4,101 COVID-19 patients, including 970 hospitalized ones. Plasma PUFAs at baseline were measured by nuclear magnetic resonance, including total PUFAs, omega-3 PUFAs, omega-6 PUFAs, docosahexaenoic acid (DHA), linoleic acid (LA), and the omega-6/omega-3 ratio. Moreover, bidirectional two-sample Mendelian randomization (MR) analyses were performed to examine the causal associations of eight individual PUFAs, measured in either plasma or red blood cells, with COVID-19 susceptibility and severity using summary statistics from existing genome-wide association studies.

**Results:** In the observational association analysis, total PUFAs, omega-3 PUFAs, omega-6 PUFAs, DHA, and LA were associated with a lower risk of severe COVID-19. Omega-3 PUFAs and DHA were also associated with a lower risk of testing positive for COVID-19. The omega-6/omega-3 ratio was positively associated with risks of both susceptibility and severity. The forward MR analysis indicated that arachidonic acid (AA) and docosapentaenoic acid (DPA-n3) might be causally associated with a lower risk of severe COVID-19, with OR (95% CI) per one SD increase in the plasma level as 0.96 (0.94, 0.99) and 0.89 (0.81, 0.99), respectively. The reverse MR analysis did not support any causal effect of COVID-19 on PUFAs.

**Conclusions:** Our observational analysis supported that higher circulating PUFAs, either omega-3 or omega-6, are protective against severe COVID-19, while omega-3 PUFAs, especially DHA, were also associated with reducing COVID-19 susceptibility. Our MR analysis further supported causal associations of AA and DPA-n3 with a lower risk of severe COVID-19.

## Introduction

The coronavirus disease 2019 (COVID-19) pandemic, caused by the severe acute respiratory syndrome coronavirus 2 (SARS-CoV-2), has resulted in over five million deaths in less than two years (1, 2). Understanding the role of nutrition in moderating susceptibility to and progression of COVID-19 is critical for the development of evidence-based dietary recommendations to prevent infection and to manage disease progression (3, 4). Omega-3 and omega-6 polyunsaturated fatty acids (PUFAs) are of special interest because of their potent immunomodulatory effects, not only in mounting immune responses against viral infection but also in promoting inflammation resolution to avoid tissue damage (5-7). COVID-19 is an infectious disease characterized by cytokine storm and hyperinflammation in severe cases (8), presenting multiple possible points of action for PUFAs.

Recent observational studies have noted significant changes in the circulating levels of various PUFAs when comparing COVID-19 patients to healthy controls and across severity subgroups of patients. In general, total PUFAs, omega-6 PUFAs, linoleic acid (LA), and the omega-3 index measured as the percentage of eicosapentaenoic acid (EPA) and docosahexaenoic acid (DHA) in red blood cell (RBC) fatty acids, are lower in COVID-19 patients and even lower in severe cases (9-12). A higher omega-3 index in patients was further associated with lower risks of requiring mechanical ventilation and death (9, 10). But conflicting patterns were also reported across cohorts and studies (11, 12), such as elevated levels of LA and arachidonic acid (AA) in COVID-19 patients (12). Moreover, the circulating levels of PUFAs in patients are likely confounded by immune responses to the viral infection and do not represent the effects of pre-infection circulating levels. There is a prospective cohort study that compared hospitalized COVID-19 patients to non-cases and found that almost all PUFA measurements, including total PUFAs, omega-6 PUFAs, omega-3 PUFAs, LA, and DHA, are associated with a lower risk of severe COVID-19. The only exception is the omega-6/omega-3 ratio, which exhibits a positive association (13). However, the study did not distinguish the effects on susceptibility and severity, and the usage of non-cases without COVID-19 status as the control did not correct for selection bias in those receiving tests. Altogether, while these observational studies provide valuable insights, they are susceptible to residual confounding and reverse causation. The causal effects of circulating PUFAs on COVID-19 susceptibility and severity remain unclear.

Mendelian randomization (MR) is an analytic tool for inferring the causal effects of an exposure on an outcome of interest (14). MR uses randomly allocated genetic variants related to the exposure as instrumental variables, which are inborn and minimally affected by confounders and reverse causation (15). This method has been widely utilized in recent studies to evaluate the causal roles of specific risk factors in COVID-19, such as body mass index (BMI), white blood cells, some circulating proteins, and smoking (16-19). On the other hand, MR studies have also provided support for the causal clinical effects of circulating PUFAs (Supplemental Table 1). The genetically predicted circulating levels of various PUFAs have been associated with clinical biomarkers, such as blood lipids, white blood cell counts, and blood pressure (20-22). They were also directly associated with risks of specific diseases, such as cardiovascular diseases, diabetes, and cancers (23-27). Therefore, MR is a valuable and cost-effective tool to evaluate the causal roles of circulating PUFAs in COVID-19 susceptibility and severity.

In this study, we first performed an observational analysis in a prospective cohort, UK Biobank, with 4,101 COVID-19 patients, including 970 hospitalized ones, and 20,626 controls that were tested negative. We performed multiple comparisons across different case and control groups to evaluate the effects of six baseline plasma PUFA measures on COVID-19 susceptibility and severity. Furthermore, we applied bidirectional two-sample MR analyses to examine the causal associations between eight individual PUFAs and COVID-19. Genetic instruments for circulating PUFAs were obtained from previous genome-wide association studies (GWAS) of corresponding PUFAs measured in either plasma or RBC (28-30). Genetic associations with COVID-19 susceptibility and severity were obtained from GWAS meta-analyses conducted by the COVID-19 Host Genetics Initiative (HGI) (31). Our study, integrating observational and genetics-instrumented MR analyses, unraveled the effects of total and individual circulating PUFAs on the risks of COVID-19 susceptibility and severity.

## Methods

### Ethical considerations

The usage of individual-level data for this study was approved by the University of Georgia Institutional Review Board and UK Biobank (application no. 48818). All participants of UK Biobank and the Framingham Heart Study (FHS) provided written informed consent before joining these studies. Informed consent was not required for publicly available summary statistics. Our study follows the guidelines for strengthening the reporting of observational studies in epidemiology (STROBE, Supplemental Table 2) and strengthening the reporting of Mendelian randomization studies (STROBE-MR, Supplemental Table 3) (32).

### Participants and study design

We performed an observational cohort study based on UK Biobank and then a bidirectional two-sample MR study with summary statistics from GWAS of PUFAs and COVID-19. UK Biobank is a population-based prospective study, including >500,000 participants aged 37–73 years at recruitment from 2006 to 2010 in the United Kingdom (33). The observational analysis was performed to examine the associations between six plasma PUFA measures and COVID-19 status in UK Biobank. The six plasma PUFA measures include total PUFAs, omega-3 PUFAs, omega-6 PUFAs, DHA, LA, and the calculated omega-6/omega-3 ratio. The MR study investigated the causal effects of eight individual PUFAs on COVID-19 susceptibility and severity. Genetic instruments for plasma PUFAs were obtained directly from published GWAS (28, 29). Genetic instruments for RBC PUFAs were determined based on a published GWAS, but their summary statistics, not reported in the original study, were calculated by ourselves with the same statistical model and individual-level data from 2,462 FHS participants (30). Six PUFAs have genetic instruments for their circulating levels in both plasma and RBC, including α-Linolenic acid (ALA), docosapentaenoic acid (DPA-n3), LA, γ-Linoleic acid (GLA), dihomo-γ-linoleic acid (DGLA), and AA. Docosatetraenoic acid (DTA) only has genetic instruments for its RBC level, while DHA only for its plasma level.

### Observational analysis

Figure 1 displays the flow of participants throughout the observational study. To minimize the possibility of bias due to population stratification, the analysis was restricted to individuals of European descent. In addition, we removed participants who had mismatched self-reported sex and genetic sex, sex chromosome aneuploidy, ten or more third-degree or closer relatives, or had withdrawn from UK Biobank. Our exposure variables were six PUFAs, as measured by nuclear magnetic resonance (NMR) in plasma samples collected between 2007 and 2010 (13, 33, 34). We used the COVID-19 testing result and inpatient status as our outcome (data accessed on June 21, 2021). The specimen collection dates were March 16, 2020 to June 14, 2021 for those in England; February 11, 2020 to March 18, 2021 in Scotland; and January 13, 2020 to June 7, 2021 in Wales. Hospitalized COVID-19 patients were identified as those with positive PCR-based diagnosis and explicit evidence of being inpatients. Of note, being an inpatient does not necessarily indicate hospitalization for COVID-19 because patients in hospitals for any reason may be prioritized for COVID-19 testing (35). Inpatient status was not available for assessment centers in Scotland and Wales. To test the association with COVID-19 severity, we performed two separate analyses with different controls: 1) non-hospitalized COVID-19 patients, and 2) individuals who tested negative. To examine the association with COVID-19 susceptibility, we focused on all COVID-19 cases which were tested positive for SARS-CoV-2. Individuals with negative tests were used as the control. This analysis of susceptibility was performed in two datasets: 1) participants from England, and 2) participants from England, Scotland, and Wales. For the 24,727 participants with both plasma PUFA measures and COVID-19 status, we applied logistic regression models on various case and control groups to estimate the associations of PUFAs with COVID-19 susceptibility and severity. Covariates included continuous variables, age, BMI, and Townsend deprivation index, and categorical variables, sex, ethnicity, and assessment center. Individuals with missing information in PUFA measures, COVID-19 status, or covariates were excluded. The comparable effect sizes were expressed per one standard deviation (SD) increase in the plasma PUFAs. All analyses in the observational study were conducted using R version 4.0.0, and nominal significance was set at *p*-value < 0.05. Bonferroni correction for multiple testing [corrected *P* significance cutoff: 0.05/2 (outcomes)/6 (exposures) = 0.0042] was used to avoid the type I error (36).

**Figure 1.**
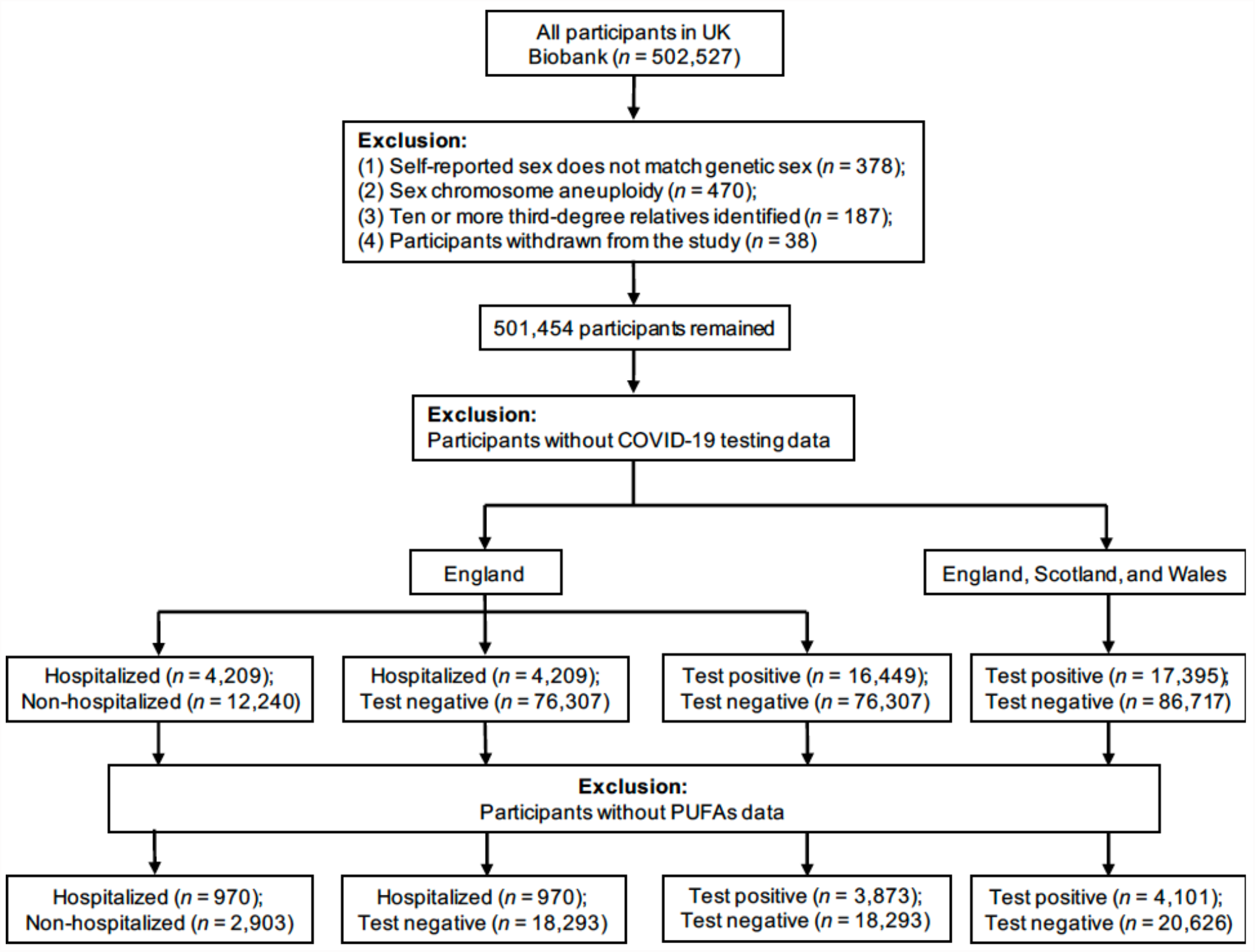
Flowchart of the UK Biobank participants from recruitment to inclusion in the observational analysis.

### Genetic associations with PUFAs

Two types of circulating PUFAs were evaluated in our MR analyses, plasma and RBC PUFAs. For plasma PUFAs, single nucleotide polymorphisms (SNPs) were obtained from published GWAS of omega-3 PUFAs (n = 8,866) and omega-6 PUFAs (n = 8,631) in participants of European ancestry (28, 29). We selected SNPs for each plasma omega-3 and omega-6 PUFA, which reached genome-wide significance level (*P* < 5 × 10^−8^) and were restricted by linkage disequilibrium (LD) clumping to ensure independence (R^2^ < 0.001 within a 10 Mb window). To ensure robustness and reduce false positives, we also used less stringent LD cutoffs (R^2^ < 0.01, 0.1, and 0.3) to select SNPs associated with plasma omega-3 PUFAs. The same LD-related sensitivity analysis was not possible for plasma omega-6 PUFAs because their genome-wide summary statistics were not available. To examine the effects of RBC PUFAs, we obtained genetic associations at a genome-wide significance level (*P* < 5 × 10^−8^) identified by Tintle *et al*. (30). We used the individual-level data from the FHS to confirm the significance of these SNPs and calculate their effect sizes and standard errors. In the same linear mixed model, covariates included age, sex, and matrix of kinship coefficients in the FHS. We respectively selected independent (R^2^ < 0.001, 0.01, 0.1, and 0.3 within a 10 Mb window) SNPs predicting RBC PUFAs at genome-wide significance (*P* < 5 × 10^−8^). We calculated *F*-statistics to test instrument strength (*F*-statistics >10 for all plasma and RBC PUFAs) (37). Summary statistics for the genetic instruments for plasma and RBC PUFAs are openly available for public access (Supplemental Tables 4 and 5).

### Genetic associations with COVID-19

To assess genetic associations with COVID-19 severity, we used three GWAS meta-analyses conducted by the HGI (release 5, released on January 18, 2021) (31). First, we used the GWAS of severe COVID-19, labeled as study A2, that compared patients confirmed with very severe respiratory symptoms (n = 5,101) to the control group of general population samples (n = 1,383,241). Second, another HGI GWAS, labeled as study B2, compared hospitalized COVID-19 patients (n = 9,986) to general population samples (n = 1,877,672). The third severe COVID-19 GWAS utilized in our study, labeled as B1, compared hospitalized COVID-19 patients (n = 4,829) to non-hospitalized COVID-19 patients (n = 11,816). To assess genetic associations with COVID-19 susceptibility, we used one GWAS by HGI, labeled as study C2, that compared any COVID-19 case (n = 38,984) to population controls (n = 1,644,784). In addition to these four COVID-19 GWAS used in our primary analysis, we repeated MR analyses using the study A2, B1, B2, and C2 from HGI release 4 (released on October 20, 2020), to examine the consistency of our findings across different data releases. Detailed information about these GWAS is available at the COVID-19 HGI website (https://www.covid19hg.org/results/).

To assess reverse causality, we obtained strong (*P* < 5 × 10^−8^) and independent (R^2^ < 0.001 within a 10 Mb clumping window) SNPs associated with COVID-19 phenotypes as genetic instruments. We also used a less stringent selection criterion (*P* < 5 × 10^−6^) to determine the robustness of our results.

### MR analyses

MR was used to infer causality between PUFAs and COVID-19 by leveraging genetic data as instrumental variables. We scaled the odds ratio (OR) estimates per SD increment of plasma and RBC PUFAs (% of total fatty acids). We obtained the SNP-specific Wald estimate (ratio of the SNP-outcome effect divided by the SNP-exposure effect) when only one SNP was available. The inverse variance-weighted (IVW) method with a multiplicative random-effects model (≥2 SNPs) was used as the primary analysis (38-40). We used the MR-Egger intercept test to evaluate the extent of unbalanced horizontal pleiotropy, which can lead to a biased causal effect estimate (39). In sensitivity analyses, we applied the MR-Egger and weighted median (WM) methods to account for pleiotropy (39-41). The MR-Egger method provides an unbiased causal estimate even when all SNPs are invalid instruments as long as that the horizontal pleiotropic effects are balanced across SNPs (39). However, MR-Egger can be imprecise and suffer from low statistical power, particularly when based on a small number of SNPs (e.g., < 10) (39). The WM method gives robust causal estimates even when up to 50% of SNPs are invalid genetic instruments (41). To test the presence of heterogeneity among genetic instruments, we calculated Cochran’s Q statistic for the IVW method and an extended version of Cochran’s Q statistic (Rücker’s Q′) for the MR-Egger method (42, 43). We utilized Bonferroni correction [corrected *P* significance cutoff: 0.05/2 (outcomes)/7 (exposures) = 0.0036] for multiple testing. Additionally, we required a relationship to be nominally significant (*P* < 0.05) with both measures of the same PUFA (plasma and RBC) and in the case of COVID-19 severity, with different outcome GWAS (study A2, B2, and B1). All MR analyses were performed in R version 4.0.0 with the TwoSampleMR package version 3.6.9 (44).

## Results

### Baseline characteristics

The flow of UK Biobank participants throughout the observational study is described in Figure 1, while their baseline characteristics are summarized in Table 1. Across all assessment centers in England, Scotland, and Wales, there were 104,112 participants with COVID-19 status. Among them, 17,395 were tested positive for COVID-19. Inpatient status was only reported by assessment centers in England. Of the 92,756 participants with COVID-19 status in England, 16,449 were tested positive, and 4,209 had confirmed inpatient status. Across England, Scotland, and Wales, COVID-19 patients were more likely to be male (t-test, *P* = 0.008), with higher BMI (*P* = 9.34 × 10^−14^), but younger than participants with negative testing results (*P* < 2.2 × 10^−16^). Across assessment centers in England, hospitalized COVID-19 patients were older (*P* < 2.2 × 10^−16^), were more likely to be male (*P* = 2.44 × 10^−5^), and had higher BMI (*P* = 1.13 × 10^−14^), when compared to non-hospitalized COVID-19 patients.

**TABLE 1.**
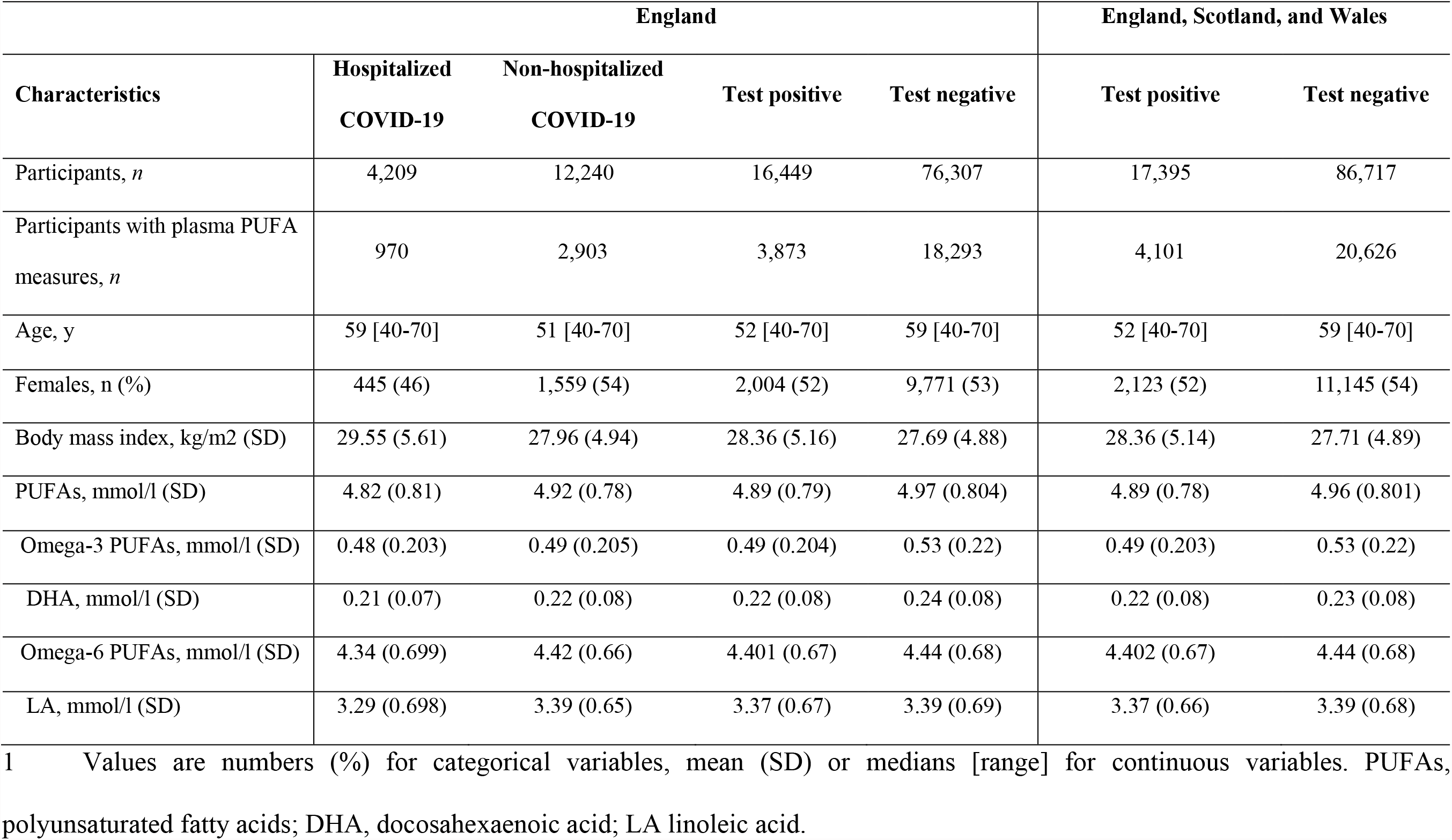
Characteristics of the UK Biobank participants at baseline^1^

### Observational association analysis

Table 2 shows the observational associations between baseline plasma PUFAs and COVID-19 susceptibility and severity. Among participants from England who also had plasma data, there were 18,293 with negative testing results and 3,873 with positive tests. Among the COVID-19 patients, 970 were hospitalized and the other 2,903 were non-hospitalized. Comparing hospitalized patients to those tested negative, we observed a lower risk of COVID-19 severity per SD increase in total PUFAs (OR: 0.88; 95% confidence interval (CI): 0.82, 0.95; *P* = 0.0005), omega-3 PUFAs (OR: 0.82; 95% CI: 0.76, 0.89; *P* = 8.1 × 10^−7^), omega-6 PUFAs (OR: 0.91; 95% CI: 0.85, 0.98; *P* = 0.0121), DHA (OR: 0.78; 95% CI: 0.72, 0.85; *P* = 4.6 × 10^−9^), and LA (OR: 0.92; 95% CI: 0.86, 0.99; *P* = 0.0228). Using 2,903 non-hospitalized COVID-19 patients as the control group, there were consistently inverse associations of COVID-19 severity with total PUFAs (*P* = 0.0012), omega-3 PUFAs (*P* = 0.0013), omega-6 PUFAs (*P* = 0.0047), DHA (*P* = 8.9 × 10^−5^), and LA (*P* = 0.0079).

**TABLE 2.**
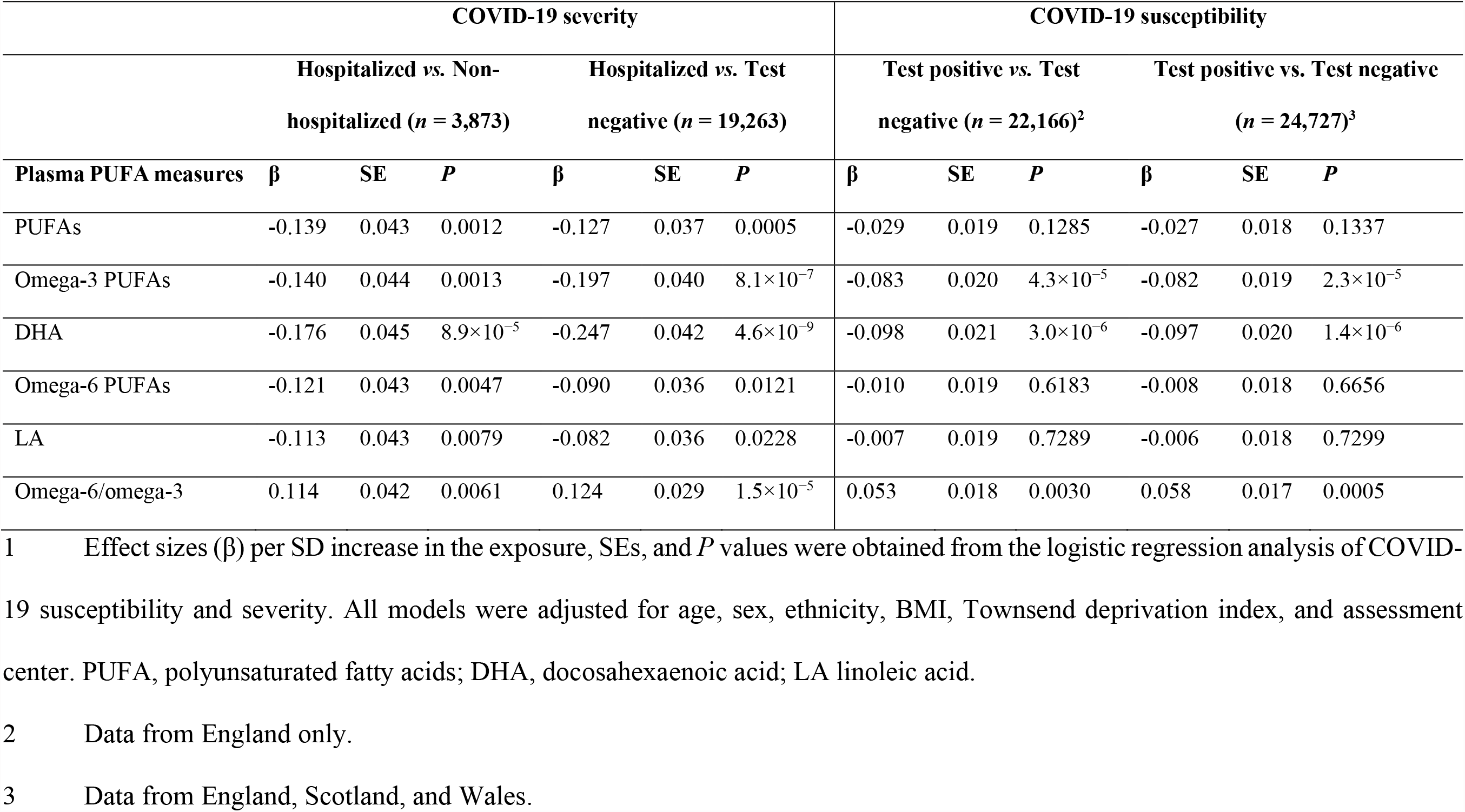
Associations of PUFAs concentrations with COVID-19 susceptibility and severity^1^

We further evaluated the effects of baseline plasma PUFAs on COVID-19 susceptibility by comparing COVID-19 patients to those tested negative. Among 24,727 participants in England, Scotland, and Wales, we found a lower risk of getting COVID-19 per SD increase in omega-3 PUFAs (OR: 0.92; 95% CI: 0.89, 0.96; *P* = 2.3 × 10^−5^) and DHA (OR: 0.91; 95% CI: 0.87, 0.94; *P* = 1.4 × 10^−6^). Among 22,166 individuals in England only, we also observed consistently significant associations for omega-3 PUFAs (OR: 0.92; 95% CI: 0.88, 0.96; *P* = 4.3 × 10^−5^) and DHA (OR: 0.91; 95% CI: 0.87, 0.94; *P* = 3.0 × 10^−6^).

The omega-6/omega-3 ratio was significantly associated with an increased risk of severe COVID-19, either by comparing hospitalized patients to participants who tested negative (OR: 1.13; 95% CI: 1.07, 1.20; *P* = 1.5 × 10^−5^) or to non-hospitalized patients (OR: 1.12; 95% CI: 1.03, 1.22; *P* = 0.0061). The ratio was also positively associated with COVID-19 susceptibility when comparing COVID-19 patients to those tested negative in England, Scotland, and Wales (OR: 1.06; 95% CI: 1.03, 1.10; *P* = 0.0005) or in England only (OR: 1.05; 95% CI: 1.02, 1.09; *P* = 0.0030). Overall, our observational analysis showed that individuals with lower baseline levels of all five examined PUFAs were associated with a higher risk of hospitalized COVID-19, and those with lower levels of omega-3 PUFAs and DHA were also at a higher risk of COVID-19 susceptibility. On the other hand, the omega-6/omega-3 ratio was positively associated with the risks of both COVID-19 susceptibility and severity.

### Bidirectional MR analyses

We performed bidirectional MR analyses to examine the causal relationships between individual PUFAs and COVID-19. First, we performed a forward MR analysis to investigate the effects of PUFAs on COVID-19 susceptibility and severity. Second, we conducted a reverse MR analysis to evaluate the causal effects of genetically instrumented COVID-19 on PUFAs. All genetic instruments for PUFAs (*F*-statistics >31.43) and COVID-19 (*F*-statistics >30.81) were strong instruments. Six individual PUFAs have existing GWAS for their levels in plasma and RBC, and there are three GWAS on severe COVID-19 (i.e., HGI study A2, B2, B1). Only results that were consistent across these different GWAS were reported here.

In the forward MR study of plasma PUFAs, genetically instrumented one-SD increase in AA (OR: 0.96; 95% CI: 0.94, 0.99; *P* = 0.007) and DPA-n3 (OR: 0.89; 95% CI: 0.81, 0.99; *P* = 0.026) were associated with a lower risk of very severe respiratory symptoms of COVID-19 based on HGI study A2 (Figure 2A). Consistently, genetically instrumented AA (OR: 0.96; 95% CI: 0.96, 0.97; *P* = 3.23 × 10^−20^) and DPA-n3 (OR: 0.93; 95% CI: 0.92, 0.95; *P* = 4.73 × 10^−20^) were associated with a lower risk of hospitalized COVID-19 based on HGI study B2, which used general population samples as the control (Figure 2B). Similar results were observed with HGI study B1, which used non-hospitalized COVID-19 patients as the control (Figure 2C). Besides plasma PUFAs, MR analyses with RBC PUFAs consistently support the protective effects of AA against severe COVID-19 based on HGI A2 (OR: 0.97; 95% CI: 0.94, 1.00; *P* = 0.048), B2 (OR: 0.95; 95% CI: 0.93, 0.97; *P* = 1.32 × 10^−5^), and B1 (OR: 0.84; 95% CI: 0.83, 0.85; *P* = 8.57 × 10^−130^) studies (Figures 2). For DPA-n3, its genetically instrumented RBC level was consistently associated with a lower risk of COVID-19 severity in our forward MR analysis with study A2 (OR: 0.79; 95% CI: 0.63, 0.99; *P* = 0.041), B2 (OR: 0.88; 95% CI: 0.82, 0.94; *P* = 9.30 × 10^−5^), and B1 (OR: 0.76; 95% CI: 0.59, 0.98; *P* = 0.036) (Figures 2). To ensure the robustness of findings, we selected genetic instruments based on various LD categories (R^2^ < 0.001, 0.01, 0.1, and 0.3). The causal estimates of AA and DPA-n3 were consistent and at least nominally significant throughout all MR analyses (Supplemental Tables 6–8). Causal estimates for AA and DPA-n3 maintained the same effect directions in MR-Egger and WM methods, and sensitivity tests identified no evidence of horizontal pleiotropy or heterogeneity of effects (Supplemental Tables 6–8). Of note, while there were nominally significant associations between plasma DHA and very severe COVID-19 with HGI A2 and between RBC DTA and hospitalized COVID-19 with HGI B1, these two relationships were not replicated in analyses with the other two GWAS of severe COVID-19 (Figure 2).

**Figure 2.**
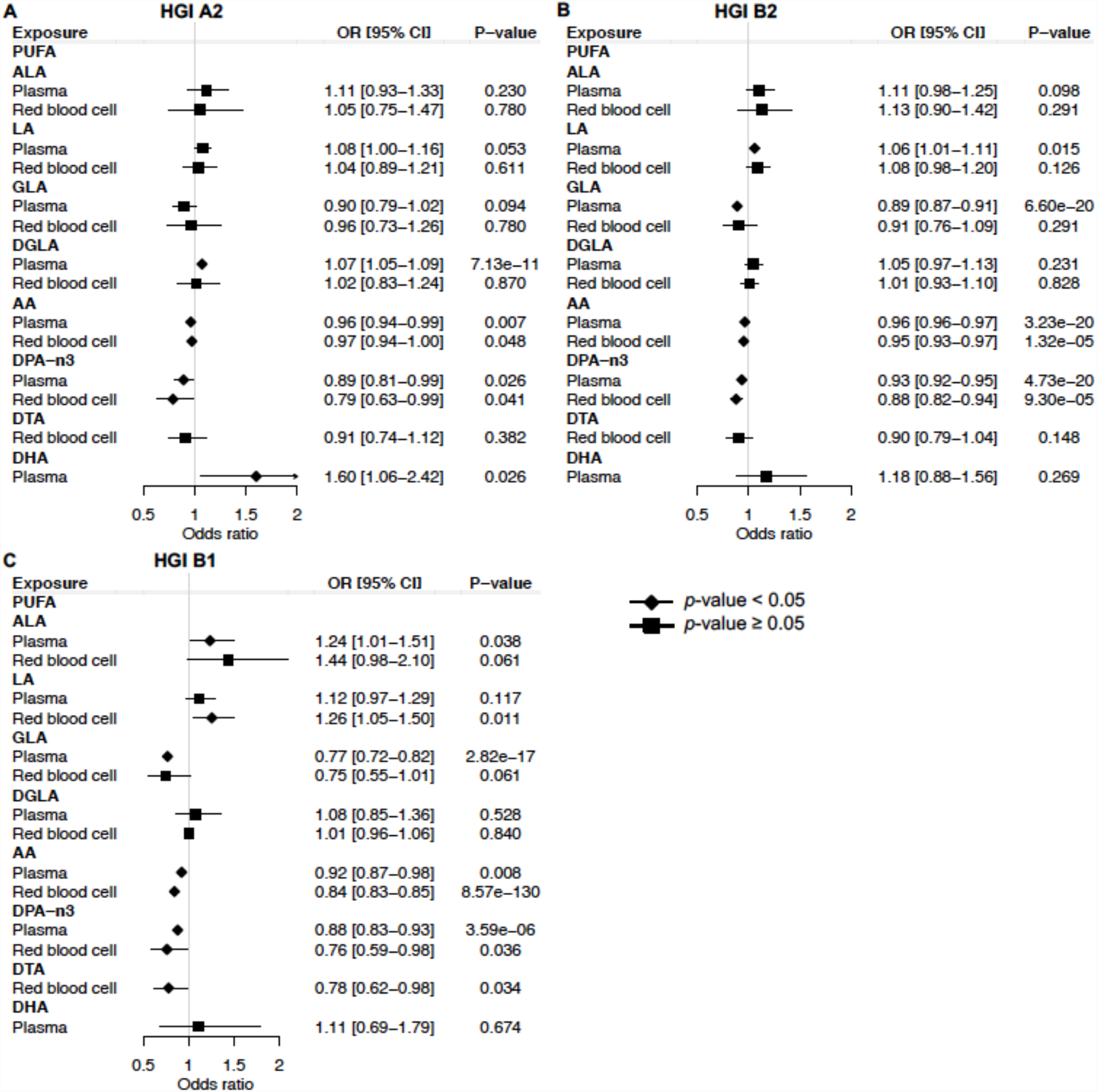
Mendelian randomization estimates of the effects of polyunsaturated fatty acids on COVID-19 severity risk. (A) Mendelian randomization analysis based on the release 5 HGI A2. (B) Mendelian randomization analysis based on the release 5 HGI B2. (C) Mendelian randomization analysis based on the release 5 HGI B1. Odds ratios are scaled to a genetically predicted SD increase in polyunsaturated fatty acids. Associations with *p*-value < 0.05 were indicated with diamonds, while others with squares. Detailed summary statistics are available in Supplemental Tables 6–8. PUFA, polyunsaturated fatty acid; ALA, α-Linolenic acid; LA: linoleic acid; GLA, γ-Linoleic acid; DGLA, dihomo-γ-linoleic acid; AA, arachidonic acid; DPA-n3, docosapentaenoic acid; DTA, docosatetraenoic acid; DHA, docosahexaenoic acid; OR, odds ratio.

In terms of COVID-19 susceptibility, we found that genetically instrumented one-SD increase of plasma DGLA (OR: 1.01; 95% CI: 1.00, 1.02; *P* = 0.031) was associated with an increased risk of any SARS-CoV-2 infection (Figure 3). MR analysis with RBC DGLA showed a similar pattern (OR: 1.01; 95% CI: 1.00, 1.02; *P* = 0.007). However, the association of genetically instrumented DGLA with the risk of testing positive for COVID-19 was not statistically significant using any other LD criteria for genetic instruments (Supplemental Table 9). Notably, our forward MR findings were confirmed using additional COVID-19 GWAS from HGI release 4 (Supplemental Tables 10–13). In summary, our forward MR analyses suggest that higher circulating levels of AA and DPA-n3 are associated with a lower risk of developing severe forms of COVID-19.

**Figure 3.**
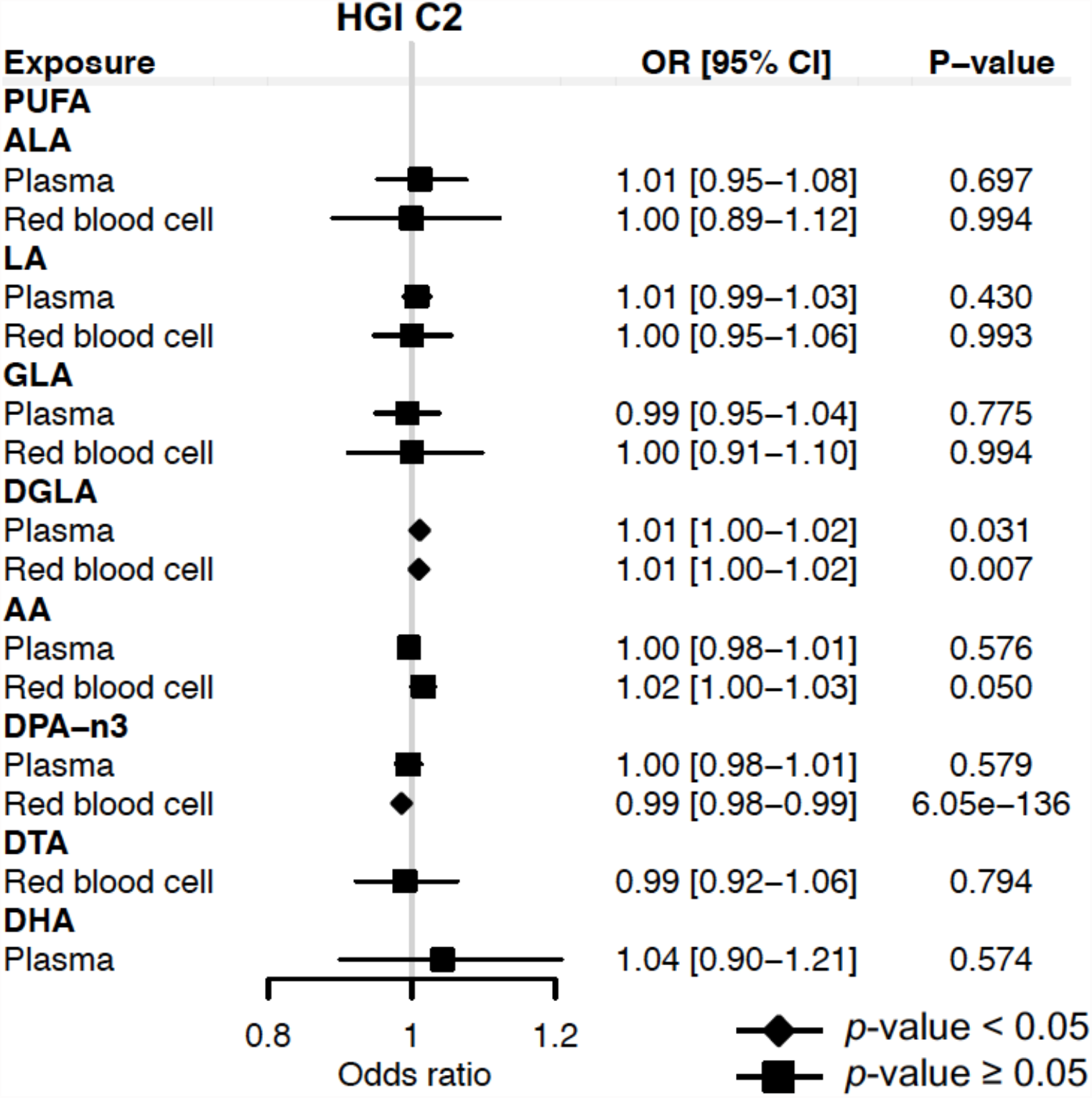
Mendelian randomization estimates of the effects of polyunsaturated fatty acids on COVID-19 susceptibility risk based on the release 5 HGI C2. Odds ratios are scaled to a genetically predicted SD increase in polyunsaturated fatty acids. Associations with *p*-value < 0.05 were indicated with diamonds, while others with squares. Detailed summary statistics are available in Supplemental Table 9. PUFA, polyunsaturated fatty acid; ALA, α-Linolenic acid; LA: linoleic acid; GLA, γ-Linoleic acid; DGLA, dihomo-γ-linoleic acid; AA, arachidonic acid; DPA-n3, docosapentaenoic acid; DTA, docosatetraenoic acid; DHA, docosahexaenoic acid; OR, odds ratio.

We further applied reverse MR analyses to investigate the causal effects of COVID-19 on each PUFA. Although several reverse MR analyses showed that genetically instrumented COVID-19 susceptibility or severity was associated with ALA, DHA, GLA, or DGLA, there was no consistent evidence for an effect of COVID-19 on these PUFAs using the conventional genome-wide significance threshold (*P* < 5 × 10^−8^) and the more lenient threshold (*P* < 5 × 10^−6^) for COVID-19 SNPs from HGI release 5 (Supplemental Tables 14–21). In addition, we used SNPs associated with COVID-19 from HGI release 4, and we did not observe any causal effect of COVID-19 on PUFAs (Supplemental Tables 22–29). Importantly, the reverse MR results showed no significant association of genetically predicted COVID-19 severity with AA and DPA-n3, suggesting that the significant forward MR results are unlikely to be confounded by reverse causation.

## Discussion

Our observational analysis in a prospective cohort showed that total PUFAs, omega-3 PUFAs, omega-6 PUFAs, DHA, and LA in baseline plasma samples were inversely associated with the risk of severe COVID-19. There were also inverse associations of omega-3 PUFAs and DHA with COVID-19 susceptibility. In contrast, the omega-6/omega-3 ratio was positively associated with both COVID-19 susceptibility and severity. In our bidirectional two-sample MR analyses, we provided evidence for the potential causal roles of higher circulating AA and DPA-n3 in a lower risk of COVID-19 severity.

Our observational findings are broadly consistent with previous observational studies and a pilot clinical trial. Julkunen *et al*. also examined the UK Biobank cohort, although with smaller sample sizes and different controls. They showed that for total PUFAs, omega-3 PUFAs, omega-6 PUFAs, DHA, and LA, their absolute levels and relative percentages in total fatty acids were both inversely associated with the risk of severe COVID-19 when comparing patients to non-cases with unknown COVID-19 status (13). Our study corrected for potential selection bias by restricting the analysis to individuals with COVID-19 testing status and used those with negative tests or non-hospitalized patients as the controls. We confirmed the same inverse association patterns for severe COVID-19. We further showed that omega-3 PUFAs and DHA were inversely associated with COVID-19 susceptibility. Another study investigated the metabolic fingerprint of COVID-19 severity in 581 samples from three cohorts, revealing inverse associations with severity for total PUFAs, omega-6 PUFAs, and LA. But inconsistent associations of omega-3 PUFAs, DHA, and the omega-6/omega-3 ratio were also observed across cohorts (11). Comparing the lipid profile of 42 severe COVID-19 patients to 22 healthy subjects, a study by Perez-Torres *et al*. found that plasma GLA, DGLA, and EPA were decreased in COVID-19 patients, but LA and AA were elevated (12). Two small studies found that the omega-3 index was significantly lower in COVID-19 patients and was inversely associated with risks of requiring mechanical ventilation and death (9, 10). The differences in these observational studies are likely results of uncontrolled confounding factors or the usage of patients at different disease stages. In support of the associated protective effect of omega-3 fatty acids, the first randomized clinical trial of supplementing 1000 mg omega-3 fatty acids in 128 critically ill COVID-19 patients showed that the intervention group has a significantly higher one-month survival rate and improved respiratory and renal function (45). Altogether with the existing literature, our study supports the protective effects of omega-3 fatty acids against the development of severe COVID-19 and likely also against viral infection. There are probably protective benefits of omega-6 fatty acids against severe COVID-19, but a high omega-6/omega-3 ratio may increase the risks of both COVID-19 susceptibility and severity.

In our MR study, we examined whether specific individual PUFAs play causal roles in COVID-19 susceptibility and severity. We found that genetically instrumented circulating levels of AA and DPA-n3 are associated with a lower risk of severe COVID-19. AA is an omega-6 fatty acid, while DPA-n3 is an omega-3 fatty acid. Although these two specific PUFAs were not available in our observational analysis, their potentially causal protective effects are consistent with the inverse associations of both omega-6 PUFAs and omega-3 PUFAs with severe COVID-19. The potential protective roles of AA and DPA-n3 in severe COVID-19 have mechanistic support. Both of them are well-known to serve as precursors of specialized pro-resolving mediators, such as lipoxins derived from AA, resolvins, protectins and maresins derived from DPA-n3, playing essential roles in promoting the resolution of inflammatory responses and tissue repair (5, 7, 46). Notably, it has been highlighted that the roles of AA in initiating timely inflammatory responses through its derived prostaglandins (PGs), such as PGE_2_, may be as important as its roles in inflammatory resolution through lipoxins (6, 47). Another possible mechanistic route for AA could be drawn from a human cell line study (48). Huh-7 cells, a hepatocyte-derived carcinoma cell line, when infected with human coronavirus 229E (HCoV-229E), exhibit significantly elevated levels of LA and AA, a pattern also observed in a study of severe COVID-19 patients (12). Interestingly, exogenous supplementation of LA and AA in HCoV-229E-infected cells significantly suppressed HCoV-229E virus replication. Similar suppressive effects were observed for the highly pathogenic Middle East respiratory syndrome coronavirus (MERS-CoV) (48), suggesting a possible general mechanism of LA and AA on coronavirus. Additionally, LA has been shown to directly and tightly bind the SARS-CoV-2 spike glycoprotein, reducing its interaction with the human ACE2 receptor (49). Similar inhibitory effects were observed for ALA, EPA, and DHA in a ligand screening study (50), which did not include AA and DPA-n3. Our MR findings call for future studies into the mechanistic roles of AA and DPA-n3 in the development of severe COVID-19.

Our study has a number of strengths and novel features. To our knowledge, this is the first MR study examining the causal effects of PUFAs on COVID-19. It is also the first MR study of PUFAs that used genetic variants for RBC PUFAs, in addition to plasma PUFAs. We applied bidirectional two-sample MR analyses to evaluate the direction of the causality and to rule out the impacts of reverse causation. To obtain robust evidence, we required the reported patterns to be observed with both plasma and RBC PUFAs. Similarly, to ensure reproducibility across data releases, we confirmed the results with analyses based on four COVID-19 GWAS (A2, B2, B1, and C2) from HGI releases 5 and 4. Bonferroni correction was used to overcome the issue of multiple testing. Another strength is the sensitivity analysis with various LD cutoffs. Additionally, comparing our MR results between severe COVID-19 and any SARS-CoV-2 infection, we found that AA and DPA-n3 might mainly impact the severity of disease progression but not susceptibility to viral infection.

Our study has several limitations. First, we could not completely rule out the possibility that some genetic variants might be pleiotropic, although we applied multiple sensitivity analyses, including the heterogeneity test, MR-Egger, and WM method. Second, a limitation of this MR study is that the effect of endogenous PUFAs may be different from the effect of dietary PUFAs, and our study did not directly examine dietary PUFAs. However, leveraging genetic instruments yields novel insights and minimizes the measurement error from self-reported dietary consumption in nutrition studies. Third, another limitation is that the population controls were utilized with no information on COVID-19 status in three COVID-19 GWAS used in our primary analysis, including the HGI A2, B2, and C2 studies. To mitigate this issue, we also utilized the HGI B1 study, which is another GWAS of COVID-19 using non-hospitalized patients as the control group. Fourth, in the observational study, UK Biobank recruited healthier individuals and thus may not be representative of the general population. Fifth, the NMR-based measurements of plasma PUFAs were collected over ten years before the COVID-19 pandemic, and the time lag likely attenuates the magnitude of association. Sixth, our observational study could be affected by ascertainment bias in differential healthcare seeking and testing. Seventh, our findings might not be extrapolated to other ethnicities because the study only focused on participants of European descent. Eighth, our study can not thoroughly explain the mechanisms. Further mechanistic research is necessary to investigate the biological pathways underpinning the roles of PUFAs in severe COVID-19.

In conclusion, our observational analysis in a prospective cohort shows that total PUFAs, omega-3 PUFAs, omega-6 PUFAs, DHA, and LA are inversely associated with the risk of severe COVID-19. Omega-3 and DHA may also be protective against SARS-CoV-2. A higher omega-6/omega-3 ratio has adverse effects on both COVID-19 susceptibility and severity. Our MR study further suggests a possible causal role of AA and DPA-n3 in reducing the risk of severe COVID-19. Our findings call for further studies into the mechanistic roles of PUFAs in COVID-19. They also support the possible usage of circulating PUFA levels as biomarkers for identifying high-risk individuals and as therapeutic targets for managing COVID-19 patients.

## Data Availability

The summary statistics for genetic instruments of PUFAs are available in the supplementary tables. The COVID-19 data (GWAS summary statistics) used in this study are freely accessible in the COVID-19 Host Genetics Initiative (https://www.covid19hg.org/). The code for the analyses is available at https://github.com/yitangsun/COVID19_PUFA_MR.

https://github.com/yitangsun/COVID19_PUFA_MR

## Acknowledgments

Our study was approved by the University of Georgia Institutional Review Board and the UK Biobank consortium (application no. 48818). We thank the investigators of the COVID-19 Host Genetics Initiative for sharing the GWAS summary statistics.

The authors’ responsibilities were as follows—YS and KY: designed the study, provided statistical advice, and interpreted the data; YS: performed data analysis and prepared visualizations; YS, RC, and AR: provided material support during the study; YS: wrote the paper; KY: critically revised the paper; and all authors: read and approved the final manuscript and took responsibility for the integrity of the work as a whole. The authors report no conflicts of interest.

## Data availability

The COVID-19 data (GWAS summary statistics) used in this study are freely accessible in the COVID-19 Host Genetics Initiative (https://www.covid19hg.org/). The code for the analyses is available at https://github.com/yitangsun/COVID19_PUFA_MR.

## Notes

Research reported in this publication was supported by the University of Georgia Research Foundation and by the National Institute of General Medical Sciences of the National Institutes of Health under Award Number R35GM143060. The content is solely the responsibility of the authors and does not necessarily represent the official views of the National Institutes of Health or the University of Georgia.

Supplemental Tables 1–29 are available from the “Supplementary data” link in the online posting of the article and from the same link in the online table of contents at https://academic.oup.com/ajcn/.

### Abbreviations used

COVID-19: coronavirus disease 2019
SARS-CoV-2: severe acute respiratory syndrome coronavirus 2
PUFAs: polyunsaturated fatty acids
LA: linoleic acid
EPA: eicosapentaenoic acid
DHA: docosahexaenoic acid
RBC: red blood cell
AA: arachidonic acid
MR: Mendelian randomization
BMI: body mass index
GWAS: genome-wide association studies
HGI: COVID-19 Host Genetics Initiative
FHS: Framingham Heart Study
STROBE: strengthening the reporting of observational studies in epidemiology
STROBE-MR: strengthening the reporting of Mendelian randomization studies
ALA: α-Linolenic acid
DPA-n3: docosapentaenoic acid
GLA: γ-Linoleic acid
DGLA: dihomo-γ-linoleic acid
DTA: docosatetraenoic acid
NMR: nuclear magnetic resonance
SD: standard deviation
SNPs: single nucleotide polymorphisms
LD: linkage disequilibrium
OR: odds ratio
IVW: inverse variance-weighted
WM: weighted median
CI: confidence interval
PGs: prostaglandins
HCoV-229E: human coronavirus 229E
MERS-CoV: Middle East respiratory syndrome coronavirus.

## Supplemental Tables

Supplemental Table 1. Notable Mendelian randomization studies of polyunsaturated fatty acids.

Supplemental Table 2. STROBE Statement—checklist of items that should be included in reports of cohort studies.

Supplemental Table 3. STROBE-MR checklist.

Supplemental Table 4. Genetic instruments for plasma polyunsaturated fatty acids.

Supplemental Table 5. Significant SNPs for red blood cell polyunsaturated fatty acids.

Supplemental Table 6. Forward Mendelian randomization estimates of associations of genetically predicted polyunsaturated fatty acids with COVID-19 severity based on the release 5 HGI A2.

Supplemental Table 7. Forward Mendelian randomization estimates of associations of genetically predicted polyunsaturated fatty acids with COVID-19 severity based on the release 5 HGI B2.

Supplemental Table 8. Forward Mendelian randomization estimates of associations of genetically predicted polyunsaturated fatty acids with COVID-19 severity based on the release 5 HGI B1.

Supplemental Table 9. Forward Mendelian randomization estimates of associations of genetically predicted polyunsaturated fatty acids with COVID-19 susceptibility based on the release 5 HGI C2.

Supplemental Table 10. Forward Mendelian randomization estimates of associations of genetically predicted polyunsaturated fatty acids with COVID-19 severity based on the release 4 HGI A2.

Supplemental Table 11. Forward Mendelian randomization estimates of associations of genetically predicted polyunsaturated fatty acids with COVID-19 severity based on the release 4 HGI B2.

Supplemental Table 12. Forward Mendelian randomization estimates of associations of genetically predicted polyunsaturated fatty acids with COVID-19 severity based on the release 4 HGI B1.

Supplemental Table 13. Forward Mendelian randomization estimates of associations of genetically predicted polyunsaturated fatty acids with COVID-19 susceptibility based on the release 4 HGI C2.

Supplemental Table 14. Reverse Mendelian randomization estimates of associations of genetically predicted COVID-19 severity with polyunsaturated fatty acids based on the release 5 HGI A2 (COVID-19 SNP P < 5e-8).

Supplemental Table 15. Reverse Mendelian randomization estimates of associations of genetically predicted COVID-19 severity with polyunsaturated fatty acids based on the release 5 HGI B2 (COVID-19 SNP P < 5e-8).

Supplemental Table 16. Reverse Mendelian randomization estimates of associations of genetically predicted COVID-19 severity with polyunsaturated fatty acids based on the release 5 HGI B1 (COVID-19 SNP P < 5e-8).

Supplemental Table 17. Reverse Mendelian randomization estimates of associations of genetically predicted COVID-19 susceptibility with polyunsaturated fatty acids based on the release 5 HGI C2 (COVID-19 SNP P < 5e-8).

Supplemental Table 18. Reverse Mendelian randomization estimates of associations of genetically predicted COVID-19 severity with polyunsaturated fatty acids based on the release 5 HGI A2 (COVID-19 SNP P < 5e-6).

Supplemental Table 19. Reverse Mendelian randomization estimates of associations of genetically predicted COVID-19 severity with polyunsaturated fatty acids based on the release 5 HGI B2 (COVID-19 SNP P < 5e-6).

Supplemental Table 20. Reverse Mendelian randomization estimates of associations of genetically predicted COVID-19 severity with polyunsaturated fatty acids based on the release 5 HGI B1 (COVID-19 SNP P < 5e-6).

Supplemental Table 21. Reverse Mendelian randomization estimates of associations of genetically predicted COVID-19 susceptibility with polyunsaturated fatty acids based on the release 5 HGI C2 (COVID-19 SNP P < 5e-6).

Supplemental Table 22. Reverse Mendelian randomization estimates of associations of genetically predicted COVID-19 severity with polyunsaturated fatty acids based on the release 4 HGI A2 (COVID-19 SNP P < 5e-8).

Supplemental Table 23. Reverse Mendelian randomization estimates of associations of genetically predicted COVID-19 severity with polyunsaturated fatty acids based on the release 4 HGI B2 (COVID-19 SNP P < 5e-8).

Supplemental Table 24. Reverse Mendelian randomization estimates of associations of genetically predicted COVID-19 severity with polyunsaturated fatty acids based on the release 4 HGI B1 (COVID-19 SNP P < 5e-8).

Supplemental Table 25. Reverse Mendelian randomization estimates of associations of genetically predicted COVID-19 susceptibility with polyunsaturated fatty acids based on the release 4 HGI C2 (COVID-19 SNP P < 5e-8).

Supplemental Table 26. Reverse Mendelian randomization estimates of associations of genetically predicted COVID-19 severity with polyunsaturated fatty acids based on the release 4 HGI A2 (COVID-19 SNP P < 5e-6).

Supplemental Table 27. Reverse Mendelian randomization estimates of associations of genetically predicted COVID-19 severity with polyunsaturated fatty acids based on the release 4 HGI B2 (COVID-19 SNP P < 5e-6).

Supplemental Table 28. Reverse Mendelian randomization estimates of associations of genetically predicted COVID-19 severity with polyunsaturated fatty acids based on the release 4 HGI B1 (COVID-19 SNP P < 5e-6).

Supplemental Table 29. Reverse Mendelian randomization estimates of associations of genetically predicted COVID-19 susceptibility with polyunsaturated fatty acids based on the release 4 HGI C2 (COVID-19 SNP P < 5e-6).

