## Supplementary Tables for "Circulating polyunsaturated fatty acids and COVID-19: a prospective cohort study and Mendelian randomization analysis"

### Supplementary Table 1. Notable Mendelian randomization studies of polyunsaturated fatty acids.

| MR study | Exposure | Exposure data sources | Exposure unit | Outcome | Outcome data sources | Findings (increasing risk, decreasing risk, null) |
|---|---|---|---|---|---|---|
| Zhao, J.V., Schooling, C.M (1) | LA | CHARGE consortium | % total increase in total fatty acids | IHD, diabetes, HDL-C, LDL-C, TC, BP, and reticulocyte count | CARDIoGRAMplusC4D 1000 Genomes, the Myocardial Infarction Genetics and CARDIoGRAM Exome, the UK Biobank SOFT CAD GWAS, the DIAbetes Genetics Replication And Meta-analysis diabetes case study, Global Lipids Genetics Consortium, and the UK Biobank | Genetically predicted higher serum LA was associated with the lower diabetes risk, lower levels of HDL-C, LDL-C, and TC. Genetically predicted LA was not associated with IHD, SBP, or reticulocyte count. |
| Zhang, T. *et al.* (2) | AA | CHARGE consortium | SD | IHD, stroke, HDL-C, LDL-C, TG, ApoB, ApoA-I, BP, adiposity, and markers of inflammation and coagulation | The CARDIoGRAMplusC4D 1000 Genomes, MEGASTROKE consortium, the UK Biobank pan-ancestry summary statistics, GLGC, the Genetic Investigation of ANthropometric Traits Consortium and the UK Biobank meta-analysis, a GWAS of cytokines in 8293 Finnish individuals, the INTERVAL study in 3301 individuals of European ancestry, the CHARGE Inflammation Working Group, and the Biobank Japan. | Genetically predicted AA was associated with higher levels of ApoB, HDL-C, LDL-C, and a lower level of TG, but not associated with IHD, stroke, ApoA-I, BP, adiposity, or other markers of inflammation and coagulation. |
| Yuan, S. *et al.* (3) | ALA, EPA, DPA, DHA, LA, AA, POA, OA, PA, and SA | CHARGE consortium | SD | CAD, IS, AF, heart failure, aortic valve stenosis, abdominal and thoracic aortic aneurysms, transient ischemic attack, intracerebral and subarachnoid hemorrhages, VTE, and PAD | CARDIoGRAMplusC4D consortium, MEGASTROKE consortium, Atrial Fibrillation consortium, and UK Biobank | Genetically predicted plasma AA and SA levels were associated with the higher CVD risk. Genetically predicted higher plasma ALA, LA, and OA levels were associated with lower risks of large-vessel stroke and venous thromboembolism. |
| Yuan, T. *et al.* (4) | LA, AA, ALA, EPA, | CHARGE consortium | SD | IS, LDL-C, HDL-C, TC, SBP, and DBP | The International Stroke Genetics Consortium, the GLGC, and the UK Biobank | Genetically predicted AA was associated with higher IS risk, higher levels of LDL-C, HDL-C, and TC. Genetically predicted serum ALA was |

| | | | | | | |
|---|---|---|---|---|---|---|
| | DHA, and DPA | | | | | associated with a lower IS risk, lower levels of LDL-C, HDL-C, and TC. Genetically predicted LA, EPA, DHA, and DPA were not associated with IS, lipids, or BP. |
| Liao, L. Z. *et al.* (5) | Omega-6 fatty acids | Large GWAS (n = 24,925) from 14 cohorts in people of European ancestry | SD | CHD, TC, LDL-C, HDL-C, and TG | CARDIoGRAMplusC4D and GLGC | Genetically predicted omega-6 was associated with a higher risk of CHD, which might be partially mediated by TC, LDL-C, and TG. |
| Mazidi, M. *et al.* (6) | 10-heptadecenoate, MA, OA, and POA | A GWAS (including 7,824 adult samples of European ancestry) | SD | CHD, MI, CS, and IS | CARDIoGRAMplusC4D 1000 Genomes, UK Biobank SOFT CAD study, two small case (n = 4,120)–control (n = 3,910) studies from Germany and Greece, and METASTROKE | Genetically predicted serum 10-heptadecenoate, MA, OA, and POA were not associated with risks of CHD, MI, CS, or IS. |
| Zhang, T. *et al.* (7) | AA | CHARGE consortium | % total increase in fatty acid | ASCVD, IHD, PAD, VTE, and other CVD | CARDIoGRAMplusC4D 1000 Genomes, MEGASTROKE, Pan-UK Biobank, Atrial Fibrillation Consortium, HERMES consortium, and FinnGen. | Genetically predicted AA was associated with higher risks of ASCVD, IHD, PAD, and VTE, with possibly stronger associations in men than women. |
| Yuan, S., & Larsson, S. C. (8) | ALA, DHA, EPA, DPA, AA, and LA | CHARGE consortium | SD | AF | Atrial Fibrillation Consortium | Genetically predicted ALA, DHA, EPA, DPA, AA, and LA were not associated with the risk of AF. |
| Park, S. *et al.* (9) | DPA, EPA, DHA, LA, GLA, DGLA, AdrA, and AA | CHARGE consortium | SD | CAD and MI | UK Biobank and CARDIoGRAMplusC4D | Genetically predicted AA was associated with a higher risk of CAD. Genetically predicted higher EPA, LA, and DGLA levels were associated with a lower risk of CAD. |
| Wang, W *et al.* (10) | MUFA and omega-3 fatty acids | Large GWAS (n = 24,925) from 14 cohorts in people of European ancestry | SD | Incident heart failure and mortality of heart failure | Cohorts for Heart and Aging Research in Genomic Epidemiology-Heart Failure Working Group | Genetically predicted higher circulating MUFA levels were associated with increased risk of heart failure, but no association with mortality of heart failure. Genetically predicted omega-3 fatty acids were not associated with heart failure. |
| Chen, H. Y. *et al.* (11) | AA | FOS cohort (n = 1310, 492 cases of AVC) and European-ancestry participants in the MESA cohort (n = 2415) | SD | AVC and AS | Genetic Epidemiology Research on Adult Health and Aging cohort and CHARGE Consortium | Genetically predicted higher AA level was associated with higher risks of AS and AVC. |

| | | | | | | |
|---|---|---|---|---|---|---|
| Yuan, S. *et al.* (12) | ALA, EPA, DHA, LA, AA, POA, OA, PA, and SA | CHARGE consortium | SD | T2DM | DIAGRAM consortium | Genetically predicted higher EPA, DPA, AA, and SA were associated with a higher risk of T2DM. Genetically predicted higher ALA, LA, POA, and OA levels were associated with a lower risk of T2DM. |
| Jäger, S. *et al.* (13) | D5D (AA/DGLA) and D6D (GLA/LA) | EPIC–Potsdam Study | SD | T2DM and CAD | DIAGRAM consortium and CARDIoGRAM. | Genetically predicted higher D6D and D5D were associated with higher risks of T2DM and CAD. |
| Adams, C. D., & Neuhausen, S. L. (14) | TOTFA, MUFA, omega-3 fatty acids, and PUFA | Large GWAS (n = 24,925) from 14 cohorts in people of European ancestry | SD | Chronotype and T2DM | UK Biobank and DIAGRAM consortium | Genetically predicted higher TOTFA and MUFA levels were associated with a lower risk of T2DM. |
| Zulyniak *et al.* (15) | AA | CHARGE consortium | SD | DI and T2DM | The Meta-Analysis of Glucose- and Insulin-Related Traits Consortium and the UK Biobank | Genetically predicted higher AA was associated with DI. Genetically predicted AA was not associated with T2DM. |
| Ma, M. *et al.* (16) | ALA, EPA, DPA, DHA, LA, and AA | CHARGE consortium | SD | BP, SBP, DBP, and PP | UK Biobank and International Consortium for Blood Pressure | Genetically predicted higher ALA was associated with higher DBP. Genetically predicted higher AA and EPA were associated with lower DBP. Genetically predicted ALA, EPA, DPA, DHA, LA, and AA were not associated with SBP. Genetically predicted DPA and DHA were not associated with DBP. |
| Zhao, J. V., & Schooling, C. M. (17) | LA | Large GWAS (n = 24,925) from 14 cohorts in people of European ancestry | SD | Asthma, eosinophil, neutrophil, and monocyte counts | Case-control study of asthma based on Trans-National Asthma Genetic Consortium and UK Biobank | Genetically predicted higher LA was associated with a lower risk of asthma, lower eosinophil count, and lower neutrophil count. |
| Liyanage, U. *et al.* (18) | EPA, ALA, LA, AA, DHA, and DPA | CHARGE consortium | SD | Overall cancer risk and cancer mortality | UK Biobank | Genetically predicted higher AA was associated with an increased risk for colorectal cancer. Genetically predicted EPA, ALA, LA, AA, DHA, and DPA were not associated with overall cancer risk or mortality. |
| Larsson, S. *et al.* (19) | AA, EPA, and DHA | CHARGE consortium | SD | Ten site-specific cancers (esophagus, stomach, | BioBank Japan, Breast Cancer Association Consortium, ILCCO, Ovarian Cancer Association Consortium, Prostate Cancer Association Group to Investigate Cancer | Genetically predicted higher plasma phospholipid AA concentrations were associated with increased risks of colorectal cancer, lung cancer, and esophageal cancer. Genetically predicted AA, EPA, and DHA were not |

| | | | | | | |
|---|---|---|---|---|---|---|
| | | | | colorectum, pancreas, lung, bladder, prostate, breast, uterus, and ovaries) | Associated Alterations in the Genome consortium | associated with cancers of the stomach, pancreas, bladder, prostate, breast, uterus, or ovary. |
| Liu, J. *et al.* (20) | DPA | CHARGE consortium | % total increase in fatty acid | Lung cancer risk | ILCCO | Genetically predicted higher DPA was associated with an increased risk for lung cancer. |
| Shen, J. *et al.* (21) | Other PUFA than 18:2 in blood, DHA, DPA, EPA, AA, DGLA, and LA | Large GWAS (n = 24,925) from 14 cohorts in people of European ancestry and a GWAS (including 7,824 adult samples of European ancestry) | SD | Lung cancer, LUAD, and LUSC | ILCCO | Genetically predicted higher other PUFA than 18:2, DPA, EPA, and AA were associated with an increased risk for lung cancer. |
| Yang, Z. *et al.* (22) | ALA, EPA, DPA, DHA, LA, AA, POA, OA, PA, and SA | CHARGE consortium | SD | Breast cancer and prostate cancer | Breast Cancer Association Consortium, Prostate Cancer Association Group to Investigate Cancer-Associated Alterations in the Genome consortium | Genetically predicted ALA, EPA, DPA, DHA, LA, AA, POA, OA, PA, and SA were not associated with breast cancer or prostate cancer. |
| Guo, F. *et al.* (23) | SFA, MUFA, and PUFA | Large GWAS (n = 24,925) from 14 cohorts in people of European ancestry | SD | Breast cancer | The National Health and Nutrition Examination Survey | Genetically predicted MUFA was associated with a reduced risk of breast cancer. |
| Khankari, N., Murff, H., Zeng, C. *et al.* (24) | LA, AA, ALA, EPA, DPA, and DHA | CHARGE consortium | SD | Prostate cancer | Prostate Cancer Association Group to Investigate Cancer Associated Alterations in the Genome | Genetically predicted LA, AA, ALA, EPA, DPA, and DHA were not associated with prostate cancer risk. |
| Ghoneim, D. H. *et al.* (25) | LA, AA, AdrA, GLA, and DGLA | CHARGE consortium | SD | Pancreatic cancer | Pancreatic Cancer Cohort Consortium and the Pancreatic Cancer Case-Control Consortium | Genetically predicted LA, AA, AdrA, GLA, and DGLA were not associated with pancreatic cancer risk. |
| Seviiri, M. *et al.* (26) | ALA, EPA, DPA, DHA, AA, and LA | CHARGE consortium | SD | Keratinocyte cancer, BCC, and SCC | UK Biobank, 23andMe, and QSkin Sun and Health Study cohort | Genetically predicted higher AA and EPA were associated with a higher BCC risk. Genetically predicted higher LA and ALA were associated with a reduced BCC risk. |

| | | | | | | |
|---|---|---|---|---|---|---|
| Khankari, N. K. et al. (27) | LA, AA, ALA, EPA, DPA, and DHA | CHARGE Consortium | % total increase in fatty acid | CRC | Colorectal Cancer Consortium | Genetically predicted higher AA, EPA, and DPA were associated with an increased CRC risk. Genetically predicted higher LA and ALA were associated with a reduced CRC risk. |
| May-Wilson, S. *et al.* (28) | EPA, DPA, DHA, LA, AA, DGLA, OA, POA, PA, and SA | CHARGE consortium | % total increase in fatty acid | CRC | Colon Cancer Family Registry, COIN trial, Finnish Colorectal Cancer Predisposition Study, colorectal Tumour Gene Identification Consortium, Colorectal Cancer Susceptibility Study, and VQ58 | Genetically predicted higher AA and SA were associated with an increased CRC risk. Genetically predicted higher LA, OA, and POA were associated with reduced CRC risk. |
| Isom, C.A. *et al.* (29) | AA | Tennessee Colorectal Polyps Study | % total increase in fatty acid | Colorectal adenoma | Tennessee Colorectal Polyps Study | Genetically predicted AA was not associated with colorectal adenoma risk. |
| Liyanage, U. *et al.* (30) | EPA, DPA, DHA, ALA, AA, and LA | CHARGE consortium | SD | Melanoma | Melanoma risk meta-analysis consisting of 11 GWAS | Genetically predicted plasma EPA, DPA, DHA, ALA, AA, and LA were not associated with melanoma risk. |
| Saunders, C. N. *et al.* (31) | MUFA, omega-3, and omega-6 fatty acids | Large GWAS (n = 24,925) from 14 cohorts in people of European ancestry | SD | Glioma | A GWAS of glioma | Genetically predicted higher MUFA and omega-3 fatty acids were associated with a reduced glioma sk. |
| Yuan, S. *et al.* (32) | ALA, EPA, DPA, DHA, LA, AA, POA, OA, PA, and SA | CHARGE Consortium | SD | Fracture and eBMD | UK Biobank | Genetically predicted higher plasma ALA, LA, PA, and OA levels were associated with an increased eBMD risk. Genetic predisposition to higher plasma AA, EPA, DPA, and SA levels were associated with an increased risk of fracture. Genetically predicted higher ALA, LA, PA, and OA levels were associated with a reduced risk of fracture. Genetically predicted higher plasma AA, EPA, DPA, and SA levels were associated with a lower risk of eBMD. Genetically predicted plasma DHA and PA were not associated with the risk of eBMD or fracture. |
| Zhao, J. V., & Schooling, C. M. (33) | LA | CHARGE consortium | NA | RA and SLE | UK Biobank, Rheumatoid Arthritis Consortium, and ImmunoBase Consortium | Genetically predicted higher LA was associated with reduced risks of RA and SLE. |
| Sun, L. *et al.* (34) | POA and OA | CHARGE consortium | SD | RA and osteoarthritis | A trans-ethnic GWASs and UK Biobank | Genetically predicted higher plasma levels of POA and OA were associated with a lower RA |

| | | | | | | |
|---|---|---|---|---|---|---|
| | | | | | | risk. Genetically predicted plasma POA and OA were not associated with osteoarthritis risk. |
| Wang *et al.* (35) | LA and AA | CHARGE consortium | % total increase in total fatty acids | AMD | International AMD Genomics Consortium | Genetically predicted higher AA was associated with an increased AMD risk. Genetically predicted higher plasma LA was associated with a lower risk of AMD. |
| Carreras-Torres, R. *et al.* (36) | PUFA, omega-3 and omega-6 fatty acids, MUFA, and TFA | Large GWAS (n = 24,925) from 14 cohorts in people of European ancestry | SD | IBD, CD, and UC | IBD Genetics Consortium and the UK IBD Genetics Consortium | Genetically predicted higher omega-3 fatty acids levels were associated with a lower risk of CD. |
| Cheng, T. S. *et al.* (37) | DGLA and PA | EPIC–Potsdam Study | SD | Puberty timing | UK Biobank | Genetically predicted higher DGLA was associated with earlier menarche in girls. |
| Milanesch i, Y. *et al.* (38) | Omega-3 fatty acids | Large GWAS (n = 24,925) from 14 cohorts in people of European ancestry | SD | MDD | The Psychiatric Genomics Consortium | Genetically predicted omega-3 fatty acids were not associated with MDD risk. |
| Tomata *et al.* (39) | LA, AA, ALA, EPA, DPA, and DHA | CHARGE consortium | SD | Late-onset AD | Alzheimer Disease Genetics Consortium, CHARGE, The European Alzheimer's Disease Initiative, and Genetic and Environmental Risk in AD/Defining Genetic, Polygenic and Environmental Risk for Alzheimer's Disease Consortium | Genetically predicted LA, AA, ALA, EPA, DPA, and DHA were not associated with late-onset AD risk. |
| Wang, Z. *et al.* (40) | AA, cis-trans-18:2, DGLA, DPA, EPA, GLA, LA, PA, and SA | CHARGE consortium | SD | AD | International Genomics of Alzheimer's Project | Genetically predicted higher plasma LA was slightly associated with a lower risk of AD. |
| Jones, H. J. *et al.* (41) | ALA, EPA, DPA, DHA, LA, AA, and AdrA | CHARGE consortium | SD | Schizophrenia | Schizophrenia Working Group of the Psychiatric Genomics Consortium | Genetically predicted higher ALA was associated with the increased schizophrenia risk. Genetically predicted higher plasma DHA was associated with a lower risk of schizophrenia. |
| Sallis, H. *et al.* (42) | DHA and EPA | ALSPAC cohort | % total increase in total fatty acids | Perinatal onset depression, antenatal depression, | ALSPAC cohort | Genetically predicted EPA and DHA were not associated with depression risk. |

| | | | | | | |
|---|---|---|---|---|---|---|
| | | | | and postnatal depression. | | |
| Thompson, A.D. *et al.* (43) | DHA, LA, omega-3, and omega-6 fatty acids | Large GWAS (n = 24,925) from 14 cohorts in people of European ancestry | SD | PE's and psychotic disorder | The Psychosis-LIKe Symptoms interview | Genetically predicted DHA, LA, omega-3, and omega-6 fatty acids were not associated with PE's or psychotic disorder. |

LA: linoleic acid; AA, arachidonic acid; ALA, α-Linolenic acid; EPA, eicosapentaenoic acid; DPA, docosapentaenoic acid; DHA, docosahexaenoic acid; POA, palmitoleic acid; MA, myristoleic acid; OA, oleic acid; PA, palmitic acid; SA, stearic acid; GLA, γ-Linoleic acid; DGLA, dihomo-γ-linoleic acid; AdrA, adrenic acid; MUFA, monounsaturated fatty acids; D5D, Δ5-desaturase; D6D, Δ6-desaturase; TOTFA, total fatty acids; PUFA, polyunsaturated fatty acids; SFA, saturated fatty acids; CHARGE, Cohorts for Heart and Aging Research in Genomic Epidemiology; FOS, Framingham Offspring Study; MESA, Multi-Ethnic Study of Atherosclerosis; EPIC, European Prospective Investigation into Cancer and Nutrition; ALSPAC, the Avon Longitudinal Study of Parents and Children; SD, standard deviation; IHD, ischemic heart disease; HDL-C, high-density lipoprotein cholesterol; LDL-C, low-density lipoprotein cholesterol; TC, total cholesterol; TG, triglycerides; BP, blood pressure; SBP, systolic blood pressure; DBP, diastolic blood pressure; ApoA-I, apolipoprotein A-I; ApoB, apolipoprotein B; CAD, coronary artery disease; AF, atrial fibrillation; IS, ischemic stroke; MI, myocardial infarction; CS, cardioembolic stroke; IS, ischemic stroke; ASCVD, atherosclerotic cardiovascular diseases; PAD, peripheral artery disease; CVD, cardiovascular diseases; VTE, venous thromboembolism; MI, myocardial infarction; AVC, aortic valve calcium; AS, aortic stenosis; T2DM, Type 2 diabetes; DI, insulin disposition index; PP, pulse pressure; LUAD, lung adenocarcinoma; LUSC, lung squamous cell carcinoma; BCC, basal cell carcinoma; SCC, squamous cell carcinoma; CRC, colorectal cancer; eBMD, estimated bone mineral density; RA, rheumatoid arthritis; SLE, systemic lupus erythematosus; AMD, age-related macular degeneration; IBD, inflammatory bowel diseases; CD, Crohn's disease; UC, ulcerative colitis; MDD, major depressive disorder; AD, Alzheimer's disease; PE's, later psychotic experiences; GLGC, Global Lipids Genetics Consortium; DIAGRAM, DIAbetes Genetics Replication And Meta-analysis; CARDIoGRAM, Coronary ARtery DIsease Genome wide Replication and Meta-analysis; ILCCO, International Lung Cancer Consortium.

**Supplementary Table 2. STROBE Statement—checklist of items that should be included in reports of cohort studies.**

| | Item No. | Recommendation | Reported on page and line | Reported on section and paragraph |
|---|---|---|---|---|
| **Title and abstract** | 1 | (a) Indicate the study's design with a commonly used term in the title or the abstract | Page 1, lines 1-2; page 2, lines 25-26 | Title: "Circulating polyunsaturated fatty acids and COVID-19: a prospective cohort study and Mendelian randomization analysis"; abstract, paragraph 3 |
| | | (b) Provide in the abstract an informative and balanced summary of what was done and what was found | Pages 2-3, lines 20-44 | Abstract, paragraphs 1-5 |
| **Introduction** | | | | |
| Background/rationale | 2 | Explain the scientific background and rationale for the investigation being reported | Pages 4-5, lines 49-78 | Introduction, paragraphs 1-2 |
| Objectives | 3 | State specific objectives, including any prespecified hypotheses | Page 6, lines 93-97 | Introduction, paragraph 4 |
| **Methods** | | | | |
| Study design | 4 | Present key elements of study design early in the paper | Page 7, lines 116-122 | Methods, section of "Participants and study design" |
| Setting | 5 | Describe the setting, locations, and relevant dates, including periods of recruitment, exposure, follow-up, and data collection | Page 7, lines 116-122; pages 8-9, lines 134-147 | Methods, section of "Participants and study design"; section of "Observational analysis" |
| Participants | 6 | (a) Give the eligibility criteria, and the sources and methods of selection of participants. Describe methods of follow-up | Page 7, lines 116-122; pages 8-9, lines 134-147 | Methods, section of "Participants and study design"; section of "Observational analysis"; Figure 1 |
| | | (b) For matched studies, give matching criteria and number of exposed and unexposed | NA | NA |
| Variables | 7 | Clearly define all outcomes, exposures, predictors, potential confounders, and effect modifiers. Give diagnostic criteria, if applicable | Page 7, lines 116-122; pages 8-9, lines 147-156 | Methods, section of "Participants and study design"; section of "Observational analysis" |
| Data sources/ measurement | 8* | For each variable of interest, give sources of data and details of methods of assessment (measurement). Describe comparability of assessment methods if there is more than one group | Pages 8-9, lines 134-156 | Methods, section of "Observational analysis"; Figure 1 |
| Bias | 9 | Describe any efforts to address potential sources of bias | Page 8, lines 143-152 | Methods, section of "Observational analysis" |
| Study size | 10 | Explain how the study size was arrived at | Pages 8-9, lines 134-152 | Methods, section of "Observational analysis"; Figure 1 |

| | | | | |
|---|---|---|---|---|
| Quantitative variables | 11 | Explain how quantitative variables were handled in the analyses. If applicable, describe which groupings were chosen and why | Pages 9, lines 155-156 | Methods, section of “Observational analysis” |
| Statistical methods | 12 | (a) Describe all statistical methods, including those used to control for confounding | Page 9, lines 152-156 | Methods, section of “Observational analysis” |
| | | (b) Describe any methods used to examine subgroups and interactions | Pages 8-9, lines 147-152 | Methods, section of “Observational analysis”; Figure 1 |
| | | (c) Explain how missing data were addressed | Page 8, lines 156-157 | Methods, section of “Observational analysis”; Figure 1 |
| | | (d) If applicable, explain how loss to follow-up was addressed | Pages 8-9, lines 134-138; 152-157 | Methods, section of “Observational analysis”; Figure 1 |
| | | (e) Describe any sensitivity analyses | Pages 8-9, lines 147-152 | Methods, section of “Observational analysis” |
| **Results** | | | | |
| Participants | 13* | (a) Report numbers of individuals at each stage of study—eg numbers potentially eligible, examined for eligibility, confirmed eligible, included in the study, completing follow-up, and analysed | Page 13, lines 229-239 | Results, section of “Baseline characteristics”; Figure 1; Table 1 |
| | | (b) Give reasons for non-participation at each stage | Page 8, lines 156-157 | Methods, section of “Observational analysis” |
| | | (c) Consider use of a flow diagram | Page 13, lines 229-230 | Results, section of “Baseline characteristics”; Figure 1 |
| Descriptive data | 14* | (a) Give characteristics of study participants (eg demographic, clinical, social) and information on exposures and potential confounders | Page 13, lines 229-239 and 241-245 | Results, section of “Baseline characteristics” and “Observational association analysis”; Table 1 |
| | | (b) Indicate number of participants with missing data for each variable of interest | Page 8, lines 156-157; Page 13, lines 229-239 and 241-245 | Methods, section of “Observational analysis” Results, section of “Baseline characteristics” and “Observational association analysis”; Figure 1; Table 1 |
| | | (c) Summarise follow-up time (eg, average and total amount) | NA | NA |
| Outcome data | 15* | Report numbers of outcome events or summary measures over time | Page 13, lines 229-239; pages 13-14, lines 241-261 | Results, section of “Baseline characteristics”; section of “Observational association analysis”; Figure 1; Tables 1-2 |
| Main results | 16 | (a) Give unadjusted estimates and, if applicable, confounder-adjusted estimates and their precision (eg, 95% | Pages 13-14, lines 241-273 | Results, section of “Observational association analysis”; Table 2 |

| | | | | |
|---|---|---|---|---|
| | | confidence interval). Make clear which confounders were adjusted for and why they were included | | |
| | | (b) Report category boundaries when continuous variables were categorized | NA | NA |
| | | (c) If relevant, consider translating estimates of relative risk into absolute risk for a meaningful time period | NA | NA |
| Other analyses | 17 | Report other analyses done—eg analyses of subgroups and interactions, and sensitivity analyses | Pages 13-14, lines 241-273 | Results, section of "Observational association analysis"; Table 2 |
| **Discussion** | | | | |
| Key results | 18 | Summarise key results with reference to study objectives | Page 18, lines 331-335 | Discussion, paragraph 1 |
| Limitations | 19 | Discuss limitations of the study, taking into account sources of potential bias or imprecision. Discuss both direction and magnitude of any potential bias | Page 21, lines 413-421 | Discussion, paragraph 5 |
| Interpretation | 20 | Give a cautious overall interpretation of results considering objectives, limitations, multiplicity of analyses, results from similar studies, and other relevant evidence | Pages 18-19, lines 339-364 | Discussion, paragraph 2 |
| Generalisability | 21 | Discuss the generalisability (external validity) of the study results | Page 21, lines 418-419 | Discussion, paragraph 5 |
| **Other information** | | | | |
| Funding | 22 | Give the source of funding and the role of the funders for the present study and, if applicable, for the original study on which the present article is based | Page 27, lines 604-609 | Notes |

*Give information separately for exposed and unexposed groups.

**Note:** An Explanation and Elaboration article discusses each checklist item and gives methodological background and published examples of transparent reporting. The STROBE checklist is best used in conjunction with this article (freely available on the Web sites of PLoS Medicine at http://www.plosmedicine.org/, Annals of Internal Medicine at http://www.annals.org/, and Epidemiology at http://www.epidem.com/). Information on the STROBE Initiative is available at http://www.strobe-statement.org.

**Supplementary Table 3. STROBE-MR checklist.**

| Item | Complete/location |
|---|---|
| 1. **Title and Abstract:** "Mendelian randomization" is named both in the title and the abstract | Title and abstract |
| **Introduction** | |
| 2. **Background:** Explain the scientific background and rationale for the reported study. Is causality between exposure and outcome plausible? Justify why MR is a helpful method to address the study question. | Introduction, paragraphs 1-3 |
| 3. **Objectives:** State specific objectives clearly, including pre-specified causal hypotheses (if any). | Introduction, paragraph 4 |
| **Methods** | |
| 4. **Study design and data sources:** Present key elements of study design early in the paper. Consider including a table listing sources of data for all phases of the study. For each data source contributing to the analysis, describe the following:<br>a) Describe the study design and the underlying population from which it was drawn. Describe also the setting, locations, and relevant dates, including periods of recruitment, exposure, follow-up, and data collection, if available.<br>b) Give the eligibility criteria, and the sources and methods of selection of participants.<br>c) Explain how the analyzed sample size was arrived at.<br>d) Describe measurement, quality and selection of genetic variants.<br>e) For each exposure, outcome and other relevant variables, describe methods of assessment and, in the case of diseases, the diagnostic criteria used.<br>f) Provide details of ethics committee approval and participant informed consent, if relevant. | a) Methods, section of "Participants and study design"<br>b) Methods, section of "Participants and study design"<br>c) Methods, section of "Participants and study design"; section of "Genetic associations with PUFAs"; section of "Genetic associations with COVID-19"<br>d) Methods, section of "Genetic associations with PUFAs"; section of "Genetic associations with COVID-19"<br>e) Methods, section of "Genetic associations with PUFAs"; section of "Genetic associations with COVID-19"<br>f) Methods, section of "Ethical considerations" |
| 5. **Assumptions:** Explicitly state assumptions for the main analysis (e.g. relevance, exclusion, independence, homogeneity) as well assumptions for any additional or sensitivity analysis. | Methods, section of "Genetic associations with PUFAs"; section of "Genetic associations with COVID-19"; section of "MR analyses" |
| 6. **Statistical methods main analysis**<br>Describe statistical methods and statistics used.<br>a) Describe how quantitative variables were handled in the analyses (i.e., scale, units, model).<br>b) Describe the process for identifying genetic variants and weights to be included in the analyses (i.e, independence and model). Consider a flow diagram. | a) Methods, section of "Genetic associations with PUFAs"; section of "Genetic associations with COVID-19" |

| | |
|---|---|
| c) Describe the MR estimator, e.g. two-stage least squares, Wald ratio, and related statistics. Detail the included covariates and, in case of two-sample MR, whether the same covariate set was used for adjustment in the two samples.<br>d) Explain how missing data were addressed.<br>e) If applicable, say how multiple testing was dealt with. | b) Methods, section of "Genetic associations with PUFAs"; section of "Genetic associations with COVID-19"<br>c) Methods, section of "MR analyses"<br>d) NA<br>e) Methods, section of "MR analyses" |
| **7. Assessment of assumptions: Describe any methods used to assess the assumptions or justify their validity.** | Methods, section of "MR analyses" |
| **8. Sensitivity analyses:** Describe any sensitivity analyses or additional analyses performed. | Methods, section of "MR analyses" |
| **9. Software and pre-registration**<br>a) Name statistical software and package(s), including version and settings used.<br>b) State whether the study protocol and details were pre-registered (as well as when and where). | a) Methods, section of section of "MR analyses"<br>b) Data availability |
| **Results** | |
| **10. Descriptive data**<br>a) Report the numbers of individuals at each stage of included studies and reasons for exclusion. Consider use of a flow-diagram.<br>b) Report summary statistics for phenotypic exposure(s), outcome(s) and other relevant variables (e.g. means, standard deviations, proportions).<br>c) If the data sources include meta-analyses of previous studies, provide the number of studies, their reported ancestry, if available, and assessments of heterogeneity across these studies. Consider using a supplementary table for each data source.<br>d) For two-sample Mendelian randomization:<br>i. Provide information on the similarity of the genetic variant-exposure associations between the exposure and outcome samples.<br>ii. Provide information on extent of sample overlap between the exposure and outcome data sources. | a) Results, section of "Bidirectional MR analyses"<br>b) Results, section of "Bidirectional MR analyses"; Supplementary Tables 4-5<br>c) Results, section of "Bidirectional MR analyses"<br>d) Results, section of "Bidirectional MR analyses" |
| **11. Main results**<br>a) Report the associations between genetic variant and exposure, and between genetic variant and outcome, preferably on an interpretable scale (e.g. comparing 25th and 75th percentile of allele count or genetic risk score, if individual-level data available).<br>b) Report causal effect estimate between exposure and outcome, and the measures of uncertainty from the MR analysis. Use an intuitive scale, such as odds ratio, or relative risk, per standard deviation difference. | a) Results, section of "Bidirectional MR analyses"<br>b) Results, section of "Bidirectional MR analyses"; Supplementary Tables 6-29<br>c) NA<br>d) Figures 2-3 |

| | |
|---|---|
| c) If relevant, consider translating estimates of relative risk into absolute risk for a meaningful time-period.<br>d) Consider any plots to visualize results (e.g. forest plot, scatterplot of associations between genetic variants and outcome versus between genetic variants and exposure). | |
| **12. Assessment of assumptions**<br>a) Assess the validity of the assumptions.<br>b) Report any additional statistics (e.g., assessments of heterogeneity, such as I2, Q statistic). | a) Results, section of "Bidirectional MR analyses"; Supplementary Tables 6-29<br>b) Results, section of "Bidirectional MR analyses"; Supplementary Tables 6-29 |
| **13. Sensitivity and additional analyses**<br>a) Use sensitivity analyses to assess the robustness of the main results to violations of the assumptions.<br>b) Report results from other sensitivity analyses (e.g., replication study with different dataset, analyses of subgroups, validation of instrument(s), simulations, etc.).<br>c) Report any assessment of direction of causality (e.g., bidirectional MR).<br>d) When relevant, report and compare with estimates from non-MR analyses.<br>e) Consider any additional plots to visualize results (e.g., leave-one-out analyses). | a) Results, section of "Bidirectional MR analyses"; Supplementary Tables 6-29<br>b) Results, section of "Bidirectional MR analyses"; Supplementary Tables 6-29<br>c) Results, section of "Bidirectional MR analyses"; Supplementary Tables 14-29<br>d) Results, section of "Observational analysis"; Table 2<br>e) Figures 2-3 |
| **Discussion** | |
| **14. Key results** | Discussion, paragraph 1 |
| **15. Limitations**<br>Discuss limitations of the study, taking into account the validity of the MR assumptions, other sources of potential bias, and imprecision. Discuss both direction and magnitude of any potential bias, and any efforts to address them. | Discussion, paragraph 5 |
| **16. Interpretations**<br>a) Give a cautious overall interpretation of results considering objectives and limitations. Compare with results from other relevant studies.<br>b) Discuss underlying biological mechanisms that could be modelled by using the genetic variants to assess the relationship between the exposure and the outcome.<br>c) Discuss whether the results have clinical or policy relevance, and whether interventions could have the same size effect. | a) Discussion, paragraphs 2-3<br>b) Discussion, paragraphs 2-3<br>c) Discussion, paragraph 2 |
| **17. Generalizability:** | Discussion, paragraph 5 |
| **18. Funding:** | Notes |

| | | |
|---|---|---|
| **19.** | **Data and data sharing:** | Data availability |
| **20.** | **Conflicts of Interest:** | Acknowledgments |

**Supplementary Table 4. Genetic instruments for plasma polyunsaturated fatty acids.**

| PUFA | SNP | Chr | Position (BP) | Nearest gene | Effect allele | Non effect allele | EAF | Beta | SE | P-value |
|---|---|---|---|---|---|---|---|---|---|---|
| ALA | rs174547 | 11 | 61570783 | *FADS1* | C | T | 0.33124 | 0.016 | 0.001 | $3.50\times10^{-641}$ |
| LA | rs10740118 | 10 | 65101207 | *JMJD1C* | C | G | 0.407725 | -0.248 | 0.043 | $8.10\times10^{-9}$ |
| LA | rs174547 | 11 | 61570783 | *FADS1* | C | T | 0.33124 | 1.474 | 0.042 | $5.00\times10^{-274}$ |
| LA | rs16966952 | 16 | 15135943 | *NTAN1* | A | G | 0.279871 | -0.351 | 0.044 | $1.20\times10^{-15}$ |
| GLA | rs174547 | 11 | 61570783 | *FADS1* | C | T | 0.33124 | -0.016 | 0.001 | $2.30\times10^{-72}$ |
| GLA | rs16966952 | 16 | 15135943 | *PDXDC1* | A | G | 0.279871 | -0.0061 | 0.001 | $5.00\times10^{-11}$ |
| DGLA | rs174547 | 11 | 61570783 | *FADS1* | C | T | 0.33124 | 0.36 | 0.01 | $2.60\times10^{-151}$ |
| DGLA | rs16966952 | 16 | 15135943 | *PDXDC1* | A | G | 0.279871 | -0.22 | 0.02 | $7.60\times10^{-15}$ |
| AA | rs174547 | 11 | 61570783 | *FADS1* | C | T | 0.33124 | -1.691 | 0.025 | $3\times10^{-971}$ |
| AA | rs16966952 | 16 | 15135943 | *NTAN1* | A | G | 0.279871 | -0.199 | 0.031 | $2.40\times10^{-10}$ |
| DPA-n3 | rs780094 | 2 | 27741237 | *GCKR* | T | C | 0.447679 | 0.017 | 0.003 | $9.04\times10^{-9}$ |
| DPA-n3 | rs3734398 | 6 | 10982973 | *ELOVL2* | C | T | 0.434361 | 0.04 | 0.003 | $9.60\times10^{-44}$ |
| DPA-n3 | rs174547 | 11 | 61570783 | *FADS1* | T | C | 0.66876 | 0.075 | 0.003 | $3.80\times10^{-154}$ |
| DHA | rs2236212 | 6 | 10995015 | *ELOVL2* | G | C | 0.569 | 0.113 | 0.014 | $1.30\times10^{-15}$ |

PUFA, polyunsaturated fatty acid; SNP, single nucleotide polymorphism; Chr, chromosome; EAF, Effect allele frequency; SE, standard error. For each PUFA, we selected SNPs that reached genome-wide significance level ($P < 5 \times 10^{-8}$) and were restricted by linkage disequilibrium (LD) clumping to ensure independence ($R^2 < 0.001$ within a 10 Mb window).

**Supplementary Table 5. Significant SNPs for red blood cell polyunsaturated fatty acids.**

| PUFA | SNP | Chr | Position (BP) | Nearest gene | Effect allele | Non effect allele | EAF | Beta | SE | P-value |
|---|---|---|---|---|---|---|---|---|---|---|
| ALA | rs174548 | 11 | 61571348 | *FADS1* | C | G | 0.69996 | -0.031079453 | 0.005544021 | $2.10\times10^{-8}$ |
| ALA | rs1535 | 11 | 61597972 | *FADS2* | A | G | 0.66595 | -0.030344391 | 0.005554751 | $4.70\times10^{-8}$ |
| ALA | rs174574 | 11 | 61600342 | *FADS2* | C | A | 0.63796 | -0.030961912 | 0.005666031 | $4.60\times10^{-8}$ |
| LA | rs174550 | 11 | 61571478 | *FADS1* | T | C | 0.67143 | -0.025923175 | 0.001983862 | $<1\times10^{-16}$ |
| LA | rs174547 | 11 | 61570783 | *FADS1* | T | C | 0.67136 | -0.025924923 | 0.001983583 | $<1\times10^{-16}$ |
| LA | rs174546 | 11 | 61569830 | *FADS1* | C | T | 0.67116 | -0.02592704 | 0.001984449 | $<1\times10^{-16}$ |
| LA | rs174545 | 11 | 61569306 | *FADS1* | C | G | 0.6711 | -0.025927951 | 0.001984711 | $<1\times10^{-16}$ |
| LA | rs174541 | 11 | 61565908 | | T | C | 0.65148 | -0.026187705 | 0.002005041 | $<1\times10^{-16}$ |
| LA | rs4246215 | 11 | 61564299 | *FEN1* | G | T | 0.65088 | -0.02618057 | 0.002007268 | $<1\times10^{-16}$ |
| LA | rs174549 | 11 | 61571382 | *FADS1* | G | A | 0.70775 | -0.026380702 | 0.002034896 | $<1\times10^{-16}$ |
| LA | rs174555 | 11 | 61579760 | *FADS1* | T | C | 0.70764 | -0.026513764 | 0.002047489 | $<1\times10^{-16}$ |
| LA | rs174556 | 11 | 61580635 | *FADS1* | C | T | 0.70764 | -0.026518518 | 0.002048022 | $<1\times10^{-16}$ |
| LA | rs174548 | 11 | 61571348 | *FADS1* | C | G | 0.69996 | -0.026169921 | 0.0020244 | $<1\times10^{-16}$ |
| LA | rs1535 | 11 | 61597972 | *FADS2* | A | G | 0.66595 | -0.02613899 | 0.002025075 | $<1\times10^{-16}$ |
| LA | rs174537 | 11 | 61552680 | *C11orf9* | G | T | 0.66488 | -0.025442966 | 0.00200945 | $<1\times10^{-16}$ |
| LA | rs174576 | 11 | 61603510 | *FADS2* | C | A | 0.65841 | -0.025850504 | 0.0020428 | $<1\times10^{-16}$ |
| LA | rs174535 | 11 | 61551356 | *C11orf9* | T | C | 0.65839 | -0.025365057 | 0.002007878 | $<1\times10^{-16}$ |
| LA | rs174536 | 11 | 61551927 | *C11orf9* | A | C | 0.66386 | -0.025322394 | 0.002009214 | $<1\times10^{-16}$ |
| LA | rs174577 | 11 | 61604814 | *FADS2* | C | A | 0.6524 | -0.025577242 | 0.002040915 | $<1\times10^{-16}$ |
| LA | rs174574 | 11 | 61600342 | *FADS2* | C | A | 0.63796 | -0.025861826 | 0.002072471 | $<1\times10^{-16}$ |
| LA | rs174578 | 11 | 61605499 | *FADS2* | T | A | 0.65154 | -0.025095132 | 0.002039835 | $<1\times10^{-16}$ |
| LA | rs102275 | 11 | 61557803 | *C11orf10* | T | C | 0.62686 | -0.02551658 | 0.002076206 | $<1\times10^{-16}$ |
| LA | rs174583 | 11 | 61609750 | *FADS2* | C | T | 0.65299 | -0.025052255 | 0.002052984 | $<1\times10^{-16}$ |
| LA | rs174538 | 11 | 61560081 | *C11orf10* | G | A | 0.69487 | -0.024581334 | 0.002023435 | $<1\times10^{-16}$ |
| LA | rs174528 | 11 | 61543499 | *C11orf9* | T | C | 0.61669 | -0.023670615 | 0.002054035 | $<1\times10^{-16}$ |
| LA | rs174601 | 11 | 61623140 | *FADS2* | C | T | 0.61373 | -0.024275521 | 0.002151782 | $<1\times10^{-16}$ |
| LA | rs108499 | 11 | 61547237 | *C11orf9* | C | T | 0.67142 | -0.021955949 | 0.002013526 | $<1\times10^{-16}$ |
| LA | rs174534 | 11 | 61549458 | *C11orf9* | A | G | 0.66316 | -0.020878982 | 0.001958043 | $<1\times10^{-16}$ |
| LA | rs174575 | 11 | 61602003 | *FADS2* | C | G | 0.7332 | -0.02085927 | 0.002270508 | $<1\times10^{-16}$ |
| LA | rs174593 | 11 | 61618831 | *FADS2* | T | C | 0.7208 | -0.019803494 | 0.002267427 | $<1\times10^{-16}$ |
| LA | rs174579 | 11 | 61605613 | *FADS2* | C | T | 0.78402 | -0.019710352 | 0.002352998 | $<1\times10^{-16}$ |
| LA | rs174591 | 11 | 61617676 | *FADS2* | T | A | 0.71832 | -0.019329175 | 0.002308087 | $<1\times10^{-16}$ |
| LA | rs968567 | 11 | 61595564 | *FADS2* | C | T | 0.83527 | -0.022516292 | 0.002717439 | $1.10\times10^{-16}$ |

| | | | | | | | | | | |
|---|---|---|---|---|---|---|---|---|---|---|
| LA | rs174585 | 11 | 61611694 | *FADS2* | G | A | 0.78304 | -0.019280656 | 0.002340284 | $2.20\times10^{-16}$ |
| LA | rs174448 | 11 | 61639573 | | A | G | 0.64303 | -0.01629102 | 0.002011707 | $5.60\times10^{-16}$ |
| LA | rs174570 | 11 | 61597212 | *FADS2* | C | T | 0.86422 | -0.023734742 | 0.002960526 | $1.10\times10^{-15}$ |
| LA | rs174589 | 11 | 61615803 | *FADS2* | C | G | 0.78765 | -0.018654832 | 0.002330472 | $1.20\times10^{-15}$ |
| LA | rs422249 | 11 | 61639488 | | C | T | 0.67098 | -0.016220448 | 0.002071517 | $4.90\times10^{-15}$ |
| LA | rs174605 | 11 | 61626921 | *FADS2* | G | T | 0.71151 | -0.016218161 | 0.002074222 | $5.30\times10^{-15}$ |
| LA | rs174597 | 11 | 61621040 | *FADS2* | G | C | 0.74735 | -0.018547979 | 0.002373894 | $5.60\times10^{-15}$ |
| LA | rs174611 | 11 | 61627881 | *FADS2* | T | C | 0.71039 | -0.01626599 | 0.002083075 | $5.80\times10^{-15}$ |
| LA | rs174449 | 11 | 61640379 | | A | G | 0.63271 | -0.015007949 | 0.001941676 | $1.10\times10^{-14}$ |
| LA | rs509360 | 11 | 61548559 | *C11orf9* | G | A | 0.649 | 0.019146856 | 0.002509696 | $2.40\times10^{-14}$ |
| LA | rs2269928 | 11 | 61537529 | *C11orf9* | T | G | 0.77978 | -0.020839115 | 0.002805667 | $1.10\times10^{-13}$ |
| LA | rs2727270 | 11 | 61603237 | *FADS2* | C | T | 0.89029 | -0.022241527 | 0.002996633 | $1.20\times10^{-13}$ |
| LA | rs2727271 | 11 | 61603358 | *FADS2* | A | T | 0.89 | -0.022240948 | 0.002998407 | $1.20\times10^{-13}$ |
| LA | rs174627 | 11 | 61637466 | | G | A | 0.835 | -0.019385566 | 0.002750477 | $1.80\times10^{-12}$ |
| LA | rs2524299 | 11 | 61604782 | *FADS2* | A | T | 0.88064 | -0.021077959 | 0.003019821 | $3.00\times10^{-12}$ |
| LA | rs174479 | 11 | 61678754 | *RAB3IL1* | C | G | 0.81614 | -0.019966658 | 0.002868303 | $3.40\times10^{-12}$ |
| LA | rs2845573 | 11 | 61601908 | *FADS2* | A | G | 0.92668 | -0.025076609 | 0.003623849 | $4.50\times10^{-12}$ |
| LA | rs2072114 | 11 | 61605215 | *FADS2* | A | G | 0.8814 | -0.020285664 | 0.002962778 | $7.50\times10^{-12}$ |
| LA | rs174455 | 11 | 61656117 | *FADS3* | G | A | 0.36538 | 0.013278008 | 0.001944147 | $8.50\times10^{-12}$ |
| LA | rs2851682 | 11 | 61616012 | *FADS2* | A | G | 0.92129 | -0.026018 | 0.003862008 | $1.60\times10^{-11}$ |
| LA | rs174616 | 11 | 61629122 | *FADS2* | G | A | 0.52756 | -0.012926856 | 0.001927302 | $2.00\times10^{-11}$ |
| LA | rs174532 | 11 | 61548874 | *C11orf9* | G | A | 0.72114 | 0.017458535 | 0.002616894 | $2.50\times10^{-11}$ |
| LA | rs174626 | 11 | 61637057 | | A | G | 0.52886 | -0.01304876 | 0.001959376 | $2.70\times10^{-11}$ |
| LA | rs174450 | 11 | 61641542 | *FADS3* | G | T | 0.47314 | 0.013061701 | 0.001993513 | $5.70\times10^{-11}$ |
| LA | rs198462 | 11 | 61524119 | *C11orf9* | G | A | 0.48644 | 0.012838099 | 0.001984777 | $9.90\times10^{-11}$ |
| LA | rs198476 | 11 | 61525730 | *C11orf9* | G | A | 0.47167 | 0.01298316 | 0.002038935 | $1.90\times10^{-10}$ |
| LA | rs198464 | 11 | 61521621 | | G | A | 0.48411 | 0.012173434 | 0.001915542 | $2.10\times10^{-10}$ |
| LA | rs412334 | 11 | 61560261 | *FEN1* | C | T | 0.85305 | 0.026964394 | 0.004252483 | $2.30\times10^{-10}$ |
| LA | rs174634 | 11 | 61647387 | *FADS3* | C | G | 0.72588 | -0.013223298 | 0.002151382 | $7.90\times10^{-10}$ |
| LA | rs174464 | 11 | 61657926 | *FADS3* | G | A | 0.72982 | -0.013261427 | 0.002186526 | $1.30\times10^{-9}$ |
| LA | rs1000778 | 11 | 61655305 | *FADS3* | G | A | 0.72904 | -0.012692579 | 0.00211719 | $2.00\times10^{-9}$ |
| LA | rs174456 | 11 | 61656182 | *FADS3* | A | C | 0.72918 | -0.012665509 | 0.002115463 | $2.10\times10^{-9}$ |
| LA | rs2526678 | 11 | 61623793 | *FADS2* | G | A | 0.93057 | -0.026337496 | 0.004428129 | $2.70\times10^{-9}$ |
| LA | rs149803 | 11 | 61539020 | *C11orf9* | C | G | 0.74636 | 0.015625177 | 0.002763177 | $1.60\times10^{-8}$ |
| LA | rs12580543 | 12 | 7100973 | *MBOAT5* | A | C | 0.93596 | -0.045842834 | 0.0039947 | $<1\times10^{-16}$ |
| LA | rs4394918 | 12 | 7110335 | *MBOAT5* | C | A | 0.93926 | -0.04537712 | 0.004016596 | $<1\times10^{-16}$ |

| | | | | | | | | | | |
|---|---|---|---|---|---|---|---|---|---|---|
| LA | rs16928105 | 12 | 7114196 | *MBOAT5* | C | T | 0.93917 | -0.045109814 | 0.00400763 | $<1\times10^{-16}$ |
| LA | rs1984564 | 12 | 7090193 | *MBOAT5* | A | G | 0.91021 | -0.036294608 | 0.003302183 | $<1\times10^{-16}$ |
| LA | rs7952839 | 12 | 7110571 | *MBOAT5* | G | A | 0.90923 | -0.036105147 | 0.003312813 | $<1\times10^{-16}$ |
| LA | rs12582990 | 12 | 7145853 | | T | C | 0.90538 | -0.037264689 | 0.003452779 | $<1\times10^{-16}$ |
| LA | rs16933023 | 12 | 7107348 | *MBOAT5* | C | G | 0.9166 | -0.037045099 | 0.003512866 | $<1\times10^{-16}$ |
| LA | rs12580233 | 12 | 7124522 | *MBOAT5* | T | C | 0.91542 | -0.036368727 | 0.003485457 | $<1\times10^{-16}$ |
| LA | rs16933011 | 12 | 7073805 | | G | T | 0.91316 | -0.036528813 | 0.003541195 | $<1\times10^{-16}$ |
| LA | rs3764031 | 12 | 7091918 | *MBOAT5* | C | T | 0.91377 | -0.035061406 | 0.003409011 | $<1\times10^{-16}$ |
| LA | rs7311050 | 12 | 7120836 | *MBOAT5* | C | T | 0.91388 | -0.03495796 | 0.003409288 | $<1\times10^{-16}$ |
| LA | rs2110073 | 12 | 7075882 | *PHB2* | C | T | 0.89587 | -0.031198306 | 0.003100397 | $<1\times10^{-16}$ |
| LA | rs7962738 | 12 | 7166672 | *C1S* | T | A | 0.94463 | -0.040285903 | 0.004311937 | $<1\times10^{-16}$ |
| LA | rs12579775 | 12 | 7085171 | | G | A | 0.90521 | -0.030763165 | 0.003293403 | $<1\times10^{-16}$ |
| LA | rs759052 | 12 | 7069620 | *PTPN6* | C | T | 0.8603 | -0.029867787 | 0.003337284 | $<1\times10^{-16}$ |
| LA | rs11064497 | 12 | 7169661 | *C1S* | C | T | 0.86151 | -0.017528017 | 0.002841673 | $6.90\times10^{-10}$ |
| LA | rs12368181 | 12 | 7181105 | | A | G | 0.86823 | -0.017538707 | 0.002862766 | $9.00\times10^{-10}$ |
| LA | rs12366520 | 12 | 7181162 | | C | A | 0.8682 | -0.017532536 | 0.002863014 | $9.10\times10^{-10}$ |
| LA | rs12146727 | 12 | 7170336 | *C1S* | G | A | 0.86979 | -0.017143651 | 0.002814623 | $1.10\times10^{-9}$ |
| LA | rs16933078 | 12 | 7171338 | *C1S* | T | A | 0.86831 | -0.017158756 | 0.002818129 | $1.10\times10^{-9}$ |
| LA | rs16933084 | 12 | 7172084 | *C1S* | A | T | 0.86906 | -0.017078607 | 0.002814758 | $1.30\times10^{-9}$ |
| LA | rs11838267 | 12 | 7175872 | *C1S* | T | C | 0.86788 | -0.017089309 | 0.002813916 | $1.30\times10^{-9}$ |
| LA | rs12368783 | 12 | 7179400 | | G | T | 0.86858 | -0.017239953 | 0.002841371 | $1.30\times10^{-9}$ |
| LA | rs7962629 | 12 | 7166770 | *C1S* | A | G | 0.86456 | -0.017224081 | 0.002842693 | $1.40\times10^{-9}$ |
| LA | rs11064501 | 12 | 7179822 | | A | C | 0.86691 | -0.017250584 | 0.002845788 | $1.30\times10^{-9}$ |
| LA | rs10849546 | 12 | 7176204 | *C1S* | G | A | 0.86876 | -0.01702788 | 0.002813537 | $1.40\times10^{-9}$ |
| LA | rs12371227 | 12 | 7176978 | *C1S* | C | T | 0.86834 | -0.017016845 | 0.002810648 | $1.40\times10^{-9}$ |
| LA | rs7183 | 12 | 7178019 | *C1S* | G | T | 0.86851 | -0.017045684 | 0.002816664 | $1.40\times10^{-9}$ |
| LA | rs11064498 | 12 | 7171507 | *C1S* | A | G | 0.85324 | -0.016734174 | 0.002842967 | $4.00\times10^{-9}$ |
| LA | rs3919533 | 12 | 7162801 | | T | C | 0.83695 | -0.015492514 | 0.002653374 | $5.30\times10^{-9}$ |
| LA | rs7311672 | 12 | 7165114 | | G | A | 0.83872 | -0.014988034 | 0.002629557 | $1.20\times10^{-8}$ |
| GLA | rs174548 | 11 | 61571348 | *FADS1* | C | G | 0.69996 | 0.056093204 | 0.008052263 | $3.30\times10^{-12}$ |
| GLA | rs174549 | 11 | 61571382 | *FADS1* | G | A | 0.70775 | 0.05399605 | 0.008102706 | $2.70\times10^{-11}$ |
| GLA | rs174555 | 11 | 61579760 | *FADS1* | T | C | 0.70764 | 0.054235078 | 0.008152221 | $2.90\times10^{-11}$ |
| GLA | rs174556 | 11 | 61580635 | *FADS1* | C | T | 0.70764 | 0.054246393 | 0.008154209 | $2.90\times10^{-11}$ |
| GLA | rs4246215 | 11 | 61564299 | *FEN1* | G | T | 0.65088 | 0.051586938 | 0.008006419 | $1.20\times10^{-11}$ |
| GLA | rs174541 | 11 | 61565908 | | T | C | 0.65148 | 0.051462442 | 0.007998025 | $1.20\times10^{-10}$ |
| GLA | rs174535 | 11 | 61551356 | *C11orf9* | T | C | 0.65839 | 0.051333021 | 0.007993626 | $1.30\times10^{-10}$ |

| | | | | | | | | | | |
|---|---|---|---|---|---|---|---|---|---|---|
| GLA | rs102275 | 11 | 61557803 | *C11orf10* | T | C | 0.62686 | 0.052391657 | 0.008240345 | $2.00\times10^{-10}$ |
| GLA | rs174574 | 11 | 61600342 | *FADS2* | C | A | 0.63796 | 0.05204465 | 0.00824537 | $2.80\times10^{-10}$ |
| GLA | rs1535 | 11 | 61597972 | *FADS2* | A | G | 0.66595 | 0.050795864 | 0.008081966 | $3.30\times10^{-10}$ |
| GLA | rs174577 | 11 | 61604814 | *FADS2* | C | A | 0.6524 | 0.050837053 | 0.008124732 | $3.90\times10^{-10}$ |
| GLA | rs174578 | 11 | 61605499 | *FADS2* | T | A | 0.65154 | 0.050440978 | 0.008113275 | $5.10\times10^{-10}$ |
| GLA | rs174536 | 11 | 61551927 | *C11orf9* | A | C | 0.66386 | 0.049696809 | 0.008001506 | $5.30\times10^{-10}$ |
| GLA | rs174537 | 11 | 61552680 | *C11orf9* | G | T | 0.66488 | 0.049704656 | 0.008006979 | $5.40\times10^{-10}$ |
| GLA | rs174550 | 11 | 61571478 | *FADS1* | T | C | 0.67143 | 0.049090576 | 0.007921297 | $5.70\times10^{-10}$ |
| GLA | rs174545 | 11 | 61569306 | *FADS1* | C | G | 0.6711 | 0.049037087 | 0.007925609 | $6.10\times10^{-10}$ |
| GLA | rs174546 | 11 | 61569830 | *FADS1* | C | T | 0.67116 | 0.049019012 | 0.007924646 | $6.20\times10^{-10}$ |
| GLA | rs174547 | 11 | 61570783 | *FADS1* | T | C | 0.67136 | 0.048973853 | 0.007921437 | $6.30\times10^{-10}$ |
| GLA | rs174576 | 11 | 61603510 | *FADS2* | C | A | 0.65841 | 0.050172662 | 0.008139624 | $7.10\times10^{-10}$ |
| GLA | rs174583 | 11 | 61609750 | *FADS2* | C | T | 0.65299 | 0.049850145 | 0.00816299 | $1.00\times10^{-9}$ |
| GLA | rs174528 | 11 | 61543499 | *C11orf9* | T | C | 0.61669 | 0.049271281 | 0.008126065 | $1.30\times10^{-9}$ |
| DGLA | rs174555 | 11 | 61579760 | *FADS1* | T | C | 0.70764 | -0.093383417 | 0.002276368 | $<1\times10^{-16}$ |
| DGLA | rs174556 | 11 | 61580635 | *FADS1* | C | T | 0.70764 | -0.093407652 | 0.002276937 | $<1\times10^{-16}$ |
| DGLA | rs174549 | 11 | 61571382 | *FADS1* | G | A | 0.70775 | -0.092786816 | 0.002263023 | $<1\times10^{-16}$ |
| DGLA | rs174548 | 11 | 61571348 | *FADS1* | C | G | 0.69996 | -0.092279425 | 0.002252042 | $<1\times10^{-16}$ |
| DGLA | rs174538 | 11 | 61560081 | *C11orf10* | G | A | 0.69487 | -0.089612614 | 0.002277243 | $<1\times10^{-16}$ |
| DGLA | rs174545 | 11 | 61569306 | *FADS1* | C | G | 0.6711 | -0.087925132 | 0.002250125 | $<1\times10^{-16}$ |
| DGLA | rs174550 | 11 | 61571478 | *FADS1* | T | C | 0.67143 | -0.087877368 | 0.002249418 | $<1\times10^{-16}$ |
| DGLA | rs174546 | 11 | 61569830 | *FADS1* | C | T | 0.67116 | -0.087912789 | 0.002249869 | $<1\times10^{-16}$ |
| DGLA | rs174547 | 11 | 61570783 | *FADS1* | T | C | 0.67136 | -0.087868043 | 0.002249103 | $<1\times10^{-16}$ |
| DGLA | rs1535 | 11 | 61597972 | *FADS2* | A | G | 0.66595 | -0.089394544 | 0.002298171 | $<1\times10^{-16}$ |
| DGLA | rs174537 | 11 | 61552680 | *C11orf9* | G | T | 0.66488 | -0.088477735 | 0.0022804 | $<1\times10^{-16}$ |
| DGLA | rs174536 | 11 | 61551927 | *C11orf9* | A | C | 0.66386 | -0.088243053 | 0.002282768 | $<1\times10^{-16}$ |
| DGLA | rs174535 | 11 | 61551356 | *C11orf9* | T | C | 0.65839 | -0.08813322 | 0.00228418 | $<1\times10^{-16}$ |
| DGLA | rs174541 | 11 | 61565908 | | T | C | 0.65148 | -0.08718654 | 0.002300447 | $<1\times10^{-16}$ |
| DGLA | rs4246215 | 11 | 61564299 | *FEN1* | G | T | 0.65088 | -0.087265838 | 0.002302899 | $<1\times10^{-16}$ |
| DGLA | rs174574 | 11 | 61600342 | *FADS2* | C | A | 0.63796 | -0.089774642 | 0.002372438 | $<1\times10^{-16}$ |
| DGLA | rs174577 | 11 | 61604814 | *FADS2* | C | A | 0.6524 | -0.088450038 | 0.002337515 | $<1\times10^{-16}$ |
| DGLA | rs174576 | 11 | 61603510 | *FADS2* | C | A | 0.65841 | -0.088521769 | 0.002340525 | $<1\times10^{-16}$ |
| DGLA | rs102275 | 11 | 61557803 | *C11orf10* | T | C | 0.62686 | -0.089486222 | 0.002382655 | $<1\times10^{-16}$ |
| DGLA | rs174578 | 11 | 61605499 | *FADS2* | T | A | 0.65154 | -0.08766831 | 0.002343702 | $<1\times10^{-16}$ |
| DGLA | rs174583 | 11 | 61609750 | *FADS2* | C | T | 0.65299 | -0.087686767 | 0.002364368 | $<1\times10^{-16}$ |
| DGLA | rs108499 | 11 | 61547237 | *C11orf9* | C | T | 0.67142 | -0.083095987 | 0.002342602 | $<1\times10^{-16}$ |

| | | | | | | | | | | |
|---|---|---|---|---|---|---|---|---|---|---|
| DGLA | rs174528 | 11 | 61543499 | *C11orf9* | T | C | 0.61669 | -0.084062886 | 0.002413168 | $<1\times10^{-16}$ |
| DGLA | rs174534 | 11 | 61549458 | *C11orf9* | A | G | 0.66316 | -0.079069068 | 0.002300709 | $<1\times10^{-16}$ |
| DGLA | rs968567 | 11 | 61595564 | *FADS2* | C | T | 0.83527 | -0.108510323 | 0.003158099 | $<1\times10^{-16}$ |
| DGLA | rs174601 | 11 | 61623140 | *FADS2* | C | T | 0.61373 | -0.084828533 | 0.002568194 | $<1\times10^{-16}$ |
| DGLA | rs174579 | 11 | 61605613 | *FADS2* | C | T | 0.78402 | -0.08787706 | 0.00282469 | $<1\times10^{-16}$ |
| DGLA | rs174585 | 11 | 61611694 | *FADS2* | G | A | 0.78304 | -0.086669437 | 0.002816734 | $<1\times10^{-16}$ |
| DGLA | rs174575 | 11 | 61602003 | *FADS2* | C | G | 0.7332 | -0.083882013 | 0.002750669 | $<1\times10^{-16}$ |
| DGLA | rs174589 | 11 | 61615803 | *FADS2* | C | G | 0.78765 | -0.084379029 | 0.002825277 | $<1\times10^{-16}$ |
| DGLA | rs174591 | 11 | 61617676 | *FADS2* | T | A | 0.71832 | -0.080976985 | 0.002834895 | $<1\times10^{-16}$ |
| DGLA | rs174593 | 11 | 61618831 | *FADS2* | T | C | 0.7208 | -0.077790899 | 0.002813594 | $<1\times10^{-16}$ |
| DGLA | rs174597 | 11 | 61621040 | *FADS2* | G | C | 0.74735 | -0.079166265 | 0.002959683 | $<1\times10^{-16}$ |
| DGLA | rs174627 | 11 | 61637466 | | G | A | 0.835 | -0.081359634 | 0.003515982 | $<1\times10^{-16}$ |
| DGLA | rs2269928 | 11 | 61537529 | *C11orf9* | T | G | 0.77978 | -0.080790453 | 0.003623491 | $<1\times10^{-16}$ |
| DGLA | rs509360 | 11 | 61548559 | *C11orf9* | G | A | 0.649 | 0.070454333 | 0.003264146 | $<1\times10^{-16}$ |
| DGLA | rs174611 | 11 | 61627881 | *FADS2* | T | C | 0.71039 | -0.057047069 | 0.002721521 | $<1\times10^{-16}$ |
| DGLA | rs174605 | 11 | 61626921 | *FADS2* | G | T | 0.71151 | -0.056782086 | 0.002710525 | $<1\times10^{-16}$ |
| DGLA | rs174448 | 11 | 61639573 | | A | G | 0.64303 | -0.052877561 | 0.002655524 | $<1\times10^{-16}$ |
| DGLA | rs422249 | 11 | 61639488 | | C | T | 0.67098 | -0.053451571 | 0.00273317 | $<1\times10^{-16}$ |
| DGLA | rs174449 | 11 | 61640379 | | A | G | 0.63271 | -0.050106436 | 0.002567625 | $<1\times10^{-16}$ |
| DGLA | rs174469 | 11 | 61667443 | *RAB3IL1* | C | T | 0.82555 | -0.067713817 | 0.00367945 | $<1\times10^{-16}$ |
| DGLA | rs174479 | 11 | 61678754 | *RAB3IL1* | C | G | 0.81614 | -0.06773423 | 0.003816048 | $<1\times10^{-16}$ |
| DGLA | rs174634 | 11 | 61647387 | *FADS3* | C | G | 0.72588 | -0.050487213 | 0.00286443 | $<1\times10^{-16}$ |
| DGLA | rs526126 | 11 | 61624885 | *FADS2* | C | G | 0.76593 | -0.072444969 | 0.004129038 | $<1\times10^{-16}$ |
| DGLA | rs174464 | 11 | 61657926 | *FADS3* | G | A | 0.72982 | -0.04998859 | 0.002920249 | $<1\times10^{-16}$ |
| DGLA | rs1000778 | 11 | 61655305 | *FADS3* | G | A | 0.72904 | -0.048000579 | 0.002829881 | $<1\times10^{-16}$ |
| DGLA | rs174456 | 11 | 61656182 | *FADS3* | A | C | 0.72918 | -0.047889577 | 0.002828034 | $<1\times10^{-16}$ |
| DGLA | rs174532 | 11 | 61548874 | *C11orf9* | G | A | 0.72114 | 0.058842315 | 0.003511758 | $<1\times10^{-16}$ |
| DGLA | rs174455 | 11 | 61656117 | *FADS3* | G | A | 0.36538 | 0.042120936 | 0.002625943 | $<1\times10^{-16}$ |
| DGLA | rs412334 | 11 | 61560261 | *FEN1* | C | T | 0.85305 | 0.084373277 | 0.005760646 | $<1\times10^{-16}$ |
| DGLA | rs174616 | 11 | 61629122 | *FADS2* | G | A | 0.52756 | -0.038302878 | 0.002618867 | $<1\times10^{-16}$ |
| DGLA | rs174626 | 11 | 61637057 | | A | G | 0.52886 | -0.038773801 | 0.002662012 | $<1\times10^{-16}$ |
| DGLA | rs174450 | 11 | 61641542 | *FADS3* | G | T | 0.47314 | 0.038438145 | 0.002713908 | $<1\times10^{-16}$ |
| DGLA | rs149803 | 11 | 61539020 | *C11orf9* | C | G | 0.74636 | 0.051444004 | 0.003767124 | $<1\times10^{-16}$ |
| DGLA | rs174570 | 11 | 61597212 | *FADS2* | C | T | 0.86422 | -0.054262313 | 0.004080799 | $<1\times10^{-16}$ |
| DGLA | rs198462 | 11 | 61524119 | *C11orf9* | G | A | 0.48644 | 0.032261333 | 0.00275405 | $<1\times10^{-16}$ |
| DGLA | rs198464 | 11 | 61521621 | | G | A | 0.48411 | 0.030068552 | 0.002661756 | $<1\times10^{-16}$ |

| | | | | | | | | | | |
|---|---|---|---|---|---|---|---|---|---|---|
| DGLA | rs7102974 | 11 | 61560035 | *C11orf10* | C | T | 0.97886 | -0.125814384 | 0.011223658 | $<1\times10^{-16}$ |
| DGLA | rs198476 | 11 | 61525730 | *C11orf9* | G | A | 0.47167 | 0.031434935 | 0.002835967 | $<1\times10^{-16}$ |
| DGLA | rs498793 | 11 | 61624705 | *FADS2* | C | T | 0.60008 | 0.045940115 | 0.004146446 | $<1\times10^{-16}$ |
| DGLA | rs174468 | 11 | 61663691 | | G | A | 0.60365 | 0.034164486 | 0.003117167 | $<1\times10^{-16}$ |
| DGLA | rs2845573 | 11 | 61601908 | *FADS2* | A | G | 0.92668 | -0.054916984 | 0.005041632 | $<1\times10^{-16}$ |
| DGLA | rs2851682 | 11 | 61616012 | *FADS2* | A | G | 0.92129 | -0.057949708 | 0.00537644 | $<1\times10^{-16}$ |
| DGLA | rs198473 | 11 | 61526556 | *C11orf9* | A | G | 0.74785 | -0.035487193 | 0.003376086 | $<1\times10^{-16}$ |
| DGLA | rs174476 | 11 | 61674118 | *RAB3IL1* | C | T | 0.60208 | 0.033121705 | 0.00318768 | $<1\times10^{-16}$ |
| DGLA | rs174478 | 11 | 61678576 | *RAB3IL1* | T | G | 0.60099 | 0.033154421 | 0.003200503 | $<1\times10^{-16}$ |
| DGLA | rs174602 | 11 | 61624414 | *FADS2* | T | C | 0.76774 | -0.045141254 | 0.004359976 | $<1\times10^{-16}$ |
| DGLA | rs666870 | 11 | 61677479 | *RAB3IL1* | G | A | 0.6013 | 0.033122844 | 0.003199448 | $<1\times10^{-16}$ |
| DGLA | rs198475 | 11 | 61526071 | *C11orf9* | C | T | 0.75215 | -0.034406845 | 0.003339929 | $<1\times10^{-16}$ |
| DGLA | rs13966 | 11 | 61664992 | *RAB3IL1* | C | T | 0.54676 | 0.031902864 | 0.003191463 | $<1\times10^{-16}$ |
| DGLA | rs2235093 | 11 | 61665122 | *RAB3IL1* | G | A | 0.54967 | 0.031732481 | 0.003194219 | $<1\times10^{-16}$ |
| DGLA | rs579383 | 11 | 61536583 | *C11orf9* | A | G | 0.57529 | 0.030046342 | 0.00304175 | $<1\times10^{-16}$ |
| DGLA | rs2727270 | 11 | 61603237 | *FADS2* | C | T | 0.89029 | -0.041144884 | 0.004187134 | $<1\times10^{-16}$ |
| DGLA | rs2727271 | 11 | 61603358 | *FADS2* | A | T | 0.89 | -0.041055738 | 0.004190331 | $<1\times10^{-16}$ |
| DGLA | rs650436 | 11 | 61536430 | *C11orf9* | C | T | 0.57937 | 0.029706454 | 0.003050024 | $<1\times10^{-16}$ |
| DGLA | rs2526678 | 11 | 61623793 | *FADS2* | G | A | 0.93057 | -0.059961422 | 0.006165896 | $<1\times10^{-16}$ |
| DGLA | rs198446 | 11 | 61503381 | *DAGLA* | G | A | 0.74811 | -0.03169164 | 0.003313523 | $<1\times10^{-16}$ |
| DGLA | rs2072114 | 11 | 61605215 | *FADS2* | A | G | 0.8814 | -0.039457728 | 0.004132432 | $<1\times10^{-16}$ |
| DGLA | rs1692120 | 11 | 61417472 | | G | A | 0.56219 | 0.03328641 | 0.003663514 | $<1\times10^{-16}$ |
| DGLA | rs174472 | 11 | 61671956 | *RAB3IL1* | G | A | 0.45428 | 0.0295229 | 0.00325842 | $<1\times10^{-16}$ |
| DGLA | rs740006 | 11 | 61557868 | *C11orf10* | T | C | 0.91448 | 0.059024656 | 0.006536418 | $<1\times10^{-16}$ |
| DGLA | rs1800009 | 11 | 61730234 | *BEST1* | C | T | 0.3478 | 0.026452212 | 0.002951664 | $<1\times10^{-16}$ |
| DGLA | rs2524299 | 11 | 61604782 | *FADS2* | A | T | 0.88064 | -0.037770077 | 0.004228786 | $<1\times10^{-16}$ |
| DGLA | rs198418 | 11 | 61496272 | *DAGLA* | A | C | 0.73879 | -0.028012495 | 0.003145658 | $<1\times10^{-16}$ |
| DGLA | rs198425 | 11 | 61491431 | *DAGLA* | A | T | 0.7406 | -0.027561243 | 0.003129851 | $<1\times10^{-16}$ |
| DGLA | rs2453710 | 11 | 61406542 | *RPLP0P2* | A | G | 0.58218 | 0.034261295 | 0.003928726 | $<1\times10^{-16}$ |
| DGLA | rs3758977 | 11 | 61737244 | *LOC399900* | G | T | 0.3436 | 0.025007725 | 0.002869771 | $<1\times10^{-16}$ |
| DGLA | rs81658 | 11 | 61487944 | *DAGLA* | G | A | 0.74192 | -0.026810422 | 0.003098609 | $<1\times10^{-16}$ |
| DGLA | rs198430 | 11 | 61487690 | *DAGLA* | G | A | 0.74192 | -0.026799632 | 0.003098162 | $<1\times10^{-16}$ |
| DGLA | rs17633020 | 11 | 61744881 | | G | A | 0.76205 | -0.026458923 | 0.003117735 | $<1\times10^{-16}$ |
| DGLA | rs17185574 | 11 | 61745694 | | T | C | 0.76159 | -0.026226638 | 0.003098671 | $<1\times10^{-16}$ |
| DGLA | rs198432 | 11 | 61484981 | *DAGLA* | C | A | 0.73843 | -0.026339822 | 0.003116762 | $<1\times10^{-16}$ |

| | | | | | | | | | | |
|---|---|---|---|---|---|---|---|---|---|---|
| DGLA | rs1791785 | 11 | 61442813 | | C | T | 0.7418 | -0.029047122 | 0.003440297 | $<1\times10^{-16}$ |
| DGLA | rs2028062 | 11 | 61745953 | | A | G | 0.34067 | 0.023546253 | 0.002808945 | $<1\times10^{-16}$ |
| DGLA | rs10792320 | 11 | 61746291 | | C | A | 0.34095 | 0.023443942 | 0.002812546 | $1.10\times10^{-16}$ |
| DGLA | rs198453 | 11 | 61464550 | *DAGLA* | C | T | 0.73648 | -0.027339291 | 0.003359827 | $4.40\times10^{-16}$ |
| DGLA | rs472031 | 11 | 61638420 | | G | A | 0.90451 | 0.039188093 | 0.004829156 | $4.40\times10^{-16}$ |
| DGLA | rs482548 | 11 | 61633182 | *FADS2* | C | T | 0.90592 | 0.03821008 | 0.004845138 | $3.10\times10^{-15}$ |
| DGLA | rs2521568 | 11 | 61700933 | *RAB3IL1* | G | C | 0.92845 | -0.039959378 | 0.005208729 | $1.70\times10^{-14}$ |
| DGLA | rs3825036 | 11 | 61516476 | | G | A | 0.81744 | 0.030058234 | 0.003918162 | $1.70\times10^{-14}$ |
| DGLA | rs2727266 | 11 | 61704334 | *RAB3IL1* | A | G | 0.92849 | -0.039999915 | 0.005295094 | $4.20\times10^{-14}$ |
| DGLA | rs2521572 | 11 | 61711475 | *RAB3IL1* | G | T | 0.96168 | -0.050851342 | 0.006866003 | $1.30\times10^{-13}$ |
| DGLA | rs2240287 | 11 | 61505583 | *DAGLA* | G | A | 0.82919 | 0.028707876 | 0.003943812 | $3.40\times10^{-13}$ |
| DGLA | rs2238001 | 11 | 61524507 | *C11orf9* | T | C | 0.8425 | 0.031498283 | 0.004338058 | $3.80\times10^{-13}$ |
| DGLA | rs879486 | 11 | 61475233 | *DAGLA* | C | T | 0.6649 | -0.022341409 | 0.003125355 | $8.80\times10^{-13}$ |
| DGLA | rs9735635 | 11 | 61490880 | *DAGLA* | C | A | 0.83017 | 0.027813027 | 0.003894957 | $9.30\times10^{-13}$ |
| DGLA | rs12274157 | 11 | 61477647 | *DAGLA* | A | C | 0.82525 | 0.027112706 | 0.003823371 | $1.30\times10^{-12}$ |
| DGLA | rs2727261 | 11 | 61712131 | *RAB3IL1* | C | T | 0.90126 | -0.037934632 | 0.005412819 | $2.40\times10^{-12}$ |
| DGLA | rs198435 | 11 | 61483324 | *DAGLA* | G | C | 0.84649 | 0.028753775 | 0.004101929 | $2.40\times10^{-12}$ |
| DGLA | rs4963243 | 11 | 61494327 | *DAGLA* | G | A | 0.84942 | 0.029374573 | 0.004225089 | $3.60\times10^{-12}$ |
| DGLA | rs4423188 | 11 | 61766418 | | T | A | 0.72099 | -0.021455644 | 0.003103223 | $4.70\times10^{-12}$ |
| DGLA | rs10736716 | 11 | 61765133 | | G | C | 0.63697 | -0.019460274 | 0.00292327 | $2.80\times10^{-11}$ |
| DGLA | rs569258 | 11 | 61520668 | *MYRF* | T | C | 0.65011 | 0.020504387 | 0.003093989 | $3.40\times10^{-11}$ |
| DGLA | rs174552 | 11 | 61574999 | *FADS1* | C | T | 0.99468 | -0.200839135 | 0.030353866 | $3.70\times10^{-11}$ |
| DGLA | rs11827215 | 11 | 61458595 | *DAGLA* | G | A | 0.83972 | 0.024691446 | 0.003732647 | $3.70\times10^{-11}$ |
| DGLA | rs198746 | 11 | 61376970 | | G | A | 0.81159 | 0.057831365 | 0.008769685 | $4.30\times10^{-11}$ |
| DGLA | rs4963308 | 11 | 61456426 | *DAGLA* | G | A | 0.83996 | 0.024468091 | 0.003719449 | $4.80\times10^{-11}$ |
| DGLA | rs883724 | 11 | 61457857 | *DAGLA* | C | T | 0.84031 | 0.024428044 | 0.003715425 | $4.90\times10^{-11}$ |
| DGLA | rs1109748 | 11 | 61722645 | *BEST1* | C | A | 0.92539 | -0.036466281 | 0.005670758 | $1.30\times10^{-10}$ |
| DGLA | rs17762402 | 11 | 61553201 | *C11orf9* | G | A | 0.94147 | 0.046143991 | 0.007250444 | $2.00\times10^{-10}$ |
| DGLA | rs4313591 | 11 | 61766569 | | T | C | 0.62638 | -0.018246733 | 0.002906377 | $3.40\times10^{-10}$ |
| DGLA | rs4963441 | 11 | 61768639 | | C | G | 0.62575 | -0.01808825 | 0.002903362 | $4.70\times10^{-10}$ |
| DGLA | rs6591657 | 11 | 61434532 | | G | T | 0.88788 | 0.028180673 | 0.004523613 | $4.70\times10^{-10}$ |
| DGLA | rs7937198 | 11 | 61768973 | | A | G | 0.62364 | -0.018037627 | 0.002900861 | $5.00\times10^{-10}$ |
| DGLA | rs4963442 | 11 | 61768706 | | T | C | 0.62445 | -0.018054672 | 0.002905273 | $5.20\times10^{-10}$ |
| DGLA | rs7925523 | 11 | 61442492 | | G | A | 0.83567 | 0.023437215 | 0.003773306 | $5.30\times10^{-10}$ |
| DGLA | rs198745 | 11 | 61376680 | | T | C | 0.8253 | 0.055397405 | 0.008959375 | $6.30\times10^{-10}$ |
| DGLA | rs198428 | 11 | 61489705 | *DAGLA* | T | A | 0.61775 | 0.018459334 | 0.003019546 | $9.80\times10^{-10}$ |

| | | | | | | | | | | |
|---|---|---|---|---|---|---|---|---|---|---|
| DGLA | rs198426 | 11 | 61490486 | *DAGLA* | C | T | 0.67872 | 0.019104342 | 0.003140967 | $1.20\times10^{-9}$ |
| DGLA | rs11230749 | 11 | 61362978 | *LOC101927495* | C | T | 0.87613 | -0.069231534 | 0.011822452 | $4.70\times10^{-9}$ |
| AA | rs2581624 | 3 | 142633869 | | G | C | 0.79336 | 0.009754995 | 0.001589181 | $8.30\times10^{-10}$ |
| AA | rs2248811 | 3 | 142606942 | *PCOLCE2* | G | C | 0.78715 | 0.008693928 | 0.001593601 | $4.90\times10^{-9}$ |
| AA | rs174545 | 11 | 61569306 | *FADS1* | C | G | 0.6711 | 0.024581721 | 0.001250655 | $<1\times10^{-16}$ |
| AA | rs174546 | 11 | 61569830 | *FADS1* | C | T | 0.67116 | 0.024577777 | 0.001250506 | $<1\times10^{-16}$ |
| AA | rs174547 | 11 | 61570783 | *FADS1* | T | C | 0.67136 | 0.024564543 | 0.001250019 | $<1\times10^{-16}$ |
| AA | rs174550 | 11 | 61571478 | *FADS1* | T | C | 0.67143 | 0.024550948 | 0.001250319 | $<1\times10^{-16}$ |
| AA | rs174537 | 11 | 61552680 | *C11orf9* | G | T | 0.66488 | 0.024690364 | 0.001264599 | $<1\times10^{-16}$ |
| AA | rs1535 | 11 | 61597972 | *FADS2* | A | G | 0.66595 | 0.024866869 | 0.00127604 | $<1\times10^{-16}$ |
| AA | rs174536 | 11 | 61551927 | *C11orf9* | A | C | 0.66386 | 0.024631536 | 0.001264481 | $<1\times10^{-16}$ |
| AA | rs174535 | 11 | 61551356 | *C11orf9* | T | C | 0.65839 | 0.024623865 | 0.001264411 | $<1\times10^{-16}$ |
| AA | rs4246215 | 11 | 61564299 | *FEN1* | G | T | 0.65088 | 0.024433841 | 0.001268673 | $<1\times10^{-16}$ |
| AA | rs174541 | 11 | 61565908 | | T | C | 0.65148 | 0.024407299 | 0.001267406 | $<1\times10^{-16}$ |
| AA | rs102275 | 11 | 61557803 | *C11orf10* | T | C | 0.62686 | 0.024886779 | 0.001309693 | $<1\times10^{-16}$ |
| AA | rs174576 | 11 | 61603510 | *FADS2* | C | A | 0.65841 | 0.024507775 | 0.001290441 | $<1\times10^{-16}$ |
| AA | rs174574 | 11 | 61600342 | *FADS2* | C | A | 0.63796 | 0.024829465 | 0.001307902 | $<1\times10^{-16}$ |
| AA | rs174577 | 11 | 61604814 | *FADS2* | C | A | 0.6524 | 0.024440279 | 0.001288954 | $<1\times10^{-16}$ |
| AA | rs174578 | 11 | 61605499 | *FADS2* | T | A | 0.65154 | 0.024219492 | 0.001288197 | $<1\times10^{-16}$ |
| AA | rs174583 | 11 | 61609750 | *FADS2* | C | T | 0.65299 | 0.024199448 | 0.001296296 | $<1\times10^{-16}$ |
| AA | rs174555 | 11 | 61579760 | *FADS1* | T | C | 0.70764 | 0.024018629 | 0.001301119 | $<1\times10^{-16}$ |
| AA | rs174556 | 11 | 61580635 | *FADS1* | C | T | 0.70764 | 0.024026422 | 0.001301439 | $<1\times10^{-16}$ |
| AA | rs174548 | 11 | 61571348 | *FADS1* | C | G | 0.69996 | 0.023714033 | 0.001287432 | $<1\times10^{-16}$ |
| AA | rs174549 | 11 | 61571382 | *FADS1* | G | A | 0.70775 | 0.023812139 | 0.001293691 | $<1\times10^{-16}$ |
| AA | rs174538 | 11 | 61560081 | *C11orf10* | G | A | 0.69487 | 0.023462625 | 0.001279837 | $<1\times10^{-16}$ |
| AA | rs174528 | 11 | 61543499 | *C11orf9* | T | C | 0.61669 | 0.023234781 | 0.001300411 | $<1\times10^{-16}$ |
| AA | rs174601 | 11 | 61623140 | *FADS2* | C | T | 0.61373 | 0.023990221 | 0.001361246 | $<1\times10^{-16}$ |
| AA | rs108499 | 11 | 61547237 | *C11orf9* | C | T | 0.67142 | 0.021386038 | 0.001279824 | $<1\times10^{-16}$ |
| AA | rs174534 | 11 | 61549458 | *C11orf9* | A | G | 0.66316 | 0.020499565 | 0.001244953 | $<1\times10^{-16}$ |
| AA | rs174570 | 11 | 61597212 | *FADS2* | C | T | 0.86422 | 0.025409935 | 0.001896779 | $<1\times10^{-16}$ |
| AA | rs174575 | 11 | 61602003 | *FADS2* | C | G | 0.7332 | 0.018928681 | 0.00146665 | $<1\times10^{-16}$ |
| AA | rs174593 | 11 | 61618831 | *FADS2* | T | C | 0.7208 | 0.018203337 | 0.001464843 | $<1\times10^{-16}$ |
| AA | rs174579 | 11 | 61605613 | *FADS2* | C | T | 0.78402 | 0.018337051 | 0.001519732 | $<1\times10^{-16}$ |
| AA | rs174591 | 11 | 61617676 | *FADS2* | T | A | 0.71832 | 0.017953589 | 0.001491899 | $<1\times10^{-16}$ |
| AA | rs174585 | 11 | 61611694 | *FADS2* | G | A | 0.78304 | 0.018018992 | 0.001511651 | $<1\times10^{-16}$ |

| | | | | | | | | | | |
|---|---|---|---|---|---|---|---|---|---|---|
| AA | rs174448 | 11 | 61639573 | | A | G | 0.64303 | 0.015302061 | 0.00129954 | $<1\times10^{-16}$ |
| AA | rs174589 | 11 | 61615803 | *FADS2* | C | G | 0.78765 | 0.017527017 | 0.001505997 | $<1\times10^{-16}$ |
| AA | rs2727270 | 11 | 61603237 | *FADS2* | C | T | 0.89029 | 0.022375137 | 0.001932447 | $<1\times10^{-16}$ |
| AA | rs2727271 | 11 | 61603358 | *FADS2* | A | T | 0.89 | 0.022344776 | 0.001933855 | $<1\times10^{-16}$ |
| AA | rs509360 | 11 | 61548559 | *C11orf9* | G | A | 0.649 | -0.018655647 | 0.001621963 | $<1\times10^{-16}$ |
| AA | rs2851682 | 11 | 61616012 | *FADS2* | A | G | 0.92129 | 0.028410035 | 0.002486314 | $<1\times10^{-16}$ |
| AA | rs2845573 | 11 | 61601908 | *FADS2* | A | G | 0.92668 | 0.026552235 | 0.002334587 | $<1\times10^{-16}$ |
| AA | rs174449 | 11 | 61640379 | | A | G | 0.63271 | 0.01413323 | 0.001255778 | $<1\times10^{-16}$ |
| AA | rs2524299 | 11 | 61604782 | *FADS2* | A | T | 0.88064 | 0.021480109 | 0.001949754 | $<1\times10^{-16}$ |
| AA | rs174597 | 11 | 61621040 | *FADS2* | G | C | 0.74735 | 0.016773041 | 0.001538603 | $<1\times10^{-16}$ |
| AA | rs968567 | 11 | 61595564 | *FADS2* | C | T | 0.83527 | 0.019099505 | 0.001763269 | $<1\times10^{-16}$ |
| AA | rs422249 | 11 | 61639488 | | C | T | 0.67098 | 0.014503632 | 0.001342686 | $<1\times10^{-16}$ |
| AA | rs2072114 | 11 | 61605215 | *FADS2* | A | G | 0.8814 | 0.020548561 | 0.00191428 | $<1\times10^{-16}$ |
| AA | rs2269928 | 11 | 61537529 | *C11orf9* | T | G | 0.77978 | 0.019499261 | 0.001818698 | $<1\times10^{-16}$ |
| AA | rs174532 | 11 | 61548874 | *C11orf9* | G | A | 0.72114 | -0.017638345 | 0.001693061 | $<1\times10^{-16}$ |
| AA | rs174611 | 11 | 61627881 | *FADS2* | T | C | 0.71039 | 0.014014928 | 0.00135256 | $<1\times10^{-16}$ |
| AA | rs174605 | 11 | 61626921 | *FADS2* | G | T | 0.71151 | 0.013918115 | 0.001347101 | $<1\times10^{-16}$ |
| AA | rs174455 | 11 | 61656117 | *FADS3* | G | A | 0.36538 | -0.012877611 | 0.001259959 | $<1\times10^{-16}$ |
| AA | rs2526678 | 11 | 61623793 | *FADS2* | G | A | 0.93057 | 0.028878715 | 0.002861908 | $<1\times10^{-16}$ |
| AA | rs412334 | 11 | 61560261 | *FEN1* | C | T | 0.85305 | -0.027620235 | 0.002751166 | $<1\times10^{-16}$ |
| AA | rs174616 | 11 | 61629122 | *FADS2* | G | A | 0.52756 | 0.012524792 | 0.001249924 | $<1\times10^{-16}$ |
| AA | rs174626 | 11 | 61637057 | | A | G | 0.52886 | 0.012688919 | 0.001270477 | $<1\times10^{-16}$ |
| AA | rs174450 | 11 | 61641542 | *FADS3* | G | T | 0.47314 | -0.012731387 | 0.001293061 | $<1\times10^{-16}$ |
| AA | rs149803 | 11 | 61539020 | *C11orf9* | C | G | 0.74636 | -0.015835817 | 0.001793894 | $<1\times10^{-16}$ |
| AA | rs174627 | 11 | 61637466 | | G | A | 0.835 | 0.015374786 | 0.001793824 | $<1\times10^{-16}$ |
| AA | rs174468 | 11 | 61663691 | | G | A | 0.60365 | -0.012450023 | 0.001468168 | $<1\times10^{-16}$ |
| AA | rs666870 | 11 | 61677479 | *RAB3IL1* | G | A | 0.6013 | -0.0125676 | 0.001503774 | $1.10\times10^{-16}$ |
| AA | rs174478 | 11 | 61678576 | *RAB3IL1* | T | G | 0.60099 | -0.012569856 | 0.001504329 | $1.10\times10^{-16}$ |
| AA | rs174634 | 11 | 61647387 | *FADS3* | C | G | 0.72588 | 0.011703135 | 0.001401397 | $1.10\times10^{-16}$ |
| AA | rs174479 | 11 | 61678754 | *RAB3IL1* | C | G | 0.81614 | 0.01555274 | 0.001871898 | $1.10\times10^{-16}$ |
| AA | rs174464 | 11 | 61657926 | *FADS3* | G | A | 0.72982 | 0.011685088 | 0.001424754 | $2.20\times10^{-16}$ |
| AA | rs174476 | 11 | 61674118 | *RAB3IL1* | C | T | 0.60208 | -0.012249595 | 0.001499231 | $3.30\times10^{-16}$ |
| AA | rs1000778 | 11 | 61655305 | *FADS3* | G | A | 0.72904 | 0.011228085 | 0.001379677 | $4.40\times10^{-16}$ |
| AA | rs174469 | 11 | 61667443 | *RAB3IL1* | C | T | 0.82555 | 0.014755401 | 0.001813425 | $4.40\times10^{-16}$ |
| AA | rs174456 | 11 | 61656182 | *FADS3* | A | C | 0.72918 | 0.011202139 | 0.001378583 | $4.40\times10^{-16}$ |
| AA | rs526126 | 11 | 61624885 | *FADS2* | C | G | 0.76593 | 0.016088208 | 0.00202466 | $1.90\times10^{-15}$ |

| | | | | | | | | | | |
|---|---|---|---|---|---|---|---|---|---|---|
| AA | rs174602 | 11 | 61624414 | *FADS2* | T | C | 0.76774 | 0.016117955 | 0.00204734 | $3.40\times10^{-15}$ |
| AA | rs198462 | 11 | 61524119 | *C11orf9* | G | A | 0.48644 | -0.010170448 | 0.001297982 | $4.70\times10^{-15}$ |
| AA | rs198476 | 11 | 61525730 | *C11orf9* | G | A | 0.47167 | -0.010063125 | 0.001334135 | $4.60\times10^{-14}$ |
| AA | rs198464 | 11 | 61521621 | | G | A | 0.48411 | -0.009390417 | 0.001253592 | $6.90\times10^{-14}$ |
| AA | rs1692120 | 11 | 61417472 | | G | A | 0.56219 | -0.011385056 | 0.001721542 | $3.80\times10^{-11}$ |
| AA | rs650436 | 11 | 61536430 | *C11orf9* | C | T | 0.57937 | -0.009229438 | 0.00143508 | $1.30\times10^{-10}$ |
| AA | rs2453710 | 11 | 61406542 | *RPLP0P2* | A | G | 0.58218 | -0.011508617 | 0.0018469 | $4.60\times10^{-10}$ |
| DPA-n3 | rs8523 | 6 | 10981053 | *ELOVL2* | A | G | 0.44465 | 0.011293305 | 0.002041436 | $3.20\times10^{-8}$ |
| DPA-n3 | rs3798707 | 6 | 10991935 | *ELOVL2* | T | C | 0.44596 | 0.011325177 | 0.002047072 | $3.20\times10^{-8}$ |
| DPA-n3 | rs4532436 | 6 | 10983971 | *ELOVL2* | G | C | 0.44571 | 0.011295698 | 0.002047548 | $3.50\times10^{-8}$ |
| DPA-n3 | rs4713103 | 6 | 10969141 | *SYCP2L* | T | G | 0.46298 | 0.01132793 | 0.002057244 | $3.70\times10^{-8}$ |
| DPA-n3 | rs1225717 | 6 | 10978240 | | G | A | 0.46545 | 0.011529213 | 0.002095265 | $3.70\times10^{-8}$ |
| DPA-n3 | rs3798711 | 6 | 11002810 | *ELOVL2* | C | T | 0.44625 | 0.01120606 | 0.002047701 | $4.40\times10^{-8}$ |
| DPA-n3 | rs2295602 | 6 | 11005842 | *ELOVL2* | C | T | 0.44643 | 0.011203922 | 0.002047984 | $4.50\times10^{-8}$ |
| DPA-n3 | rs174549 | 11 | 61571382 | *FADS1* | G | A | 0.70775 | 0.01248346 | 0.002243944 | $2.60\times10^{-8}$ |
| DPA-n3 | rs174555 | 11 | 61579760 | *FADS1* | T | C | 0.70764 | 0.012503236 | 0.002257835 | $3.10\times10^{-8}$ |
| DPA-n3 | rs174556 | 11 | 61580635 | *FADS1* | C | T | 0.70764 | 0.01250357 | 0.002258417 | $3.10\times10^{-8}$ |
| DTA | rs174546 | 11 | 61569830 | *FADS1* | C | T | 0.67116 | 0.028954763 | 0.003208542 | $<1\times10^{-16}$ |
| DTA | rs174545 | 11 | 61569306 | *FADS1* | C | G | 0.6711 | 0.028960669 | 0.003208942 | $<1\times10^{-16}$ |
| DTA | rs174547 | 11 | 61570783 | *FADS1* | T | C | 0.67136 | 0.02893559 | 0.003207219 | $<1\times10^{-16}$ |
| DTA | rs174550 | 11 | 61571478 | *FADS1* | T | C | 0.67143 | 0.028901207 | 0.00320778 | $<1\times10^{-16}$ |
| DTA | rs174537 | 11 | 61552680 | *C11orf9* | G | T | 0.66488 | 0.029222614 | 0.003242213 | $<1\times10^{-16}$ |
| DTA | rs174536 | 11 | 61551927 | *C11orf9* | A | C | 0.66386 | 0.029169235 | 0.00324112 | $<1\times10^{-16}$ |
| DTA | rs174535 | 11 | 61551356 | *C11orf9* | T | C | 0.65839 | 0.028930165 | 0.003242579 | $<1\times10^{-16}$ |
| DTA | rs1535 | 11 | 61597972 | *FADS2* | A | G | 0.66595 | 0.029022244 | 0.003272799 | $<1\times10^{-16}$ |
| DTA | rs174538 | 11 | 61560081 | *C11orf10* | G | A | 0.69487 | 0.028802552 | 0.003253572 | $<1\times10^{-16}$ |
| DTA | rs4246215 | 11 | 61564299 | *FEN1* | G | T | 0.65088 | 0.028385671 | 0.003248709 | $<1\times10^{-16}$ |
| DTA | rs174541 | 11 | 61565908 | | T | C | 0.65148 | 0.028344796 | 0.003245402 | $<1\times10^{-16}$ |
| DTA | rs174576 | 11 | 61603510 | *FADS2* | C | A | 0.65841 | 0.027746738 | 0.003302934 | $<1\times10^{-16}$ |
| DTA | rs174574 | 11 | 61600342 | *FADS2* | C | A | 0.63796 | 0.028012059 | 0.00334974 | $1.10\times10^{-16}$ |
| DTA | rs174555 | 11 | 61579760 | *FADS1* | T | C | 0.70764 | 0.027768362 | 0.003322717 | $1.10\times10^{-16}$ |
| DTA | rs174556 | 11 | 61580635 | *FADS1* | C | T | 0.70764 | 0.027778084 | 0.003323559 | $1.10\times10^{-16}$ |
| DTA | rs174549 | 11 | 61571382 | *FADS1* | G | A | 0.70775 | 0.027546529 | 0.003302683 | $1.10\times10^{-16}$ |
| DTA | rs102275 | 11 | 61557803 | *C11orf10* | T | C | 0.62686 | 0.027917164 | 0.003355333 | $1.10\times10^{-16}$ |
| DTA | rs174577 | 11 | 61604814 | *FADS2* | C | A | 0.6524 | 0.02741364 | 0.00330046 | $1.10\times10^{-16}$ |
| DTA | rs174548 | 11 | 61571348 | *FADS1* | C | G | 0.69996 | 0.027024764 | 0.003288992 | $2.20\times10^{-16}$ |

| | | | | | | | | | | |
|---|---|---|---|---|---|---|---|---|---|---|
| DTA | rs174578 | 11 | 61605499 | *FADS2* | T | A | 0.65154 | 0.026865055 | 0.003296302 | $3.30\times10^{-16}$ |
| DTA | rs174583 | 11 | 61609750 | *FADS2* | C | T | 0.65299 | 0.026853695 | 0.003314948 | $5.60\times10^{-16}$ |
| DTA | rs174534 | 11 | 61549458 | *C11orf9* | A | G | 0.66316 | 0.0246155 | 0.003138156 | $4.30\times10^{-15}$ |
| DTA | rs108499 | 11 | 61547237 | *C11orf9* | C | T | 0.67142 | 0.025336206 | 0.003231711 | $4.60\times10^{-15}$ |
| DTA | rs174528 | 11 | 61543499 | *C11orf9* | T | C | 0.61669 | 0.025454057 | 0.003314775 | $1.60\times10^{-14}$ |
| DTA | rs174601 | 11 | 61623140 | *FADS2* | C | T | 0.61373 | 0.025711832 | 0.003466068 | $1.20\times10^{-13}$ |
| DTA | rs174579 | 11 | 61605613 | *FADS2* | C | T | 0.78402 | 0.024832406 | 0.003742543 | $3.20\times10^{-11}$ |
| DTA | rs174585 | 11 | 61611694 | *FADS2* | G | A | 0.78304 | 0.024103702 | 0.003722047 | $9.40\times10^{-11}$ |
| DTA | rs174589 | 11 | 61615803 | *FADS2* | C | G | 0.78765 | 0.023079185 | 0.003705559 | $4.70\times10^{-10}$ |
| DTA | rs174575 | 11 | 61602003 | *FADS2* | C | G | 0.7332 | 0.022193588 | 0.00363031 | $9.80\times10^{-10}$ |
| DTA | rs174570 | 11 | 61597212 | *FADS2* | C | T | 0.86422 | 0.028406585 | 0.004715405 | $1.70\times10^{-9}$ |
| DTA | rs968567 | 11 | 61595564 | *FADS2* | C | T | 0.83527 | 0.025899899 | 0.004325846 | $2.10\times10^{-9}$ |
| DTA | rs2269928 | 11 | 61537529 | *C11orf9* | T | G | 0.77978 | 0.026124322 | 0.004452842 | $4.40\times10^{-9}$ |
| DTA | rs174593 | 11 | 61618831 | *FADS2* | T | C | 0.7208 | 0.020456431 | 0.003622509 | $1.60\times10^{-8}$ |

PUFA, polyunsaturated fatty acid; SNP, single nucleotide polymorphism; Chr, chromosome; EAF, Effect allele frequency; SE, standard error. For the 413 genome-wide significant associations (188 unique SNPs) reported by Tintle *et al.* 2015, a total of 369 associations (171 unique SNPs) remained significant at $p < 5.0\times10^{-8}$ in our repeating analysis with the Framingham Heart Study. Their summary statistics were reported in this table. Among them, genetic instruments were further selected based on different LD cutoffs.

**Supplementary Table 6. Forward Mendelian randomization estimates of associations of genetically predicted polyunsaturated fatty acids with COVID-19 severity based on the release 5 HGI A2.**

| | | | IVW_MRE | | | MR-Egger | | | Wald ratio | | | MR-Egger intercept | | Heterogeneity test | | Weighted median | | | | |
|---|---|---|---|---|---|---|---|---|---|---|---|---|---|---|---|---|---|---|---|---|
| **Group name** | **PUFA** | **$R^2$** | **β** | **SE** | ***P*** | **β** | **SE** | ***P*** | **β** | **SE** | ***P*** | **β** | ***P*** | ***P*_IVW** | ***P*_Egger** | **β** | **SE** | ***P*** | **nsnps** | ***F*-statistic** |
| Plasma | ALA | 0.001 | - | - | - | - | - | - | 0.107 | 0.089 | 0.23 | - | - | - | - | - | - | - | 1 | 256 |
| Plasma | ALA | 0.01 | - | - | - | - | - | - | 0.108 | 0.09 | 0.23 | - | - | - | - | - | - | - | 1 | 312 |
| Plasma | ALA | 0.1 | 0.099 | 0.027 | 2.00E-04 | 0.042 | 0.17 | 0.828 | - | - | - | 0.016 | 0.745 | 0.935 | 0.866 | 0.108 | 0.078 | 0.164 | 4 | 436 |
| Plasma | ALA | 0.3 | 0.125 | 0.046 | 0.006 | -0.075 | 0.132 | 0.58 | - | - | - | 0.05 | 0.134 | 0.441 | 0.582 | 0.103 | 0.063 | 0.104 | 13 | 1107 |
| RBC | ALA | 0.001 | - | - | - | - | - | - | 0.048 | 0.172 | 0.78 | - | - | - | - | - | - | - | 1 | 31 |
| RBC | ALA | 0.01 | - | - | - | - | - | - | 0.048 | 0.172 | 0.78 | - | - | - | - | - | - | - | 1 | 31 |
| RBC | ALA | 0.1 | - | - | - | - | - | - | 0.048 | 0.172 | 0.78 | - | - | - | - | - | - | - | 1 | 31 |
| RBC | ALA | 0.3 | - | - | - | - | - | - | 0.048 | 0.172 | 0.78 | - | - | - | - | - | - | - | 1 | 31 |
| Plasma | LA | 0.001 | 0.073 | 0.038 | 0.053 | 0.013 | 0.081 | 0.899 | - | - | - | 0.027 | 0.517 | 0.574 | 0.645 | 0.065 | 0.051 | 0.204 | 3 | 1329 |
| RBC | LA | 0.001 | - | - | - | - | - | - | 0.04 | 0.079 | 0.611 | - | - | - | - | - | - | - | 1 | 133 |
| RBC | LA | 0.01 | - | - | - | - | - | - | 0.04 | 0.079 | 0.611 | - | - | - | - | - | - | - | 1 | 133 |
| RBC | LA | 0.1 | 0.057 | 0.03 | 0.054 | - | - | - | - | - | - | - | - | - | - | - | - | - | 2 | 175 |
| RBC | LA | 0.3 | 0.072 | 0.037 | 0.053 | -0.099 | 0.2 | 0.635 | - | - | - | 0.05 | 0.414 | 0.534 | 0.509 | 0.042 | 0.051 | 0.406 | 9 | 566 |
| Plasma | GLA | 0.001 | -0.11 | 0.066 | 0.094 | - | - | - | - | - | - | - | - | - | - | - | - | - | 2 | 293 |
| RBC | GLA | 0.001 | - | - | - | - | - | - | -0.038 | 0.138 | 0.78 | - | - | - | - | - | - | - | 1 | 49 |
| RBC | GLA | 0.01 | - | - | - | - | - | - | -0.038 | 0.138 | 0.78 | - | - | - | - | - | - | - | 1 | 49 |
| RBC | GLA | 0.1 | - | - | - | - | - | - | -0.038 | 0.138 | 0.78 | - | - | - | - | - | - | - | 1 | 49 |
| RBC | GLA | 0.3 | - | - | - | - | - | - | -0.038 | 0.138 | 0.78 | - | - | - | - | - | - | - | 1 | 49 |
| Plasma | DGLA | 0.001 | 0.069 | 0.011 | 7.00E-11 | - | - | - | - | - | - | - | - | - | - | - | - | - | 2 | 1417 |
| RBC | DGLA | 0.001 | 0.017 | 0.103 | 0.87 | - | - | - | - | - | - | - | - | - | - | - | - | - | 2 | 542 |
| RBC | DGLA | 0.01 | 0.044 | 0.056 | 0.436 | -0.103 | 0.125 | 0.498 | - | - | - | 0.085 | 0.329 | 0.042 | 0.104 | 0.059 | 0.047 | 0.201 | 4 | 788 |
| RBC | DGLA | 0.1 | 0.045 | 0.019 | 0.017 | 0.015 | 0.045 | 0.747 | - | - | - | 0.02 | 0.464 | 0.488 | 0.453 | 0.01 | 0.026 | 0.708 | 14 | 2375 |
| RBC | DGLA | 0.3 | 0.041 | 0.012 | 0.001 | -3.00E-04 | 0.032 | 0.992 | - | - | - | 0.027 | 0.168 | 0.672 | 0.733 | 0.016 | 0.019 | 0.408 | 26 | 5970 |
| Plasma | AA | 0.001 | -0.039 | 0.014 | 0.007 | - | - | - | - | - | - | - | - | - | - | - | - | - | 2 | 4616 |
| RBC | AA | 0.001 | -0.029 | 0.015 | 0.048 | -0.089 | 0.182 | 0.712 | - | - | - | 0.024 | 0.788 | 0.938 | 0.927 | -0.035 | 0.066 | 0.599 | 3 | 268 |
| RBC | AA | 0.01 | -0.041 | 0.021 | 0.049 | -0.114 | 0.166 | 0.616 | - | - | - | 0.031 | 0.72 | 0.872 | 0.816 | -0.044 | 0.065 | 0.493 | 3 | 287 |
| RBC | AA | 0.1 | -0.111 | 0.038 | 0.003 | -0.038 | 0.129 | 0.782 | - | - | - | -0.029 | 0.575 | 0.429 | 0.352 | -0.081 | 0.05 | 0.109 | 7 | 584 |
| RBC | AA | 0.3 | -0.078 | 0.024 | 0.001 | -0.015 | 0.076 | 0.842 | - | - | - | -0.026 | 0.4 | 0.345 | 0.33 | -0.051 | 0.033 | 0.118 | 14 | 1440 |
| Plasma | DPA-n3 | 0.001 | -0.114 | 0.051 | 0.026 | -0.066 | 0.157 | 0.746 | - | - | - | -0.016 | 0.792 | 0.426 | 0.216 | -0.113 | 0.058 | 0.051 | 3 | 835 |
| Plasma | DPA-n3 | 0.01 | -0.115 | 0.051 | 0.024 | -0.069 | 0.157 | 0.736 | - | - | - | -0.016 | 0.801 | 0.434 | 0.219 | -0.114 | 0.058 | 0.05 | 3 | 937 |
| Plasma | DPA-n3 | 0.1 | -0.116 | 0.027 | 2.00E-05 | -0.047 | 0.082 | 0.578 | - | - | - | -0.021 | 0.358 | 0.954 | 0.963 | -0.087 | 0.054 | 0.108 | 14 | 1506 |
| Plasma | DPA-n3 | 0.3 | -0.119 | 0.025 | 1.00E-06 | -0.02 | 0.062 | 0.744 | - | - | - | -0.03 | 0.084 | 0.77 | 0.869 | -0.084 | 0.041 | 0.039 | 29 | 3466 |
| RBC | DPA-n3 | 0.001 | -0.237 | 0.116 | 0.041 | - | - | - | - | - | - | - | - | - | - | - | - | - | 2 | 62 |
| RBC | DPA-n3 | 0.01 | -0.237 | 0.116 | 0.041 | - | - | - | - | - | - | - | - | - | - | - | - | - | 2 | 62 |
| RBC | DPA-n3 | 0.1 | -0.237 | 0.116 | 0.041 | - | - | - | - | - | - | - | - | - | - | - | - | - | 2 | 62 |
| RBC | DPA-n3 | 0.3 | -0.237 | 0.116 | 0.041 | - | - | - | - | - | - | - | - | - | - | - | - | - | 2 | 62 |
| RBC | DTA | 0.001 | - | - | - | - | - | - | -0.091 | 0.104 | 0.382 | - | - | - | - | - | - | - | 1 | 80 |
| RBC | DTA | 0.01 | - | - | - | - | - | - | -0.091 | 0.104 | 0.382 | - | - | - | - | - | - | - | 1 | 80 |
| RBC | DTA | 0.1 | - | - | - | - | - | - | -0.091 | 0.104 | 0.382 | - | - | - | - | - | - | - | 1 | 80 |
| RBC | DTA | 0.3 | -0.081 | 0.016 | 7.00E-07 | - | - | - | - | - | - | - | - | - | - | - | - | - | 2 | 114 |
| Plasma | DHA | 0.001 | - | - | - | - | - | - | 0.47 | 0.212 | 0.026 | - | - | - | - | - | - | - | 1 | 65 |
| Plasma | DHA | 0.01 | - | - | - | - | - | - | 0.469 | 0.211 | 0.026 | - | - | - | - | - | - | - | 1 | 64 |
| Plasma | DHA | 0.1 | - | - | - | - | - | - | 0.469 | 0.211 | 0.026 | - | - | - | - | - | - | - | 1 | 64 |
| Plasma | DHA | 0.3 | 0.393 | 0.137 | 0.004 | - | - | - | - | - | - | - | - | - | - | - | - | - | 2 | 95 |

PUFA, polyunsaturated fatty acid; RBC, red blood cell; ALA, α-Linolenic acid; LA: linoleic acid; GLA, γ-Linoleic acid; DGLA, dihomo-γ-linoleic acid; AA, arachidonic acid; DPA-n3, docosapentaenoic acid; DTA, docosatetraenoic acid; DHA, docosahexaenoic acid; β, causal effect size; SE, standard error; IVW_MRE, inverse-variance weighted random-effects model; Egger, MR-Egger; nsnps, number of SNPs retained for this analysis.

**Supplementary Table 7. Forward Mendelian randomization estimates of associations of genetically predicted polyunsaturated fatty acids with COVID-19 severity based on the release 5 HGI B2.**

| | | | IVW_MRE | | | MR-Egger | | | Wald ratio | | | MR-Egger intercept | | Heterogeneity test | | Weighted median | | | | |
|---|---|---|---|---|---|---|---|---|---|---|---|---|---|---|---|---|---|---|---|---|
| **Group name** | **PUFA** | **$R^2$** | **β** | **SE** | ***P*** | **β** | **SE** | ***P*** | **β** | **SE** | ***P*** | **β** | ***P*** | ***P*_IVW** | ***P*_Egger** | **β** | **SE** | ***P*** | **nsnps** | ***F*-statistic** |
| Plasma | ALA | 0.001 | - | - | - | - | - | - | 0.101 | 0.061 | 0.098 | - | - | - | - | - | - | - | 1 | 256 |
| Plasma | ALA | 0.01 | - | - | - | - | - | - | 0.101 | 0.061 | 0.098 | - | - | - | - | - | - | - | 1 | 312 |
| Plasma | ALA | 0.1 | 0.109 | 0.083 | 0.192 | -0.090 | 0.195 | 0.689 | - | - | - | 0.057 | 0.380 | 0.020 | 0.049 | 0.118 | 0.056 | 0.034 | 4 | 436 |
| Plasma | ALA | 0.3 | 0.101 | 0.038 | 0.008 | -0.108 | 0.096 | 0.284 | - | - | - | 0.052 | 0.041 | 0.042 | 0.203 | 0.094 | 0.039 | 0.016 | 13 | 1107 |
| RBC | ALA | 0.001 | - | - | - | - | - | - | 0.122 | 0.116 | 0.291 | - | - | - | - | - | - | - | 1 | 31 |
| RBC | ALA | 0.01 | - | - | - | - | - | - | 0.122 | 0.116 | 0.291 | - | - | - | - | - | - | - | 1 | 31 |
| RBC | ALA | 0.1 | - | - | - | - | - | - | 0.122 | 0.116 | 0.291 | - | - | - | - | - | - | - | 1 | 31 |
| RBC | ALA | 0.3 | - | - | - | - | - | - | 0.122 | 0.116 | 0.291 | - | - | - | - | - | - | - | 1 | 31 |
| Plasma | LA | 0.001 | 0.058 | 0.024 | 0.015 | 0.052 | 0.055 | 0.517 | - | - | - | 0.003 | 0.915 | 0.617 | 0.331 | 0.057 | 0.036 | 0.108 | 3 | 1329 |
| RBC | LA | 0.001 | - | - | - | - | - | - | 0.082 | 0.053 | 0.126 | - | - | - | - | - | - | - | 1 | 133 |
| RBC | LA | 0.01 | - | - | - | - | - | - | 0.082 | 0.053 | 0.126 | - | - | - | - | - | - | - | 1 | 133 |
| RBC | LA | 0.1 | 0.109 | 0.048 | 0.024 | - | - | - | - | - | - | - | - | - | - | - | - | - | 2 | 175 |
| RBC | LA | 0.3 | 0.076 | 0.023 | 0.001 | -0.045 | 0.136 | 0.752 | - | - | - | 0.036 | 0.397 | 0.622 | 0.610 | 0.074 | 0.034 | 0.030 | 9 | 566 |
| Plasma | GLA | 0.001 | -0.117 | 0.013 | 6.60e-20 | - | - | - | - | - | - | - | - | - | - | - | - | - | 2 | 293 |
| RBC | GLA | 0.001 | - | - | - | - | - | - | -0.098 | 0.093 | 0.291 | - | - | - | - | - | - | - | 1 | 49 |
| RBC | GLA | 0.01 | - | - | - | - | - | - | -0.098 | 0.093 | 0.291 | - | - | - | - | - | - | - | 1 | 49 |
| RBC | GLA | 0.1 | - | - | - | - | - | - | -0.098 | 0.093 | 0.291 | - | - | - | - | - | - | - | 1 | 49 |
| RBC | GLA | 0.3 | - | - | - | - | - | - | -0.098 | 0.093 | 0.291 | - | - | - | - | - | - | - | 1 | 49 |
| Plasma | DGLA | 0.001 | 0.048 | 0.040 | 0.231 | - | - | - | - | - | - | - | - | - | - | - | - | - | 2 | 1417 |
| RBC | DGLA | 0.001 | 0.010 | 0.044 | 0.828 | - | - | - | - | - | - | - | - | - | - | - | - | - | 2 | 542 |
| RBC | DGLA | 0.01 | 0.006 | 0.022 | 0.774 | -0.069 | 0.054 | 0.328 | - | - | - | 0.044 | 0.266 | 0.332 | 0.582 | -0.007 | 0.024 | 0.760 | 4 | 788 |
| RBC | DGLA | 0.1 | 0.029 | 0.017 | 0.080 | 0.039 | 0.040 | 0.353 | - | - | - | -0.006 | 0.795 | 0.091 | 0.065 | 0.023 | 0.017 | 0.177 | 13 | 2341 |
| RBC | DGLA | 0.3 | 0.033 | 0.010 | 0.001 | 0.006 | 0.024 | 0.819 | - | - | - | 0.017 | 0.238 | 0.089 | 0.106 | 0.026 | 0.011 | 0.021 | 25 | 5936 |
| Plasma | AA | 0.001 | -0.038 | 0.004 | 3.23e-20 | - | - | - | - | - | - | - | - | - | - | - | - | - | 2 | 4616 |
| RBC | AA | 0.001 | -0.049 | 0.011 | 1.32e-05 | -0.097 | 0.123 | 0.574 | - | - | - | 0.019 | 0.753 | 0.912 | 0.894 | -0.053 | 0.041 | 0.189 | 3 | 268 |
| RBC | AA | 0.01 | -0.056 | 0.015 | 1.78e-04 | -0.112 | 0.113 | 0.502 | - | - | - | 0.024 | 0.694 | 0.844 | 0.795 | -0.059 | 0.039 | 0.135 | 3 | 287 |
| RBC | AA | 0.1 | -0.069 | 0.015 | 3.89e-06 | -0.056 | 0.082 | 0.528 | - | - | - | -0.005 | 0.871 | 0.888 | 0.807 | -0.059 | 0.030 | 0.047 | 7 | 584 |
| RBC | AA | 0.3 | -0.055 | 0.012 | 9.17e-06 | -0.039 | 0.048 | 0.425 | - | - | - | -0.006 | 0.736 | 0.791 | 0.734 | -0.052 | 0.021 | 0.012 | 14 | 1440 |
| Plasma | DPA-n3 | 0.001 | -0.069 | 0.007 | 4.73e-20 | -0.089 | 0.082 | 0.475 | - | - | - | 0.007 | 0.827 | 0.962 | 0.977 | -0.070 | 0.038 | 0.067 | 3 | 835 |
| Plasma | DPA-n3 | 0.01 | -0.069 | 0.008 | 6.21e-20 | -0.089 | 0.083 | 0.475 | - | - | - | 0.007 | 0.826 | 0.961 | 0.990 | -0.070 | 0.039 | 0.073 | 3 | 937 |
| Plasma | DPA-n3 | 0.1 | -0.076 | 0.028 | 0.007 | 0.012 | 0.053 | 0.830 | - | - | - | -0.027 | 0.081 | 0.302 | 0.491 | -0.070 | 0.034 | 0.042 | 14 | 1506 |
| Plasma | DPA-n3 | 0.3 | -0.072 | 0.019 | 1.65e-04 | 0.015 | 0.040 | 0.710 | - | - | - | -0.026 | 0.022 | 0.188 | 0.401 | -0.065 | 0.026 | 0.012 | 29 | 3466 |
| RBC | DPA-n3 | 0.001 | -0.130 | 0.033 | 9.30e-05 | - | - | - | - | - | - | - | - | - | - | - | - | - | 2 | 62 |
| RBC | DPA-n3 | 0.01 | -0.130 | 0.033 | 9.30e-05 | - | - | - | - | - | - | - | - | - | - | - | - | - | 2 | 62 |
| RBC | DPA-n3 | 0.1 | -0.130 | 0.033 | 9.30e-05 | - | - | - | - | - | - | - | - | - | - | - | - | - | 2 | 62 |
| RBC | DPA-n3 | 0.3 | -0.130 | 0.033 | 9.30e-05 | - | - | - | - | - | - | - | - | - | - | - | - | - | 2 | 62 |
| RBC | DTA | 0.001 | - | - | - | - | - | - | -0.102 | 0.071 | 0.148 | - | - | - | - | - | - | - | 1 | 80 |
| RBC | DTA | 0.01 | - | - | - | - | - | - | -0.102 | 0.071 | 0.148 | - | - | - | - | - | - | - | 1 | 80 |
| RBC | DTA | 0.1 | - | - | - | - | - | - | -0.102 | 0.071 | 0.148 | - | - | - | - | - | - | - | 1 | 80 |
| RBC | DTA | 0.3 | -0.098 | 0.006 | 3.04e-60 | - | - | - | - | - | - | - | - | - | - | - | - | - | 2 | 114 |
| Plasma | DHA | 0.001 | - | - | - | - | - | - | 0.161 | 0.146 | 0.269 | - | - | - | - | - | - | - | 1 | 65 |
| Plasma | DHA | 0.01 | - | - | - | - | - | - | 0.161 | 0.146 | 0.269 | - | - | - | - | - | - | - | 1 | 64 |
| Plasma | DHA | 0.1 | - | - | - | - | - | - | 0.161 | 0.146 | 0.269 | - | - | - | - | - | - | - | 1 | 64 |
| Plasma | DHA | 0.3 | 0.170 | 0.014 | 5.32e-34 | - | - | - | - | - | - | - | - | - | - | - | - | - | 2 | 95 |

PUFA, polyunsaturated fatty acid; RBC, red blood cell; ALA, α-Linolenic acid; LA: linoleic acid; GLA, γ-Linoleic acid; DGLA, dihomo-γ-linoleic acid; AA, arachidonic acid; DPA-n3, docosapentaenoic acid; DTA, docosatetraenoic acid; DHA, docosahexaenoic acid; β, causal effect size; SE, standard error; IVW_MRE, inverse-variance weighted random-effects model; Egger, MR-Egger; nsnps, number of SNPs retained for this analysis.

**Supplementary Table 8. Forward Mendelian randomization estimates of associations of genetically predicted polyunsaturated fatty acids with COVID-19 severity based on the release 5 HGI B1.**

| | | | IVW_MRE | | | MR-Egger | | | Wald ratio | | | MR-Egger intercept | | Heterogeneity test | | Weighted median | | | | |
|---|---|---|---|---|---|---|---|---|---|---|---|---|---|---|---|---|---|---|---|---|
| **Group name** | **PUFA** | **$R^2$** | **β** | **SE** | ***P*** | **β** | **SE** | ***P*** | **β** | **SE** | ***P*** | **β** | ***P*** | ***P*_IVW** | ***P*_Egger** | **β** | **SE** | ***P*** | **nsnps** | ***F*-statistic** |
| Plasma | ALA | 0.001 | - | - | - | - | - | - | 0.211 | 0.102 | 0.038 | - | - | - | - | - | - | - | 1 | 256 |
| Plasma | ALA | 0.01 | - | - | - | - | - | - | 0.213 | 0.102 | 0.038 | - | - | - | - | - | - | - | 1 | 312 |
| Plasma | ALA | 0.1 | 0.224 | 0.179 | 0.211 | -0.174 | 0.45 | 0.737 | - | - | - | 0.115 | 0.437 | 0.001 | 0.002 | 0.229 | 0.097 | 0.018 | 4 | 436 |
| Plasma | ALA | 0.3 | 0.183 | 0.07 | 0.009 | -0.053 | 0.204 | 0.8 | - | - | - | 0.058 | 0.245 | 0.004 | 0.007 | 0.209 | 0.065 | 0.001 | 13 | 1107 |
| RBC | ALA | 0.001 | - | - | - | - | - | - | 0.362 | 0.193 | 0.061 | - | - | - | - | - | - | - | 1 | 31 |
| RBC | ALA | 0.01 | - | - | - | - | - | - | 0.362 | 0.193 | 0.061 | - | - | - | - | - | - | - | 1 | 31 |
| RBC | ALA | 0.1 | - | - | - | - | - | - | 0.362 | 0.193 | 0.061 | - | - | - | - | - | - | - | 1 | 31 |
| RBC | ALA | 0.3 | - | - | - | - | - | - | 0.362 | 0.193 | 0.061 | - | - | - | - | - | - | - | 1 | 31 |
| Plasma | LA | 0.001 | 0.112 | 0.071 | 0.117 | 0.138 | 0.159 | 0.545 | - | - | - | -0.011 | 0.868 | 0.213 | 0.085 | 0.113 | 0.058 | 0.05 | 3 | 1329 |
| RBC | LA | 0.001 | - | - | - | - | - | - | 0.227 | 0.09 | 0.011 | - | - | - | - | - | - | - | 1 | 133 |
| RBC | LA | 0.01 | - | - | - | - | - | - | 0.227 | 0.09 | 0.011 | - | - | - | - | - | - | - | 1 | 133 |
| RBC | LA | 0.1 | 0.27 | 0.076 | 3.90E-04 | - | - | - | - | - | - | - | - | - | - | - | - | - | 2 | 175 |
| RBC | LA | 0.3 | 0.183 | 0.038 | 1.75E-06 | 0.057 | 0.228 | 0.81 | - | - | - | 0.037 | 0.592 | 0.55 | 0.476 | 0.216 | 0.056 | 1.04E-04 | 9 | 566 |
| Plasma | GLA | 0.001 | -0.265 | 0.031 | 2.82E-17 | - | - | - | - | - | - | - | - | - | - | - | - | - | 2 | 293 |
| RBC | GLA | 0.001 | - | - | - | - | - | - | -0.29 | 0.155 | 0.061 | - | - | - | - | - | - | - | 1 | 49 |
| RBC | GLA | 0.01 | - | - | - | - | - | - | -0.29 | 0.155 | 0.061 | - | - | - | - | - | - | - | 1 | 49 |
| RBC | GLA | 0.1 | - | - | - | - | - | - | -0.29 | 0.155 | 0.061 | - | - | - | - | - | - | - | 1 | 49 |
| RBC | GLA | 0.3 | - | - | - | - | - | - | -0.29 | 0.155 | 0.061 | - | - | - | - | - | - | - | 1 | 49 |
| Plasma | DGLA | 0.001 | 0.074 | 0.118 | 0.528 | - | - | - | - | - | - | - | - | - | - | - | - | - | 2 | 1417 |
| RBC | DGLA | 0.001 | 0.005 | 0.025 | 0.84 | - | - | - | - | - | - | - | - | - | - | - | - | - | 2 | 542 |
| RBC | DGLA | 0.01 | -0.015 | 0.027 | 0.591 | -0.014 | 0.086 | 0.886 | - | - | - | -4.98E-04 | 0.993 | 0.55 | 0.348 | -0.006 | 0.036 | 0.876 | 4 | 788 |
| RBC | DGLA | 0.1 | 0.048 | 0.029 | 0.101 | 0.087 | 0.07 | 0.24 | - | - | - | -0.025 | 0.551 | 0.029 | 0.024 | 0.053 | 0.029 | 0.069 | 13 | 2341 |
| RBC | DGLA | 0.3 | 0.059 | 0.017 | 0.001 | 0.049 | 0.044 | 0.279 | - | - | - | 0.007 | 0.804 | 0.02 | 0.014 | 0.054 | 0.02 | 0.007 | 25 | 5936 |
| Plasma | AA | 0.001 | -0.082 | 0.031 | 0.008 | - | - | - | - | - | - | - | - | - | - | - | - | - | 2 | 4616 |
| RBC | AA | 0.001 | -0.171 | 0.007 | 8.57E-130 | -0.139 | 0.204 | 0.621 | - | - | - | -0.013 | 0.895 | 0.986 | 0.998 | -0.167 | 0.067 | 0.013 | 3 | 268 |
| RBC | AA | 0.01 | -0.167 | 0.009 | 4.60E-80 | -0.13 | 0.188 | 0.615 | - | - | - | -0.016 | 0.871 | 0.978 | 0.956 | -0.164 | 0.066 | 0.012 | 3 | 287 |
| RBC | AA | 0.1 | -0.109 | 0.031 | 4.43E-04 | -0.185 | 0.137 | 0.234 | - | - | - | 0.03 | 0.587 | 0.687 | 0.61 | -0.113 | 0.05 | 0.025 | 7 | 584 |
| RBC | AA | 0.3 | -0.1 | 0.025 | 5.99E-05 | -0.137 | 0.083 | 0.126 | - | - | - | 0.015 | 0.652 | 0.418 | 0.358 | -0.114 | 0.032 | 3.62E-04 | 14 | 1440 |
| Plasma | DPA-n3 | 0.001 | -0.131 | 0.028 | 3.59E-06 | -0.164 | 0.132 | 0.431 | - | - | - | 0.011 | 0.82 | 0.818 | 0.573 | -0.149 | 0.064 | 0.021 | 3 | 835 |
| Plasma | DPA-n3 | 0.01 | -0.131 | 0.029 | 6.00E-06 | -0.164 | 0.132 | 0.433 | - | - | - | 0.011 | 0.824 | 0.811 | 0.561 | -0.15 | 0.067 | 0.025 | 3 | 937 |
| Plasma | DPA-n3 | 0.1 | -0.138 | 0.058 | 0.018 | -0.017 | 0.115 | 0.887 | - | - | - | -0.038 | 0.244 | 0.02 | 0.031 | -0.153 | 0.064 | 0.017 | 14 | 1506 |
| Plasma | DPA-n3 | 0.3 | -0.121 | 0.034 | 3.48E-04 | -0.028 | 0.075 | 0.718 | - | - | - | -0.028 | 0.178 | 0.034 | 0.048 | -0.15 | 0.041 | 2.88E-04 | 29 | 3466 |
| RBC | DPA-n3 | 0.001 | -0.274 | 0.131 | 0.036 | - | - | - | - | - | - | - | - | - | - | - | - | - | 2 | 62 |
| RBC | DPA-n3 | 0.01 | -0.274 | 0.131 | 0.036 | - | - | - | - | - | - | - | - | - | - | - | - | - | 2 | 62 |
| RBC | DPA-n3 | 0.1 | -0.274 | 0.131 | 0.036 | - | - | - | - | - | - | - | - | - | - | - | - | - | 2 | 62 |
| RBC | DPA-n3 | 0.3 | -0.274 | 0.131 | 0.036 | - | - | - | - | - | - | - | - | - | - | - | - | - | 2 | 62 |
| RBC | DTA | 0.001 | - | - | - | - | - | - | -0.25 | 0.118 | 0.034 | - | - | - | - | - | - | - | 1 | 80 |
| RBC | DTA | 0.01 | - | - | - | - | - | - | -0.25 | 0.118 | 0.034 | - | - | - | - | - | - | - | 1 | 80 |
| RBC | DTA | 0.1 | - | - | - | - | - | - | -0.25 | 0.118 | 0.034 | - | - | - | - | - | - | - | 1 | 80 |
| RBC | DTA | 0.3 | -0.271 | 0.027 | 2.21E-23 | - | - | - | - | - | - | - | - | - | - | - | - | - | 2 | 114 |
| Plasma | DHA | 0.001 | - | - | - | - | - | - | 0.103 | 0.246 | 0.674 | - | - | - | - | - | - | - | 1 | 65 |
| Plasma | DHA | 0.01 | - | - | - | - | - | - | 0.103 | 0.245 | 0.674 | - | - | - | - | - | - | - | 1 | 64 |
| Plasma | DHA | 0.1 | - | - | - | - | - | - | 0.103 | 0.245 | 0.674 | - | - | - | - | - | - | - | 1 | 64 |
| Plasma | DHA | 0.3 | -0.005 | 0.158 | 0.974 | - | - | - | - | - | - | - | - | - | - | - | - | - | 2 | 95 |

PUFA, polyunsaturated fatty acid; RBC, red blood cell; ALA, α-Linolenic acid; LA: linoleic acid; GLA, γ-Linoleic acid; DGLA, dihomo-γ-linoleic acid; AA, arachidonic acid; DPA-n3, docosapentaenoic acid; DTA, docosatetraenoic acid; DHA, docosahexaenoic acid; β, causal effect size; SE, standard error; IVW_MRE, inverse-variance weighted random-effects model; Egger, MR-Egger; nsnps, number of SNPs retained for this analysis.

**Supplementary Table 9. Forward Mendelian randomization estimates of associations of genetically predicted polyunsaturated fatty acids with COVID-19 susceptibility based on the release 5 HGI C2.**

| | | | IVW_MRE | | | MR-Egger | | | Wald ratio | | | MR-Egger intercept | | Heterogeneity test | | Weighted median | | | | |
|---|---|---|---|---|---|---|---|---|---|---|---|---|---|---|---|---|---|---|---|---|
| **Group name** | **PUFA** | **$R^2$** | **β** | **SE** | ***P*** | **β** | **SE** | ***P*** | **β** | **SE** | ***P*** | **β** | ***P*** | ***P*_IVW** | ***P*_Egger** | **β** | **SE** | ***P*** | **nsnps** | ***F*-statistic** |
| Plasma | ALA | 0.001 | - | - | - | - | - | - | 0.012 | 0.031 | 0.697 | - | - | - | - | - | - | - | 1 | 256 |
| Plasma | ALA | 0.01 | - | - | - | - | - | - | 0.012 | 0.031 | 0.697 | - | - | - | - | - | - | - | 1 | 312 |
| Plasma | ALA | 0.1 | 0.010 | 0.008 | 0.211 | 0.031 | 0.056 | 0.636 | - | - | - | -0.006 | 0.728 | 0.938 | 0.882 | 0.010 | 0.025 | 0.693 | 4 | 436 |
| Plasma | ALA | 0.3 | 0.012 | 0.009 | 0.200 | -0.012 | 0.041 | 0.770 | - | - | - | 0.006 | 0.545 | 0.943 | 0.930 | 0.012 | 0.017 | 0.477 | 13 | 1107 |
| RBC | ALA | 0.001 | - | - | - | - | - | - | -4.57e-04 | 0.059 | 0.994 | - | - | - | - | - | - | - | 1 | 31 |
| RBC | ALA | 0.01 | - | - | - | - | - | - | -4.57e-04 | 0.059 | 0.994 | - | - | - | - | - | - | - | 1 | 31 |
| RBC | ALA | 0.1 | - | - | - | - | - | - | -4.57e-04 | 0.059 | 0.994 | - | - | - | - | - | - | - | 1 | 31 |
| RBC | ALA | 0.3 | - | - | - | - | - | - | -4.57e-04 | 0.059 | 0.994 | - | - | - | - | - | - | - | 1 | 31 |
| Plasma | LA | 0.001 | 0.007 | 0.009 | 0.430 | 0.010 | 0.028 | 0.779 | - | - | - | -0.001 | 0.914 | 0.761 | 0.468 | 0.008 | 0.018 | 0.669 | 3 | 1329 |
| RBC | LA | 0.001 | - | - | - | - | - | - | 2.25e-04 | 0.028 | 0.993 | - | - | - | - | - | - | - | 1 | 133 |
| RBC | LA | 0.01 | - | - | - | - | - | - | 2.25e-04 | 0.028 | 0.993 | - | - | - | - | - | - | - | 1 | 133 |
| RBC | LA | 0.1 | -0.004 | 0.008 | 0.594 | - | - | - | - | - | - | - | - | - | - | - | - | - | 2 | 175 |
| RBC | LA | 0.3 | -0.005 | 0.005 | 0.245 | 0.012 | 0.070 | 0.872 | - | - | - | -0.005 | 0.812 | 0.998 | 0.996 | -0.009 | 0.017 | 0.593 | 9 | 566 |
| Plasma | GLA | 0.001 | -0.006 | 0.023 | 0.775 | - | - | - | - | - | - | - | - | - | - | - | - | - | 2 | 293 |
| RBC | GLA | 0.001 | - | - | - | - | - | - | 3.67e-04 | 0.047 | 0.994 | - | - | - | - | - | - | - | 1 | 49 |
| RBC | GLA | 0.01 | - | - | - | - | - | - | 3.67e-04 | 0.047 | 0.994 | - | - | - | - | - | - | - | 1 | 49 |
| RBC | GLA | 0.1 | - | - | - | - | - | - | 3.67e-04 | 0.047 | 0.994 | - | - | - | - | - | - | - | 1 | 49 |
| RBC | GLA | 0.3 | - | - | - | - | - | - | 3.67e-04 | 0.047 | 0.994 | - | - | - | - | - | - | - | 1 | 49 |
| Plasma | DGLA | 0.001 | 0.011 | 0.005 | 0.031 | - | - | - | - | - | - | - | - | - | - | - | - | - | 2 | 1417 |
| RBC | DGLA | 0.001 | 0.010 | 0.004 | 0.007 | - | - | - | - | - | - | - | - | - | - | - | - | - | 2 | 542 |
| RBC | DGLA | 0.01 | 0.007 | 0.006 | 0.284 | 0.017 | 0.026 | 0.579 | - | - | - | -0.006 | 0.702 | 0.786 | 0.647 | 0.009 | 0.012 | 0.438 | 4 | 788 |
| RBC | DGLA | 0.1 | 0.001 | 0.005 | 0.793 | 0.001 | 0.015 | 0.943 | - | - | - | 1.23e-04 | 0.989 | 0.850 | 0.792 | -0.002 | 0.008 | 0.767 | 14 | 2375 |
| RBC | DGLA | 0.3 | 3.29e-04 | 0.003 | 0.896 | 0.002 | 0.010 | 0.854 | - | - | - | -0.001 | 0.869 | 0.997 | 0.996 | -0.002 | 0.005 | 0.702 | 26 | 5970 |
| Plasma | AA | 0.001 | -0.004 | 0.007 | 0.576 | - | - | - | - | - | - | - | - | - | - | - | - | - | 2 | 4616 |
| RBC | AA | 0.001 | 0.016 | 0.008 | 0.050 | -0.021 | 0.063 | 0.793 | - | - | - | 0.015 | 0.648 | 0.821 | 0.914 | 0.013 | 0.020 | 0.527 | 3 | 268 |
| RBC | AA | 0.01 | 0.011 | 0.010 | 0.294 | -0.031 | 0.058 | 0.684 | - | - | - | 0.018 | 0.582 | 0.711 | 0.767 | 0.008 | 0.020 | 0.670 | 3 | 287 |
| RBC | AA | 0.1 | -0.004 | 0.008 | 0.580 | -0.024 | 0.042 | 0.599 | - | - | - | 0.008 | 0.651 | 0.876 | 0.821 | -0.012 | 0.014 | 0.399 | 7 | 584 |
| RBC | AA | 0.3 | 0.001 | 0.004 | 0.754 | -0.002 | 0.024 | 0.946 | - | - | - | 0.001 | 0.899 | 0.991 | 0.983 | 0.002 | 0.010 | 0.873 | 14 | 1440 |
| Plasma | DPA-n3 | 0.001 | -0.005 | 0.009 | 0.579 | -0.027 | 0.041 | 0.628 | - | - | - | 0.007 | 0.650 | 0.809 | 0.828 | -0.007 | 0.020 | 0.716 | 3 | 835 |
| Plasma | DPA-n3 | 0.01 | -0.005 | 0.009 | 0.577 | -0.027 | 0.041 | 0.627 | - | - | - | 0.007 | 0.648 | 0.809 | 0.836 | -0.007 | 0.019 | 0.708 | 3 | 937 |
| Plasma | DPA-n3 | 0.1 | -0.009 | 0.009 | 0.293 | -0.035 | 0.026 | 0.202 | - | - | - | 0.008 | 0.273 | 0.949 | 0.970 | -0.006 | 0.016 | 0.731 | 14 | 1506 |
| Plasma | DPA-n3 | 0.3 | -0.009 | 0.007 | 0.224 | -0.006 | 0.019 | 0.741 | - | - | - | -0.001 | 0.900 | 0.883 | 0.852 | -0.006 | 0.012 | 0.582 | 29 | 3466 |
| RBC | DPA-n3 | 0.001 | -0.014 | 0.001 | 6.05e-136 | - | - | - | - | - | - | - | - | - | - | - | - | - | 2 | 62 |
| RBC | DPA-n3 | 0.01 | -0.014 | 0.001 | 6.05e-136 | - | - | - | - | - | - | - | - | - | - | - | - | - | 2 | 62 |
| RBC | DPA-n3 | 0.1 | -0.014 | 0.001 | 6.05e-136 | - | - | - | - | - | - | - | - | - | - | - | - | - | 2 | 62 |
| RBC | DPA-n3 | 0.3 | -0.014 | 0.001 | 6.05e-136 | - | - | - | - | - | - | - | - | - | - | - | - | - | 2 | 62 |
| RBC | DTA | 0.001 | - | - | - | - | - | - | -0.009 | 0.036 | 0.794 | - | - | - | - | - | - | - | 1 | 80 |
| RBC | DTA | 0.01 | - | - | - | - | - | - | -0.009 | 0.036 | 0.794 | - | - | - | - | - | - | - | 1 | 80 |
| RBC | DTA | 0.1 | - | - | - | - | - | - | -0.009 | 0.036 | 0.794 | - | - | - | - | - | - | - | 1 | 80 |
| RBC | DTA | 0.3 | 0.003 | 0.017 | 0.858 | - | - | - | - | - | - | - | - | - | - | - | - | - | 2 | 114 |
| Plasma | DHA | 0.001 | - | - | - | - | - | - | 0.042 | 0.075 | 0.574 | - | - | - | - | - | - | - | 1 | 65 |
| Plasma | DHA | 0.01 | - | - | - | - | - | - | 0.042 | 0.075 | 0.574 | - | - | - | - | - | - | - | 1 | 64 |
| Plasma | DHA | 0.1 | - | - | - | - | - | - | 0.042 | 0.075 | 0.574 | - | - | - | - | - | - | - | 1 | 64 |
| Plasma | DHA | 0.3 | 0.049 | 0.010 | 1.56e-06 | - | - | - | - | - | - | - | - | - | - | - | - | - | 2 | 95 |

PUFA, polyunsaturated fatty acid; RBC, red blood cell; ALA, α-Linolenic acid; LA: linoleic acid; GLA, γ-Linoleic acid; DGLA, dihomo-γ-linoleic acid; AA, arachidonic acid; DPA-n3, docosapentaenoic acid; DTA, docosatetraenoic acid; DHA, docosahexaenoic acid; β, causal effect size; SE, standard error; IVW_MRE, inverse-variance weighted random-effects model; Egger, MR-Egger; nsnps, number of SNPs retained for this analysis.

**Supplementary Table 10. Forward Mendelian randomization estimates of associations of genetically predicted polyunsaturated fatty acids with COVID-19 severity based on the release 4 HGI A2.**

| | | | IVW_MRE | | | MR-Egger | | | Wald ratio | | | MR-Egger intercept | | Heterogeneity test | | Weighted median | | | | |
|---|---|---|---|---|---|---|---|---|---|---|---|---|---|---|---|---|---|---|---|---|
| **Group name** | **PUFA** | **$R^2$** | **β** | **SE** | ***P*** | **β** | **SE** | ***P*** | **β** | **SE** | ***P*** | **β** | ***P*** | ***P*_IVW** | ***P*_Egger** | **β** | **SE** | ***P*** | **nsnps** | ***F*-statistic** |
| Plasma | ALA | 0.001 | - | - | - | - | - | - | 0.067 | 0.092 | 0.467 | - | - | - | - | - | - | - | 1 | 256 |
| Plasma | ALA | 0.01 | - | - | - | - | - | - | 0.067 | 0.093 | 0.467 | - | - | - | - | - | - | - | 1 | 312 |
| Plasma | ALA | 0.1 | 0.056 | 0.049 | 0.247 | -0.092 | 0.177 | 0.653 | - | - | - | 0.041 | 0.451 | 0.739 | 0.820 | 0.045 | 0.084 | 0.596 | 4 | 436 |
| Plasma | ALA | 0.3 | 0.087 | 0.049 | 0.075 | -0.130 | 0.137 | 0.364 | - | - | - | 0.054 | 0.120 | 0.398 | 0.551 | 0.069 | 0.066 | 0.295 | 13 | 1107 |
| RBC | ALA | 0.001 | - | - | - | - | - | - | -0.070 | 0.173 | 0.687 | - | - | - | - | - | - | - | 1 | 31 |
| RBC | ALA | 0.01 | - | - | - | - | - | - | -0.070 | 0.173 | 0.687 | - | - | - | - | - | - | - | 1 | 31 |
| RBC | ALA | 0.1 | - | - | - | - | - | - | -0.070 | 0.173 | 0.687 | - | - | - | - | - | - | - | 1 | 31 |
| RBC | ALA | 0.3 | - | - | - | - | - | - | -0.070 | 0.173 | 0.687 | - | - | - | - | - | - | - | 1 | 31 |
| Plasma | LA | 0.001 | 0.050 | 0.030 | 0.093 | 0.001 | 0.083 | 0.993 | - | - | - | 0.021 | 0.590 | 0.725 | 0.777 | 0.048 | 0.052 | 0.351 | 3 | 1329 |
| RBC | LA | 0.001 | - | - | - | - | - | - | 0.037 | 0.081 | 0.645 | - | - | - | - | - | - | - | 1 | 133 |
| RBC | LA | 0.01 | - | - | - | - | - | - | 0.037 | 0.081 | 0.645 | - | - | - | - | - | - | - | 1 | 133 |
| RBC | LA | 0.1 | 0.059 | 0.038 | 0.121 | - | - | - | - | - | - | - | - | - | - | - | - | - | 2 | 175 |
| RBC | LA | 0.3 | 0.059 | 0.045 | 0.185 | -0.177 | 0.221 | 0.448 | - | - | - | 0.070 | 0.311 | 0.312 | 0.332 | 0.041 | 0.057 | 0.468 | 9 | 566 |
| Plasma | GLA | 0.001 | -0.051 | 0.102 | 0.614 | - | - | - | - | - | - | - | - | - | - | - | - | - | 2 | 293 |
| RBC | GLA | 0.001 | - | - | - | - | - | - | 0.056 | 0.139 | 0.687 | - | - | - | - | - | - | - | 1 | 49 |
| RBC | GLA | 0.01 | - | - | - | - | - | - | 0.056 | 0.139 | 0.687 | - | - | - | - | - | - | - | 1 | 49 |
| RBC | GLA | 0.1 | - | - | - | - | - | - | 0.056 | 0.139 | 0.687 | - | - | - | - | - | - | - | 1 | 49 |
| RBC | GLA | 0.3 | - | - | - | - | - | - | 0.056 | 0.139 | 0.687 | - | - | - | - | - | - | - | 1 | 49 |
| Plasma | DGLA | 0.001 | 0.057 | 0.024 | 0.016 | - | - | - | - | - | - | - | - | - | - | - | - | - | 2 | 1417 |
| RBC | DGLA | 0.001 | -0.003 | 0.109 | 0.975 | - | - | - | - | - | - | - | - | - | - | - | - | - | 2 | 542 |
| RBC | DGLA | 0.01 | 0.034 | 0.064 | 0.598 | -0.130 | 0.142 | 0.456 | - | - | - | 0.095 | 0.334 | 0.021 | 0.068 | 0.053 | 0.052 | 0.303 | 4 | 788 |
| RBC | DGLA | 0.1 | 0.052 | 0.023 | 0.021 | 0.022 | 0.053 | 0.690 | - | - | - | 0.020 | 0.538 | 0.229 | 0.198 | 0.037 | 0.028 | 0.187 | 14 | 2375 |
| RBC | DGLA | 0.3 | 0.044 | 0.014 | 0.002 | 0.001 | 0.034 | 0.974 | - | - | - | 0.028 | 0.185 | 0.299 | 0.345 | 0.028 | 0.020 | 0.166 | 26 | 5970 |
| Plasma | AA | 0.001 | -0.022 | 0.029 | 0.440 | - | - | - | - | - | - | - | - | - | - | - | - | - | 2 | 4616 |
| RBC | AA | 0.001 | 4.31e-04 | 0.067 | 0.995 | -0.195 | 0.215 | 0.532 | - | - | - | 0.077 | 0.514 | 0.285 | 0.253 | -0.002 | 0.073 | 0.978 | 3 | 268 |
| RBC | AA | 0.01 | 0.005 | 0.067 | 0.937 | -0.146 | 0.223 | 0.630 | - | - | - | 0.063 | 0.601 | 0.275 | 0.193 | 0.005 | 0.068 | 0.939 | 3 | 287 |
| RBC | AA | 0.1 | -0.099 | 0.042 | 0.017 | -0.087 | 0.145 | 0.574 | - | - | - | -0.005 | 0.935 | 0.358 | 0.252 | -0.076 | 0.052 | 0.144 | 7 | 584 |
| RBC | AA | 0.3 | -0.069 | 0.028 | 0.014 | 0.004 | 0.086 | 0.966 | - | - | - | -0.030 | 0.387 | 0.176 | 0.172 | -0.059 | 0.034 | 0.083 | 14 | 1440 |
| Plasma | DPA-n3 | 0.001 | -0.089 | 0.053 | 0.093 | -0.022 | 0.157 | 0.913 | - | - | - | -0.023 | 0.717 | 0.424 | 0.237 | -0.086 | 0.061 | 0.158 | 3 | 835 |
| Plasma | DPA-n3 | 0.01 | -0.090 | 0.053 | 0.089 | -0.024 | 0.157 | 0.903 | - | - | - | -0.022 | 0.725 | 0.430 | 0.238 | -0.087 | 0.059 | 0.141 | 3 | 937 |
| Plasma | DPA-n3 | 0.1 | -0.105 | 0.039 | 0.006 | 0.004 | 0.086 | 0.963 | - | - | - | -0.034 | 0.171 | 0.614 | 0.716 | -0.054 | 0.057 | 0.350 | 14 | 1506 |
| Plasma | DPA-n3 | 0.3 | -0.101 | 0.029 | 0.001 | -0.009 | 0.064 | 0.894 | - | - | - | -0.028 | 0.120 | 0.437 | 0.522 | -0.052 | 0.042 | 0.218 | 29 | 3466 |
| RBC | DPA-n3 | 0.001 | -0.178 | 0.199 | 0.370 | - | - | - | - | - | - | - | - | - | - | - | - | - | 2 | 62 |
| RBC | DPA-n3 | 0.01 | -0.178 | 0.199 | 0.370 | - | - | - | - | - | - | - | - | - | - | - | - | - | 2 | 62 |
| RBC | DPA-n3 | 0.1 | -0.178 | 0.199 | 0.370 | - | - | - | - | - | - | - | - | - | - | - | - | - | 2 | 62 |
| RBC | DPA-n3 | 0.3 | -0.178 | 0.199 | 0.370 | - | - | - | - | - | - | - | - | - | - | - | - | - | 2 | 62 |
| RBC | DTA | 0.001 | - | - | - | - | - | - | -0.052 | 0.108 | 0.632 | - | - | - | - | - | - | - | 1 | 80 |
| RBC | DTA | 0.01 | - | - | - | - | - | - | -0.052 | 0.108 | 0.632 | - | - | - | - | - | - | - | 1 | 80 |
| RBC | DTA | 0.1 | - | - | - | - | - | - | -0.052 | 0.108 | 0.632 | - | - | - | - | - | - | - | 1 | 80 |
| RBC | DTA | 0.3 | -0.091 | 0.068 | 0.182 | - | - | - | - | - | - | - | - | - | - | - | - | - | 2 | 114 |
| Plasma | DHA | 0.001 | - | - | - | - | - | - | 0.491 | 0.217 | 0.024 | - | - | - | - | - | - | - | 1 | 65 |
| Plasma | DHA | 0.01 | - | - | - | - | - | - | 0.490 | 0.217 | 0.024 | - | - | - | - | - | - | - | 1 | 64 |
| Plasma | DHA | 0.1 | - | - | - | - | - | - | 0.490 | 0.217 | 0.024 | - | - | - | - | - | - | - | 1 | 64 |
| Plasma | DHA | 0.3 | 0.401 | 0.164 | 0.014 | - | - | - | - | - | - | - | - | - | - | - | - | - | 2 | 95 |

PUFA, polyunsaturated fatty acid; RBC, red blood cell; ALA, α-Linolenic acid; LA: linoleic acid; GLA, γ-Linoleic acid; DGLA, dihomo-γ-linoleic acid; AA, arachidonic acid; DPA-n3, docosapentaenoic acid; DTA, docosatetraenoic acid; DHA, docosahexaenoic acid; β, causal effect size; SE, standard error; IVW_MRE, inverse-variance weighted random-effects model; Egger, MR-Egger; nsnps, number of SNPs retained for this analysis.

**Supplementary Table 11. Forward Mendelian randomization estimates of associations of genetically predicted polyunsaturated fatty acids with COVID-19 severity based on the release 4 HGI B2.**

| | | | IVW_MRE | | | MR-Egger | | | Wald ratio | | | MR-Egger intercept | | Heterogeneity test | | Weighted median | | | | |
|---|---|---|---|---|---|---|---|---|---|---|---|---|---|---|---|---|---|---|---|---|
| Group name | PUFA | $R^2$ | β | SE | *P* | β | SE | *P* | β | SE | *P* | β | *P* | *P*_IVW | *P*_Egger | β | SE | *P* | nsnps | *F*-statistic |
| Plasma | ALA | 0.001 | - | - | - | - | - | - | 0.105 | 0.076 | 0.166 | - | - | - | - | - | - | - | 1 | 256 |
| Plasma | ALA | 0.01 | - | - | - | - | - | - | 0.106 | 0.076 | 0.166 | - | - | - | - | - | - | - | 1 | 312 |
| Plasma | ALA | 0.1 | 0.136 | 0.082 | 0.096 | -0.040 | 0.201 | 0.859 | - | - | - | 0.050 | 0.436 | 0.124 | 0.141 | 0.130 | 0.071 | 0.067 | 4 | 436 |
| Plasma | ALA | 0.3 | 0.150 | 0.040 | 1.44e-04 | -0.117 | 0.109 | 0.309 | - | - | - | 0.066 | 0.025 | 0.340 | 0.825 | 0.108 | 0.050 | 0.032 | 13 | 1107 |
| RBC | ALA | 0.001 | - | - | - | - | - | - | 0.092 | 0.146 | 0.526 | - | - | - | - | - | - | - | 1 | 31 |
| RBC | ALA | 0.01 | - | - | - | - | - | - | 0.092 | 0.146 | 0.526 | - | - | - | - | - | - | - | 1 | 31 |
| RBC | ALA | 0.1 | - | - | - | - | - | - | 0.092 | 0.146 | 0.526 | - | - | - | - | - | - | - | 1 | 31 |
| RBC | ALA | 0.3 | - | - | - | - | - | - | 0.092 | 0.146 | 0.526 | - | - | - | - | - | - | - | 1 | 31 |
| Plasma | LA | 0.001 | 0.073 | 0.034 | 0.032 | 0.011 | 0.070 | 0.900 | - | - | - | 0.027 | 0.463 | 0.531 | 0.969 | 0.071 | 0.043 | 0.098 | 3 | 1329 |
| RBC | LA | 0.001 | - | - | - | - | - | - | 0.118 | 0.067 | 0.079 | - | - | - | - | - | - | - | 1 | 133 |
| RBC | LA | 0.01 | - | - | - | - | - | - | 0.118 | 0.067 | 0.079 | - | - | - | - | - | - | - | 1 | 133 |
| RBC | LA | 0.1 | 0.138 | 0.035 | 9.47e-05 | - | - | - | - | - | - | - | - | - | - | - | - | - | 2 | 175 |
| RBC | LA | 0.3 | 0.123 | 0.019 | 1.84e-10 | -0.025 | 0.172 | 0.888 | - | - | - | 0.044 | 0.410 | 0.950 | 0.962 | 0.122 | 0.041 | 0.003 | 9 | 566 |
| Plasma | GLA | 0.001 | -0.091 | 0.113 | 0.417 | - | - | - | - | - | - | - | - | - | - | - | - | - | 2 | 293 |
| RBC | GLA | 0.001 | - | - | - | - | - | - | -0.074 | 0.117 | 0.526 | - | - | - | - | - | - | - | 1 | 49 |
| RBC | GLA | 0.01 | - | - | - | - | - | - | -0.074 | 0.117 | 0.526 | - | - | - | - | - | - | - | 1 | 49 |
| RBC | GLA | 0.1 | - | - | - | - | - | - | -0.074 | 0.117 | 0.526 | - | - | - | - | - | - | - | 1 | 49 |
| RBC | GLA | 0.3 | - | - | - | - | - | - | -0.074 | 0.117 | 0.526 | - | - | - | - | - | - | - | 1 | 49 |
| Plasma | DGLA | 0.001 | 0.077 | 0.010 | 8.31e-14 | - | - | - | - | - | - | - | - | - | - | - | - | - | 2 | 1417 |
| RBC | DGLA | 0.001 | 0.048 | 0.044 | 0.271 | - | - | - | - | - | - | - | - | - | - | - | - | - | 2 | 542 |
| RBC | DGLA | 0.01 | 0.056 | 0.026 | 0.029 | -0.039 | 0.070 | 0.637 | - | - | - | 0.055 | 0.279 | 0.472 | 0.837 | 0.032 | 0.032 | 0.316 | 4 | 788 |
| RBC | DGLA | 0.1 | 0.043 | 0.017 | 0.010 | 0.036 | 0.040 | 0.394 | - | - | - | 0.005 | 0.841 | 0.396 | 0.324 | 0.032 | 0.022 | 0.138 | 14 | 2375 |
| RBC | DGLA | 0.3 | 0.051 | 0.010 | 1.07e-07 | 0.023 | 0.026 | 0.388 | - | - | - | 0.018 | 0.263 | 0.736 | 0.758 | 0.046 | 0.016 | 0.003 | 26 | 5970 |
| Plasma | AA | 0.001 | -0.036 | 0.030 | 0.221 | - | - | - | - | - | - | - | - | - | - | - | - | - | 2 | 4616 |
| RBC | AA | 0.001 | -0.080 | 0.041 | 0.053 | -0.099 | 0.188 | 0.692 | - | - | - | 0.008 | 0.933 | 0.481 | 0.229 | -0.054 | 0.060 | 0.369 | 3 | 268 |
| RBC | AA | 0.01 | -0.078 | 0.041 | 0.058 | -0.076 | 0.176 | 0.740 | - | - | - | -0.001 | 0.994 | 0.470 | 0.219 | -0.051 | 0.056 | 0.360 | 3 | 287 |
| RBC | AA | 0.1 | -0.116 | 0.019 | 1.50e-09 | -0.077 | 0.105 | 0.497 | - | - | - | -0.015 | 0.711 | 0.897 | 0.839 | -0.120 | 0.039 | 0.002 | 7 | 584 |
| RBC | AA | 0.3 | -0.084 | 0.013 | 5.54e-11 | 0.005 | 0.060 | 0.940 | - | - | - | -0.037 | 0.149 | 0.961 | 0.994 | -0.068 | 0.026 | 0.008 | 14 | 1440 |
| Plasma | DPA-n3 | 0.001 | -0.082 | 0.024 | 0.001 | -0.108 | 0.106 | 0.495 | - | - | - | 0.009 | 0.831 | 0.769 | 0.502 | -0.086 | 0.050 | 0.088 | 3 | 835 |
| Plasma | DPA-n3 | 0.01 | -0.082 | 0.024 | 0.001 | -0.109 | 0.107 | 0.492 | - | - | - | 0.009 | 0.824 | 0.775 | 0.512 | -0.086 | 0.050 | 0.086 | 3 | 937 |
| Plasma | DPA-n3 | 0.1 | -0.120 | 0.031 | 1.04e-04 | -0.033 | 0.069 | 0.642 | - | - | - | -0.027 | 0.169 | 0.620 | 0.724 | -0.078 | 0.048 | 0.100 | 14 | 1506 |
| Plasma | DPA-n3 | 0.3 | -0.116 | 0.021 | 5.54e-08 | -0.029 | 0.051 | 0.580 | - | - | - | -0.026 | 0.068 | 0.661 | 0.797 | -0.085 | 0.033 | 0.010 | 29 | 3466 |
| RBC | DPA-n3 | 0.001 | -0.162 | 0.028 | 1.19e-08 | - | - | - | - | - | - | - | - | - | - | - | - | - | 2 | 62 |
| RBC | DPA-n3 | 0.01 | -0.162 | 0.028 | 1.19e-08 | - | - | - | - | - | - | - | - | - | - | - | - | - | 2 | 62 |
| RBC | DPA-n3 | 0.1 | -0.162 | 0.028 | 1.19e-08 | - | - | - | - | - | - | - | - | - | - | - | - | - | 2 | 62 |
| RBC | DPA-n3 | 0.3 | -0.162 | 0.028 | 1.19e-08 | - | - | - | - | - | - | - | - | - | - | - | - | - | 2 | 62 |
| RBC | DTA | 0.001 | - | - | - | - | - | - | -0.103 | 0.088 | 0.244 | - | - | - | - | - | - | - | 1 | 80 |
| RBC | DTA | 0.01 | - | - | - | - | - | - | -0.103 | 0.088 | 0.244 | - | - | - | - | - | - | - | 1 | 80 |
| RBC | DTA | 0.1 | - | - | - | - | - | - | -0.103 | 0.088 | 0.244 | - | - | - | - | - | - | - | 1 | 80 |
| RBC | DTA | 0.3 | -0.152 | 0.077 | 0.048 | - | - | - | - | - | - | - | - | - | - | - | - | - | 2 | 114 |
| Plasma | DHA | 0.001 | - | - | - | - | - | - | 0.260 | 0.185 | 0.159 | - | - | - | - | - | - | - | 1 | 65 |
| Plasma | DHA | 0.01 | - | - | - | - | - | - | 0.260 | 0.184 | 0.159 | - | - | - | - | - | - | - | 1 | 64 |
| Plasma | DHA | 0.1 | - | - | - | - | - | - | 0.260 | 0.184 | 0.159 | - | - | - | - | - | - | - | 1 | 64 |
| Plasma | DHA | 0.3 | 0.270 | 0.018 | 1.86e-51 | - | - | - | - | - | - | - | - | - | - | - | - | - | 2 | 95 |

PUFA, polyunsaturated fatty acid; RBC, red blood cell; ALA, α-Linolenic acid; LA: linoleic acid; GLA, γ-Linoleic acid; DGLA, dihomo-γ-linoleic acid; AA, arachidonic acid; DPA-n3, docosapentaenoic acid; DTA, docosatetraenoic acid; DHA, docosahexaenoic acid; β, causal effect size; SE, standard error; IVW_MRE, inverse-variance weighted random-effects model; Egger, MR-Egger; nsnps, number of SNPs retained for this analysis.

**Supplementary Table 12. Forward Mendelian randomization estimates of associations of genetically predicted polyunsaturated fatty acids with COVID-19 severity based on the release 4 HGI B1.**

| | | | IVW_MRE | | | MR-Egger | | | Wald ratio | | | MR-Egger intercept | | Heterogeneity test | | Weighted median | | | | |
|---|---|---|---|---|---|---|---|---|---|---|---|---|---|---|---|---|---|---|---|---|
| **Group name** | **PUFA** | **$R^2$** | **β** | **SE** | ***P*** | **β** | **SE** | ***P*** | **β** | **SE** | ***P*** | **β** | ***P*** | ***P*_IVW** | ***P*_Egger** | **β** | **SE** | ***P*** | **nsnps** | ***F*-statistic** |
| Plasma | ALA | 0.001 | - | - | - | - | - | - | 0.080 | 0.137 | 0.558 | - | - | - | - | - | - | - | 1 | 256 |
| Plasma | ALA | 0.01 | - | - | - | - | - | - | 0.081 | 0.138 | 0.558 | - | - | - | - | - | - | - | 1 | 312 |
| Plasma | ALA | 0.1 | 0.174 | 0.155 | 0.261 | -0.085 | 0.408 | 0.855 | - | - | - | 0.073 | 0.558 | 0.068 | 0.057 | 0.124 | 0.130 | 0.338 | 4 | 436 |
| Plasma | ALA | 0.3 | 0.142 | 0.072 | 0.050 | -0.119 | 0.210 | 0.583 | - | - | - | 0.064 | 0.214 | 0.100 | 0.140 | 0.052 | 0.083 | 0.531 | 13 | 1107 |
| RBC | ALA | 0.001 | - | - | - | - | - | - | 0.125 | 0.248 | 0.615 | - | - | - | - | - | - | - | 1 | 31 |
| RBC | ALA | 0.01 | - | - | - | - | - | - | 0.125 | 0.248 | 0.615 | - | - | - | - | - | - | - | 1 | 31 |
| RBC | ALA | 0.1 | - | - | - | - | - | - | 0.125 | 0.248 | 0.615 | - | - | - | - | - | - | - | 1 | 31 |
| RBC | ALA | 0.3 | - | - | - | - | - | - | 0.125 | 0.248 | 0.615 | - | - | - | - | - | - | - | 1 | 31 |
| Plasma | LA | 0.001 | 0.036 | 0.028 | 0.198 | 0.085 | 0.127 | 0.624 | - | - | - | -0.021 | 0.712 | 0.877 | 0.871 | 0.037 | 0.080 | 0.640 | 3 | 1329 |
| RBC | LA | 0.001 | - | - | - | - | - | - | 0.051 | 0.113 | 0.651 | - | - | - | - | - | - | - | 1 | 133 |
| RBC | LA | 0.01 | - | - | - | - | - | - | 0.051 | 0.113 | 0.651 | - | - | - | - | - | - | - | 1 | 133 |
| RBC | LA | 0.1 | 0.113 | 0.110 | 0.306 | - | - | - | - | - | - | - | - | - | - | - | - | - | 2 | 175 |
| RBC | LA | 0.3 | 0.146 | 0.049 | 0.003 | -0.098 | 0.296 | 0.750 | - | - | - | 0.072 | 0.430 | 0.598 | 0.570 | 0.145 | 0.076 | 0.057 | 9 | 566 |
| Plasma | GLA | 0.001 | -0.103 | 0.017 | 2.42e-09 | - | - | - | - | - | - | - | - | - | - | - | - | - | 2 | 293 |
| RBC | GLA | 0.001 | - | - | - | - | - | - | -0.100 | 0.199 | 0.615 | - | - | - | - | - | - | - | 1 | 49 |
| RBC | GLA | 0.01 | - | - | - | - | - | - | -0.100 | 0.199 | 0.615 | - | - | - | - | - | - | - | 1 | 49 |
| RBC | GLA | 0.1 | - | - | - | - | - | - | -0.100 | 0.199 | 0.615 | - | - | - | - | - | - | - | 1 | 49 |
| RBC | GLA | 0.3 | - | - | - | - | - | - | -0.100 | 0.199 | 0.615 | - | - | - | - | - | - | - | 1 | 49 |
| Plasma | DGLA | 0.001 | 0.025 | 0.048 | 0.595 | - | - | - | - | - | - | - | - | - | - | - | - | - | 2 | 1417 |
| RBC | DGLA | 0.001 | -0.015 | 0.013 | 0.231 | - | - | - | - | - | - | - | - | - | - | - | - | - | 2 | 542 |
| RBC | DGLA | 0.01 | 0.005 | 0.028 | 0.855 | -0.076 | 0.117 | 0.582 | - | - | - | 0.048 | 0.530 | 0.754 | 0.731 | -0.008 | 0.050 | 0.867 | 4 | 788 |
| RBC | DGLA | 0.1 | 0.041 | 0.032 | 0.202 | 0.062 | 0.081 | 0.458 | - | - | - | -0.013 | 0.778 | 0.175 | 0.134 | 0.046 | 0.038 | 0.224 | 14 | 2375 |
| RBC | DGLA | 0.3 | 0.041 | 0.020 | 0.034 | 0.055 | 0.051 | 0.293 | - | - | - | -0.008 | 0.780 | 0.137 | 0.111 | 0.038 | 0.025 | 0.130 | 26 | 5970 |
| Plasma | AA | 0.001 | -0.031 | 0.014 | 0.023 | - | - | - | - | - | - | - | - | - | - | - | - | - | 2 | 4616 |
| RBC | AA | 0.001 | -0.216 | 0.082 | 0.008 | 0.061 | 0.251 | 0.848 | - | - | - | -0.113 | 0.454 | 0.312 | 0.320 | -0.207 | 0.088 | 0.019 | 3 | 268 |
| RBC | AA | 0.01 | -0.218 | 0.080 | 0.006 | 0.055 | 0.235 | 0.854 | - | - | - | -0.114 | 0.435 | 0.326 | 0.393 | -0.209 | 0.088 | 0.017 | 3 | 287 |
| RBC | AA | 0.1 | -0.112 | 0.062 | 0.073 | 0.079 | 0.225 | 0.738 | - | - | - | -0.074 | 0.416 | 0.172 | 0.167 | -0.039 | 0.069 | 0.577 | 7 | 584 |
| RBC | AA | 0.3 | -0.068 | 0.035 | 0.051 | 0.107 | 0.103 | 0.322 | - | - | - | -0.071 | 0.101 | 0.319 | 0.474 | -0.042 | 0.046 | 0.361 | 14 | 1440 |
| Plasma | DPA-n3 | 0.001 | -0.117 | 0.071 | 0.097 | -0.018 | 0.177 | 0.936 | - | - | - | -0.033 | 0.637 | 0.495 | 0.319 | -0.087 | 0.087 | 0.317 | 3 | 835 |
| Plasma | DPA-n3 | 0.01 | -0.118 | 0.071 | 0.093 | -0.021 | 0.177 | 0.926 | - | - | - | -0.032 | 0.643 | 0.501 | 0.320 | -0.089 | 0.093 | 0.340 | 3 | 937 |
| Plasma | DPA-n3 | 0.1 | -0.121 | 0.065 | 0.061 | 0.003 | 0.131 | 0.980 | - | - | - | -0.039 | 0.296 | 0.143 | 0.160 | -0.096 | 0.079 | 0.226 | 14 | 1506 |
| Plasma | DPA-n3 | 0.3 | -0.124 | 0.042 | 0.003 | 0.008 | 0.094 | 0.932 | - | - | - | -0.040 | 0.132 | 0.060 | 0.092 | -0.053 | 0.055 | 0.336 | 29 | 3466 |
| RBC | DPA-n3 | 0.001 | -0.240 | 0.048 | 6.15e-07 | - | - | - | - | - | - | - | - | - | - | - | - | - | 2 | 62 |
| RBC | DPA-n3 | 0.01 | -0.240 | 0.048 | 6.15e-07 | - | - | - | - | - | - | - | - | - | - | - | - | - | 2 | 62 |
| RBC | DPA-n3 | 0.1 | -0.240 | 0.048 | 6.15e-07 | - | - | - | - | - | - | - | - | - | - | - | - | - | 2 | 62 |
| RBC | DPA-n3 | 0.3 | -0.240 | 0.048 | 6.15e-07 | - | - | - | - | - | - | - | - | - | - | - | - | - | 2 | 62 |
| RBC | DTA | 0.001 | - | - | - | - | - | - | -0.089 | 0.154 | 0.563 | - | - | - | - | - | - | - | 1 | 80 |
| RBC | DTA | 0.01 | - | - | - | - | - | - | -0.089 | 0.154 | 0.563 | - | - | - | - | - | - | - | 1 | 80 |
| RBC | DTA | 0.1 | - | - | - | - | - | - | -0.089 | 0.154 | 0.563 | - | - | - | - | - | - | - | 1 | 80 |
| RBC | DTA | 0.3 | -0.232 | 0.190 | 0.223 | - | - | - | - | - | - | - | - | - | - | - | - | - | 2 | 114 |
| Plasma | DHA | 0.001 | - | - | - | - | - | - | 0.468 | 0.329 | 0.155 | - | - | - | - | - | - | - | 1 | 65 |
| Plasma | DHA | 0.01 | - | - | - | - | - | - | 0.467 | 0.328 | 0.155 | - | - | - | - | - | - | - | 1 | 64 |
| Plasma | DHA | 0.1 | - | - | - | - | - | - | 0.467 | 0.328 | 0.155 | - | - | - | - | - | - | - | 1 | 64 |
| Plasma | DHA | 0.3 | 0.362 | 0.145 | 0.013 | - | - | - | - | - | - | - | - | - | - | - | - | - | 2 | 95 |

PUFA, polyunsaturated fatty acid; RBC, red blood cell; ALA, α-Linolenic acid; LA: linoleic acid; GLA, γ-Linoleic acid; DGLA, dihomo-γ-linoleic acid; AA, arachidonic acid; DPA-n3, docosapentaenoic acid; DTA, docosatetraenoic acid; DHA, docosahexaenoic acid; β, causal effect size; SE, standard error; IVW_MRE, inverse-variance weighted random-effects model; Egger, MR-Egger; nsnps, number of SNPs retained for this analysis.

**Supplementary Table 13. Forward Mendelian randomization estimates of associations of genetically predicted polyunsaturated fatty acids with COVID-19 susceptibility based on the release 4 HGI C2.**

| | | | IVW_MRE | | | MR-Egger | | | Wald ratio | | | MR-Egger intercept | | Heterogeneity test | | Weighted median | | | | |
|---|---|---|---|---|---|---|---|---|---|---|---|---|---|---|---|---|---|---|---|---|
| **Group name** | **PUFA** | **$R^2$** | **β** | **SE** | ***P*** | **β** | **SE** | ***P*** | **β** | **SE** | ***P*** | **β** | ***P*** | ***P*_IVW** | ***P*_Egger** | **β** | **SE** | ***P*** | **nsnps** | ***F*-statistic** |
| Plasma | ALA | 0.001 | - | - | - | - | - | - | 0.005 | 0.047 | 0.908 | - | - | - | - | - | - | - | 1 | 256 |
| Plasma | ALA | 0.01 | - | - | - | - | - | - | 0.005 | 0.048 | 0.908 | - | - | - | - | - | - | - | 1 | 312 |
| Plasma | ALA | 0.1 | 0.003 | 0.011 | 0.782 | -0.012 | 0.086 | 0.904 | - | - | - | 0.004 | 0.868 | 0.957 | 0.870 | 0.007 | 0.039 | 0.861 | 4 | 436 |
| Plasma | ALA | 0.3 | -0.003 | 0.015 | 0.811 | -0.019 | 0.063 | 0.775 | - | - | - | 0.004 | 0.805 | 0.937 | 0.905 | -0.013 | 0.028 | 0.643 | 13 | 1107 |
| RBC | ALA | 0.001 | - | - | - | - | - | - | -0.023 | 0.090 | 0.796 | - | - | - | - | - | - | - | 1 | 31 |
| RBC | ALA | 0.01 | - | - | - | - | - | - | -0.023 | 0.090 | 0.796 | - | - | - | - | - | - | - | 1 | 31 |
| RBC | ALA | 0.1 | - | - | - | - | - | - | -0.023 | 0.090 | 0.796 | - | - | - | - | - | - | - | 1 | 31 |
| RBC | ALA | 0.3 | - | - | - | - | - | - | -0.023 | 0.090 | 0.796 | - | - | - | - | - | - | - | 1 | 31 |
| Plasma | LA | 0.001 | 0.009 | 0.028 | 0.740 | -0.029 | 0.044 | 0.625 | - | - | - | 0.017 | 0.465 | 0.326 | 0.318 | 0.003 | 0.027 | 0.900 | 3 | 1329 |
| RBC | LA | 0.001 | - | - | - | - | - | - | -0.003 | 0.042 | 0.949 | - | - | - | - | - | - | - | 1 | 133 |
| RBC | LA | 0.01 | - | - | - | - | - | - | -0.003 | 0.042 | 0.949 | - | - | - | - | - | - | - | 1 | 133 |
| RBC | LA | 0.1 | -0.002 | 0.001 | 0.168 | - | - | - | - | - | - | - | - | - | - | - | - | - | 2 | 175 |
| RBC | LA | 0.3 | -0.009 | 0.014 | 0.518 | 0.016 | 0.106 | 0.885 | - | - | - | -0.007 | 0.816 | 0.835 | 0.758 | -0.016 | 0.025 | 0.527 | 9 | 566 |
| Plasma | GLA | 0.001 | -0.005 | 0.004 | 0.230 | - | - | - | - | - | - | - | - | - | - | - | - | - | 2 | 293 |
| RBC | GLA | 0.001 | - | - | - | - | - | - | 0.019 | 0.072 | 0.796 | - | - | - | - | - | - | - | 1 | 49 |
| RBC | GLA | 0.01 | - | - | - | - | - | - | 0.019 | 0.072 | 0.796 | - | - | - | - | - | - | - | 1 | 49 |
| RBC | GLA | 0.1 | - | - | - | - | - | - | 0.019 | 0.072 | 0.796 | - | - | - | - | - | - | - | 1 | 49 |
| RBC | GLA | 0.3 | - | - | - | - | - | - | 0.019 | 0.072 | 0.796 | - | - | - | - | - | - | - | 1 | 49 |
| Plasma | DGLA | 0.001 | 0.003 | 0.001 | 1.64e-11 | - | - | - | - | - | - | - | - | - | - | - | - | - | 2 | 1417 |
| RBC | DGLA | 0.001 | 0.029 | 0.011 | 0.008 | - | - | - | - | - | - | - | - | - | - | - | - | - | 2 | 542 |
| RBC | DGLA | 0.01 | 0.017 | 0.015 | 0.270 | 0.032 | 0.046 | 0.556 | - | - | - | -0.009 | 0.751 | 0.429 | 0.273 | 0.022 | 0.019 | 0.240 | 4 | 788 |
| RBC | DGLA | 0.1 | 0.003 | 0.009 | 0.742 | -0.004 | 0.023 | 0.853 | - | - | - | 0.005 | 0.727 | 0.507 | 0.436 | -0.015 | 0.013 | 0.254 | 14 | 2375 |
| RBC | DGLA | 0.3 | -4.41e-04 | 0.005 | 0.936 | 0.001 | 0.016 | 0.965 | - | - | - | -0.001 | 0.938 | 0.800 | 0.755 | -0.009 | 0.009 | 0.306 | 26 | 5970 |
| Plasma | AA | 0.001 | -0.002 | 0.001 | 0.047 | - | - | - | - | - | - | - | - | - | - | - | - | - | 2 | 4616 |
| RBC | AA | 0.001 | -0.001 | 0.015 | 0.952 | 0.004 | 0.096 | 0.973 | - | - | - | -0.002 | 0.966 | 0.756 | 0.455 | -0.007 | 0.031 | 0.823 | 3 | 268 |
| RBC | AA | 0.01 | -0.007 | 0.019 | 0.705 | -0.019 | 0.087 | 0.865 | - | - | - | 0.005 | 0.911 | 0.634 | 0.345 | -0.010 | 0.032 | 0.767 | 3 | 287 |
| RBC | AA | 0.1 | -0.004 | 0.017 | 0.796 | -0.011 | 0.064 | 0.873 | - | - | - | 0.002 | 0.921 | 0.542 | 0.416 | 0.013 | 0.025 | 0.607 | 7 | 584 |
| RBC | AA | 0.3 | 0.003 | 0.010 | 0.772 | -0.003 | 0.037 | 0.941 | - | - | - | 0.002 | 0.877 | 0.792 | 0.727 | 0.007 | 0.016 | 0.651 | 14 | 1440 |
| Plasma | DPA-n3 | 0.001 | -0.001 | 0.024 | 0.961 | -0.049 | 0.063 | 0.580 | - | - | - | 0.016 | 0.549 | 0.509 | 0.432 | -0.007 | 0.030 | 0.810 | 3 | 835 |
| Plasma | DPA-n3 | 0.01 | -0.001 | 0.024 | 0.956 | -0.050 | 0.063 | 0.575 | - | - | - | 0.016 | 0.545 | 0.509 | 0.439 | -0.007 | 0.030 | 0.812 | 3 | 937 |
| Plasma | DPA-n3 | 0.1 | -0.005 | 0.015 | 0.717 | -0.036 | 0.040 | 0.384 | - | - | - | 0.010 | 0.392 | 0.902 | 0.905 | -0.006 | 0.026 | 0.834 | 14 | 1506 |
| Plasma | DPA-n3 | 0.3 | 0.001 | 0.010 | 0.937 | 0.010 | 0.030 | 0.736 | - | - | - | -0.003 | 0.729 | 0.955 | 0.943 | 0.017 | 0.018 | 0.358 | 29 | 3466 |
| RBC | DPA-n3 | 0.001 | -0.010 | 0.004 | 0.004 | - | - | - | - | - | - | - | - | - | - | - | - | - | 2 | 62 |
| RBC | DPA-n3 | 0.01 | -0.010 | 0.004 | 0.004 | - | - | - | - | - | - | - | - | - | - | - | - | - | 2 | 62 |
| RBC | DPA-n3 | 0.1 | -0.010 | 0.004 | 0.004 | - | - | - | - | - | - | - | - | - | - | - | - | - | 2 | 62 |
| RBC | DPA-n3 | 0.3 | -0.010 | 0.004 | 0.004 | - | - | - | - | - | - | - | - | - | - | - | - | - | 2 | 62 |
| RBC | DTA | 0.001 | - | - | - | - | - | - | 0.003 | 0.055 | 0.963 | - | - | - | - | - | - | - | 1 | 80 |
| RBC | DTA | 0.01 | - | - | - | - | - | - | 0.003 | 0.055 | 0.963 | - | - | - | - | - | - | - | 1 | 80 |
| RBC | DTA | 0.1 | - | - | - | - | - | - | 0.003 | 0.055 | 0.963 | - | - | - | - | - | - | - | 1 | 80 |
| RBC | DTA | 0.3 | 0.015 | 0.018 | 0.391 | - | - | - | - | - | - | - | - | - | - | - | - | - | 2 | 114 |
| Plasma | DHA | 0.001 | - | - | - | - | - | - | 0.046 | 0.114 | 0.689 | - | - | - | - | - | - | - | 1 | 65 |
| Plasma | DHA | 0.01 | - | - | - | - | - | - | 0.045 | 0.113 | 0.689 | - | - | - | - | - | - | - | 1 | 64 |
| Plasma | DHA | 0.1 | - | - | - | - | - | - | 0.045 | 0.113 | 0.689 | - | - | - | - | - | - | - | 1 | 64 |
| Plasma | DHA | 0.3 | 0.029 | 0.025 | 0.245 | - | - | - | - | - | - | - | - | - | - | - | - | - | 2 | 95 |

PUFA, polyunsaturated fatty acid; RBC, red blood cell; ALA, α-Linolenic acid; LA: linoleic acid; GLA, γ-Linoleic acid; DGLA, dihomo-γ-linoleic acid; AA, arachidonic acid; DPA-n3, docosapentaenoic acid; DTA, docosatetraenoic acid; DHA, docosahexaenoic acid; β, causal effect size; SE, standard error; IVW_MRE, inverse-variance weighted random-effects model; Egger, MR-Egger; nsnps, number of SNPs retained for this analysis.

**Supplementary Table 14. Reverse Mendelian randomization estimates of associations of genetically predicted COVID-19 severity with polyunsaturated fatty acids based on the release 5 HGI A2 (COVID-19 SNP *P* < 5e-8).**

| | | IVW_MRE | | | MR-Egger | | | MR-Egger intercept | | Heterogeneity test | | Weighted median | | | | |
|---|---|---|---|---|---|---|---|---|---|---|---|---|---|---|---|---|
| **Group name** | **PUFA** | **β** | **SE** | ***P*** | **β** | **SE** | ***P*** | **β** | ***P*** | ***P*_IVW** | ***P*_Egger** | **β** | **SE** | ***P*** | **nsnps** | ***F*-statistic** |
| Plasma | ALA | 0.02 | 0.009 | 0.019 | - | - | - | - | - | - | - | - | - | - | 2 | 119 |
| RBC | ALA | 0.008 | 0.009 | 0.37 | 0.019 | 0.023 | 0.43 | -0.004 | 0.596 | 0.544 | 0.464 | 0.013 | 0.012 | 0.31 | 8 | 536 |
| RBC | LA | -0.004 | 0.003 | 0.241 | -0.014 | 0.009 | 0.146 | 0.003 | 0.229 | 0.527 | 0.634 | -0.005 | 0.004 | 0.233 | 8 | 536 |
| RBC | GLA | -0.006 | 0.006 | 0.279 | 0.015 | 0.033 | 0.668 | -0.007 | 0.515 | 0.991 | 0.994 | -0.008 | 0.016 | 0.603 | 8 | 536 |
| RBC | DGLA | -0.004 | 0.004 | 0.29 | -0.01 | 0.012 | 0.444 | 0.002 | 0.601 | 0.802 | 0.744 | -0.004 | 0.006 | 0.494 | 8 | 536 |
| RBC | AA | 0.001 | 0.002 | 0.595 | 0.006 | 0.006 | 0.335 | -0.002 | 0.373 | 0.747 | 0.764 | 0.001 | 0.003 | 0.799 | 8 | 536 |
| Plasma | DPA-n3 | -0.031 | 0.053 | 0.554 | - | - | - | - | - | - | - | - | - | - | 2 | 119 |
| RBC | DPA-n3 | 0.004 | 0.003 | 0.165 | -0.002 | 0.009 | 0.862 | 0.002 | 0.539 | 0.8 | 0.757 | 0.003 | 0.005 | 0.466 | 8 | 536 |
| RBC | DTA | 0.007 | 0.005 | 0.152 | -0.002 | 0.014 | 0.906 | 0.003 | 0.509 | 0.58 | 0.523 | -0.001 | 0.007 | 0.914 | 8 | 536 |
| Plasma | DHA | -0.061 | 0.164 | 0.71 | - | - | - | - | - | - | - | - | - | - | 2 | 119 |

PUFA, polyunsaturated fatty acid; RBC, red blood cell; ALA, α-Linolenic acid; LA: linoleic acid; GLA, γ-Linoleic acid; DGLA, dihomo-γ-linoleic acid; AA, arachidonic acid; DPA-n3, docosapentaenoic acid; DTA, docosatetraenoic acid; DHA, docosahexaenoic acid; β, causal effect size; SE, standard error; IVW_MRE, inverse-variance weighted random-effects model; Egger, MR-Egger; nsnps, number of SNPs retained for this analysis.

**Supplementary Table 15. Reverse Mendelian randomization estimates of associations of genetically predicted COVID-19 severity with polyunsaturated fatty acids based on the release 5 HGI B2 (COVID-19 SNP *P* < 5e-8).**

| | | IVW_MRE | | | MR-Egger | | | MR-Egger intercept | | Heterogeneity test | | Weighted median | | | | |
|---|---|---|---|---|---|---|---|---|---|---|---|---|---|---|---|---|
| **Group name** | **PUFA** | **β** | **SE** | ***P*** | **β** | **SE** | ***P*** | **β** | ***P*** | ***P*_IVW** | ***P*_Egger** | **β** | **SE** | ***P*** | **nsnps** | ***F*-statistic** |
| Plasma | ALA | 0.005 | 0.013 | 0.706 | - | - | - | - | - | - | - | - | - | - | 2 | 94 |
| RBC | ALA | 0.010 | 0.019 | 0.606 | 0.035 | 0.040 | 0.453 | -0.006 | 0.527 | 0.132 | 0.109 | 0.021 | 0.016 | 0.192 | 5 | 438 |
| RBC | LA | -0.005 | 0.005 | 0.302 | -0.016 | 0.011 | 0.239 | 0.002 | 0.350 | 0.405 | 0.426 | -0.008 | 0.006 | 0.208 | 5 | 438 |
| RBC | GLA | -0.017 | 0.010 | 0.109 | 0.019 | 0.042 | 0.672 | -0.008 | 0.392 | 0.901 | 0.996 | -0.019 | 0.025 | 0.439 | 5 | 438 |
| RBC | DGLA | -0.003 | 0.006 | 0.595 | -0.004 | 0.015 | 0.831 | 1.08e-04 | 0.974 | 0.656 | 0.487 | -0.005 | 0.009 | 0.547 | 5 | 438 |
| RBC | AA | 0.001 | 0.003 | 0.806 | 0.003 | 0.008 | 0.752 | -4.23e-04 | 0.800 | 0.457 | 0.314 | 2.77e-05 | 0.004 | 0.995 | 5 | 438 |
| Plasma | DPA-n3 | 0.022 | 0.012 | 0.062 | - | - | - | - | - | - | - | - | - | - | 2 | 94 |
| RBC | DPA-n3 | 0.006 | 0.004 | 0.125 | 0.004 | 0.012 | 0.732 | 4.95e-04 | 0.843 | 0.686 | 0.528 | 0.005 | 0.007 | 0.423 | 5 | 438 |
| RBC | DTA | 0.005 | 0.009 | 0.551 | -0.015 | 0.017 | 0.430 | 0.005 | 0.257 | 0.339 | 0.461 | -0.002 | 0.010 | 0.805 | 5 | 438 |
| Plasma | DHA | -0.107 | 0.044 | 0.014 | - | - | - | - | - | - | - | - | - | - | 2 | 94 |

PUFA, polyunsaturated fatty acid; RBC, red blood cell; ALA, α-Linolenic acid; LA: linoleic acid; GLA, γ-Linoleic acid; DGLA, dihomo-γ-linoleic acid; AA, arachidonic acid; DPA-n3, docosapentaenoic acid; DTA, docosatetraenoic acid; DHA, docosahexaenoic acid; β, causal effect size; SE, standard error; IVW_MRE, inverse-variance weighted random-effects model; Egger, MR-Egger; nsnps, number of SNPs retained for this analysis.

**Supplementary Table 16. Reverse Mendelian randomization estimates of associations of genetically predicted COVID-19 severity with polyunsaturated fatty acids based on the release 5 HGI B1 (COVID-19 SNP *P* < 5e-8).**

| | | Wald ratio | | | | |
|---|---|---|---|---|---|---|
| **Group name** | **PUFA** | **β** | **SE** | ***P*** | **nsnps** | ***F*-statistic** |
| Plasma | ALA | 3.68E-04 | 0.005 | 0.939 | 1 | 34 |
| RBC | ALA | -0.025 | 0.029 | 0.402 | 1 | 34 |
| RBC | LA | -0.012 | 0.011 | 0.289 | 1 | 34 |
| RBC | GLA | 0.022 | 0.043 | 0.604 | 1 | 34 |
| RBC | DGLA | -0.003 | 0.016 | 0.844 | 1 | 34 |
| RBC | AA | 0.012 | 0.007 | 0.1 | 1 | 34 |
| Plasma | DPA-n3 | -0.001 | 0.016 | 0.946 | 1 | 34 |
| RBC | DPA-n3 | -0.005 | 0.012 | 0.7 | 1 | 34 |
| RBC | DTA | 0.01 | 0.018 | 0.562 | 1 | 34 |
| Plasma | DHA | 0.104 | 0.079 | 0.186 | 1 | 34 |

PUFA, polyunsaturated fatty acid; RBC, red blood cell; ALA, α-Linolenic acid; LA: linoleic acid; GLA, γ-Linoleic acid; DGLA, dihomo-γ-linoleic acid; AA, arachidonic acid; DPA-n3, docosapentaenoic acid; DTA, docosatetraenoic acid; DHA, docosahexaenoic acid; β, causal effect size; SE, standard error; IVW_MRE, inverse-variance weighted random-effects model; Egger, MR-Egger; nsnps, number of SNPs retained for this analysis.

**Supplementary Table 17. Reverse Mendelian randomization estimates of associations of genetically predicted COVID-19 susceptibility with polyunsaturated fatty acids based on the release 5 HGI C2 (COVID-19 SNP *P* < 5e-8).**

| | | IVW_MRE | | | MR-Egger | | | MR-Egger intercept | | Heterogeneity test | | Weighted median | | | | |
|---|---|---|---|---|---|---|---|---|---|---|---|---|---|---|---|---|
| **Group name** | **PUFA** | **β** | **SE** | ***P*** | **β** | **SE** | ***P*** | **β** | ***P*** | ***P*_IVW** | ***P*_Egger** | **β** | **SE** | ***P*** | **nsnps** | ***F*-statistic** |
| Plasma | ALA | 0.009 | 0.004 | 0.017 | 0.024 | 0.041 | 0.660 | -0.001 | 0.770 | 0.794 | 0.572 | 0.008 | 0.009 | 0.393 | 3 | 139 |
| RBC | ALA | 0.038 | 0.017 | 0.026 | 0.133 | 0.150 | 0.425 | -0.008 | 0.548 | 0.952 | 0.952 | 0.025 | 0.045 | 0.574 | 6 | 319 |
| RBC | LA | -0.004 | 0.013 | 0.772 | 0.001 | 0.062 | 0.992 | -3.71e-04 | 0.944 | 0.424 | 0.295 | 0.012 | 0.017 | 0.495 | 6 | 319 |
| RBC | GLA | -0.026 | 0.057 | 0.656 | -0.347 | 0.219 | 0.188 | 0.026 | 0.204 | 0.299 | 0.437 | -0.021 | 0.068 | 0.760 | 6 | 319 |
| RBC | DGLA | -0.004 | 0.024 | 0.870 | 0.060 | 0.108 | 0.606 | -0.005 | 0.573 | 0.145 | 0.112 | -0.026 | 0.026 | 0.322 | 6 | 319 |
| RBC | AA | 0.003 | 0.005 | 0.527 | -0.022 | 0.037 | 0.577 | 0.002 | 0.513 | 0.876 | 0.863 | 0.004 | 0.011 | 0.735 | 6 | 319 |
| Plasma | DPA-n3 | -0.025 | 0.025 | 0.328 | -0.202 | 0.137 | 0.380 | 0.014 | 0.414 | 0.400 | 0.749 | -0.037 | 0.030 | 0.226 | 3 | 139 |
| RBC | DPA-n3 | -0.001 | 0.010 | 0.888 | 0.023 | 0.060 | 0.723 | -0.002 | 0.700 | 0.814 | 0.722 | 0.006 | 0.018 | 0.756 | 6 | 319 |
| RBC | DTA | -0.005 | 0.018 | 0.771 | -0.122 | 0.089 | 0.243 | 0.009 | 0.249 | 0.616 | 0.785 | -0.003 | 0.026 | 0.904 | 6 | 319 |
| Plasma | DHA | 0.149 | 0.014 | 6.08e-26 | 0.250 | 0.657 | 0.769 | -0.008 | 0.901 | 0.988 | 0.998 | 0.160 | 0.141 | 0.256 | 3 | 139 |

PUFA, polyunsaturated fatty acid; RBC, red blood cell; ALA, α-Linolenic acid; LA: linoleic acid; GLA, γ-Linoleic acid; DGLA, dihomo-γ-linoleic acid; AA, arachidonic acid; DPA-n3, docosapentaenoic acid; DTA, docosatetraenoic acid; DHA, docosahexaenoic acid; β, causal effect size; SE, standard error; IVW_MRE, inverse-variance weighted random-effects model; Egger, MR-Egger; nsnps, number of SNPs retained for this analysis.

**Supplementary Table 18. Reverse Mendelian randomization estimates of associations of genetically predicted COVID-19 severity with polyunsaturated fatty acids based on the release 5 HGI A2 (COVID-19 SNP *P* < 5e-6).**

| | | IVW_MRE | | | MR-Egger | | | MR-Egger intercept | | Heterogeneity test | | Weighted median | | | | |
|---|---|---|---|---|---|---|---|---|---|---|---|---|---|---|---|---|
| **Group name** | **PUFA** | **β** | **SE** | ***P*** | **β** | **SE** | ***P*** | **β** | ***P*** | ***P*_IVW** | ***P*_Egger** | **β** | **SE** | ***P*** | **nsnps** | ***F*-statistic** |
| Plasma | ALA | 0.002 | 0.003 | 0.568 | -3.09e-04 | 0.011 | 0.979 | 3.44e-04 | 0.868 | 0.302 | 0.216 | 0.002 | 0.003 | 0.583 | 8 | 262 |
| RBC | ALA | 0.010 | 0.006 | 0.099 | 0.007 | 0.017 | 0.696 | 0.001 | 0.833 | 0.653 | 0.605 | 0.014 | 0.010 | 0.150 | 31 | 1078 |
| RBC | LA | -0.004 | 0.002 | 0.057 | -0.006 | 0.006 | 0.360 | 4.00e-04 | 0.775 | 0.722 | 0.679 | -0.005 | 0.004 | 0.187 | 31 | 1078 |
| RBC | GLA | -0.015 | 0.007 | 0.050 | 0.011 | 0.025 | 0.648 | -0.006 | 0.265 | 0.948 | 0.957 | -0.015 | 0.014 | 0.276 | 31 | 1078 |
| RBC | DGLA | -0.002 | 0.003 | 0.459 | -0.004 | 0.009 | 0.651 | 4.09e-04 | 0.836 | 0.666 | 0.618 | -0.004 | 0.005 | 0.422 | 31 | 1078 |
| RBC | AA | 0.002 | 0.001 | 0.139 | 0.003 | 0.004 | 0.538 | -1.56e-04 | 0.865 | 0.907 | 0.883 | 0.003 | 0.002 | 0.292 | 31 | 1078 |
| Plasma | DPA-n3 | -0.002 | 0.011 | 0.822 | -0.016 | 0.045 | 0.736 | 0.002 | 0.768 | 0.075 | 0.049 | 0.008 | 0.012 | 0.533 | 8 | 262 |
| RBC | DPA-n3 | 2.17e-04 | 0.003 | 0.933 | 0.002 | 0.007 | 0.763 | -4.42e-04 | 0.770 | 0.479 | 0.432 | 0.002 | 0.004 | 0.567 | 31 | 1078 |
| RBC | DTA | 0.004 | 0.004 | 0.349 | 0.011 | 0.010 | 0.302 | -0.002 | 0.455 | 0.521 | 0.499 | 0.003 | 0.006 | 0.645 | 31 | 1078 |
| Plasma | DHA | -0.005 | 0.029 | 0.862 | 0.034 | 0.141 | 0.818 | -0.007 | 0.783 | 0.769 | 0.675 | -0.022 | 0.049 | 0.657 | 8 | 262 |

PUFA, polyunsaturated fatty acid; RBC, red blood cell; ALA, α-Linolenic acid; LA: linoleic acid; GLA, γ-Linoleic acid; DGLA, dihomo-γ-linoleic acid; AA, arachidonic acid; DPA-n3, docosapentaenoic acid; DTA, docosatetraenoic acid; DHA, docosahexaenoic acid; β, causal effect size; SE, standard error; IVW_MRE, inverse-variance weighted random-effects model; Egger, MR-Egger; nsnps, number of SNPs retained for this analysis.

**Supplementary Table 19. Reverse Mendelian randomization estimates of associations of genetically predicted COVID-19 severity with polyunsaturated fatty acids based on the release 5 HGI B2 (COVID-19 SNP *P* < 5e-6).**

| | | IVW_MRE | | | MR-Egger | | | MR-Egger intercept | | Heterogeneity test | | Weighted median | | | | |
|---|---|---|---|---|---|---|---|---|---|---|---|---|---|---|---|---|
| **Group name** | **PUFA** | **β** | **SE** | ***P*** | **β** | **SE** | ***P*** | **β** | ***P*** | ***P*_IVW** | ***P*_Egger** | **β** | **SE** | ***P*** | **nsnps** | ***F*-statistic** |
| Plasma | ALA | 0.002 | 0.003 | 0.458 | -0.006 | 0.014 | 0.688 | 0.001 | 0.556 | 0.190 | 0.160 | 0.002 | 0.004 | 0.613 | 13 | 350 |
| RBC | ALA | 0.006 | 0.008 | 0.514 | 0.010 | 0.022 | 0.653 | -0.001 | 0.823 | 0.832 | 0.794 | 0.018 | 0.015 | 0.215 | 27 | 960 |
| RBC | LA | -0.002 | 0.003 | 0.560 | -0.012 | 0.008 | 0.175 | 0.002 | 0.193 | 0.948 | 0.967 | -0.007 | 0.005 | 0.194 | 27 | 960 |
| RBC | GLA | 3.35e-04 | 0.012 | 0.977 | 0.027 | 0.032 | 0.399 | -0.005 | 0.352 | 0.900 | 0.903 | -0.004 | 0.022 | 0.867 | 27 | 960 |
| RBC | DGLA | 0.003 | 0.006 | 0.633 | -0.002 | 0.013 | 0.887 | 0.001 | 0.694 | 0.179 | 0.152 | -0.006 | 0.008 | 0.482 | 27 | 960 |
| RBC | AA | -0.002 | 0.003 | 0.442 | 0.005 | 0.006 | 0.360 | -0.001 | 0.162 | 0.216 | 0.266 | 0.001 | 0.004 | 0.813 | 27 | 960 |
| Plasma | DPA-n3 | -0.002 | 0.008 | 0.765 | -0.053 | 0.039 | 0.201 | 0.006 | 0.207 | 0.777 | 0.852 | -0.002 | 0.013 | 0.898 | 13 | 350 |
| RBC | DPA-n3 | 0.004 | 0.005 | 0.403 | 0.001 | 0.010 | 0.961 | 0.001 | 0.723 | 0.108 | 0.089 | 0.005 | 0.006 | 0.456 | 27 | 960 |
| RBC | DTA | -0.002 | 0.006 | 0.684 | 2.81e-05 | 0.013 | 0.998 | -4.08e-04 | 0.840 | 0.497 | 0.443 | -0.003 | 0.009 | 0.718 | 27 | 960 |
| Plasma | DHA | -0.033 | 0.033 | 0.314 | -0.073 | 0.193 | 0.712 | 0.005 | 0.836 | 0.880 | 0.831 | -0.033 | 0.058 | 0.573 | 13 | 350 |

PUFA, polyunsaturated fatty acid; RBC, red blood cell; ALA, α-Linolenic acid; LA: linoleic acid; GLA, γ-Linoleic acid; DGLA, dihomo-γ-linoleic acid; AA, arachidonic acid; DPA-n3, docosapentaenoic acid; DTA, docosatetraenoic acid; DHA, docosahexaenoic acid; β, causal effect size; SE, standard error; IVW_MRE, inverse-variance weighted random-effects model; Egger, MR-Egger; nsnps, number of SNPs retained for this analysis.

**Supplementary Table 20. Reverse Mendelian randomization estimates of associations of genetically predicted COVID-19 severity with polyunsaturated fatty acids based on the release 5 HGI B1 (COVID-19 SNP *P* < 5e-6).**

| | | IVW_MRE | | | MR-Egger | | | MR-Egger intercept | | Heterogeneity test | | Weighted median | | | | |
|---|---|---|---|---|---|---|---|---|---|---|---|---|---|---|---|---|
| **Group name** | **PUFA** | **β** | **SE** | ***P*** | **β** | **SE** | ***P*** | **β** | ***P*** | ***P*_IVW** | ***P*_Egger** | **β** | **SE** | ***P*** | **nsnps** | ***F*-statistic** |
| Plasma | ALA | 0.003 | 0.002 | 0.162 | -0.004 | 0.014 | 0.810 | 0.002 | 0.679 | 0.674 | 0.486 | 0.003 | 0.004 | 0.440 | 3 | 81 |
| RBC | ALA | 0.009 | 0.014 | 0.529 | 0.046 | 0.038 | 0.244 | -0.011 | 0.306 | 0.068 | 0.080 | 1.97e-04 | 0.016 | 0.990 | 13 | 317 |
| RBC | LA | -0.001 | 0.004 | 0.891 | -0.010 | 0.011 | 0.404 | 0.003 | 0.397 | 0.451 | 0.431 | -0.002 | 0.006 | 0.698 | 13 | 317 |
| RBC | GLA | 0.025 | 0.015 | 0.097 | 0.032 | 0.043 | 0.467 | -0.002 | 0.850 | 0.598 | 0.516 | 0.022 | 0.022 | 0.299 | 13 | 317 |
| RBC | DGLA | -0.010 | 0.004 | 0.014 | -0.017 | 0.016 | 0.293 | 0.002 | 0.610 | 0.947 | 0.929 | -0.006 | 0.007 | 0.395 | 13 | 317 |
| RBC | AA | -0.001 | 0.003 | 0.753 | -0.005 | 0.009 | 0.629 | 0.001 | 0.687 | 0.091 | 0.068 | -0.001 | 0.004 | 0.744 | 13 | 317 |
| Plasma | DPA-n3 | -0.004 | 0.007 | 0.566 | -0.015 | 0.047 | 0.809 | 0.002 | 0.857 | 0.696 | 0.412 | -0.002 | 0.014 | 0.856 | 3 | 81 |
| RBC | DPA-n3 | 0.005 | 0.003 | 0.147 | -0.001 | 0.012 | 0.948 | 0.002 | 0.615 | 0.866 | 0.831 | 0.005 | 0.006 | 0.388 | 13 | 317 |
| RBC | DTA | -0.009 | 0.009 | 0.340 | 0.004 | 0.025 | 0.862 | -0.004 | 0.582 | 0.035 | 0.028 | -2.75e-05 | 0.010 | 0.998 | 13 | 317 |
| Plasma | DHA | 0.048 | 0.051 | 0.345 | 0.210 | 0.237 | 0.539 | -0.035 | 0.609 | 0.482 | 0.327 | 0.050 | 0.066 | 0.453 | 3 | 81 |

PUFA, polyunsaturated fatty acid; RBC, red blood cell; ALA, α-Linolenic acid; LA: linoleic acid; GLA, γ-Linoleic acid; DGLA, dihomo-γ-linoleic acid; AA, arachidonic acid; DPA-n3, docosapentaenoic acid; DTA, docosatetraenoic acid; DHA, docosahexaenoic acid; β, causal effect size; SE, standard error; IVW_MRE, inverse-variance weighted random-effects model; Egger, MR-Egger; nsnps, number of SNPs retained for this analysis.

**Supplementary Table 21. Reverse Mendelian randomization estimates of associations of genetically predicted COVID-19 susceptibility with polyunsaturated fatty acids based on the release 5 HGI C2 (COVID-19 SNP *P* < 5e-6).**

| | | IVW_MRE | | | MR-Egger | | | MR-Egger intercept | | Heterogeneity test | | Weighted median | | | | |
|---|---|---|---|---|---|---|---|---|---|---|---|---|---|---|---|---|
| **Group name** | **PUFA** | **β** | **SE** | ***P*** | **β** | **SE** | ***P*** | **β** | ***P*** | ***P*_IVW** | ***P*_Egger** | **β** | **SE** | ***P*** | **nsnps** | ***F*-statistic** |
| Plasma | ALA | -0.003 | 0.009 | 0.759 | 0.046 | 0.037 | 0.262 | -0.004 | 0.227 | 0.060 | 0.108 | 0.004 | 0.009 | 0.634 | 8 | 268 |
| RBC | ALA | 0.059 | 0.024 | 0.015 | 0.084 | 0.087 | 0.343 | -0.002 | 0.760 | 0.304 | 0.261 | 0.053 | 0.032 | 0.103 | 26 | 792 |
| RBC | LA | -0.004 | 0.007 | 0.572 | -0.041 | 0.030 | 0.185 | 0.003 | 0.212 | 0.863 | 0.894 | 0.001 | 0.012 | 0.952 | 26 | 792 |
| RBC | GLA | -0.008 | 0.034 | 0.826 | -0.116 | 0.122 | 0.352 | 0.008 | 0.365 | 0.377 | 0.370 | -0.047 | 0.048 | 0.323 | 26 | 792 |
| RBC | DGLA | -0.011 | 0.015 | 0.451 | -0.057 | 0.053 | 0.293 | 0.003 | 0.378 | 0.044 | 0.044 | -0.003 | 0.019 | 0.868 | 26 | 792 |
| RBC | AA | 1.24e-04 | 0.005 | 0.980 | 0.001 | 0.020 | 0.952 | -7.98e-05 | 0.955 | 0.761 | 0.712 | 0.001 | 0.008 | 0.910 | 26 | 792 |
| Plasma | DPA-n3 | -0.030 | 0.030 | 0.310 | -0.056 | 0.144 | 0.712 | 0.002 | 0.862 | 0.054 | 0.032 | -0.033 | 0.028 | 0.236 | 8 | 268 |
| RBC | DPA-n3 | 4.15e-04 | 0.009 | 0.963 | 0.033 | 0.032 | 0.316 | -0.002 | 0.303 | 0.511 | 0.518 | 0.004 | 0.013 | 0.744 | 26 | 792 |
| RBC | DTA | 0.011 | 0.013 | 0.378 | -0.037 | 0.048 | 0.446 | 0.004 | 0.302 | 0.573 | 0.582 | 0.001 | 0.019 | 0.945 | 26 | 792 |
| Plasma | DHA | -0.074 | 0.160 | 0.645 | 0.429 | 0.739 | 0.583 | -0.036 | 0.511 | 0.014 | 0.012 | 0.132 | 0.127 | 0.300 | 8 | 268 |

PUFA, polyunsaturated fatty acid; RBC, red blood cell; ALA, α-Linolenic acid; LA: linoleic acid; GLA, γ-Linoleic acid; DGLA, dihomo-γ-linoleic acid; AA, arachidonic acid; DPA-n3, docosapentaenoic acid; DTA, docosatetraenoic acid; DHA, docosahexaenoic acid; β, causal effect size; SE, standard error; IVW_MRE, inverse-variance weighted random-effects model; Egger, MR-Egger; nsnps, number of SNPs retained for this analysis.

**Supplementary Table 22. Reverse Mendelian randomization estimates of associations of genetically predicted COVID-19 severity with polyunsaturated fatty acids based on the release 4 HGI A2 (COVID-19 SNP *P* < 5e-8).**

| | | IVW_MRE | | | MR-Egger | | | MR-Egger intercept | | Heterogeneity test | | Weighted median | | | | |
|---|---|---|---|---|---|---|---|---|---|---|---|---|---|---|---|---|
| **Group name** | **PUFA** | **β** | **SE** | ***P*** | **β** | **SE** | ***P*** | **β** | ***P*** | ***P*_IVW** | ***P*_Egger** | **β** | **SE** | ***P*** | **nsnps** | ***F*-statistic** |
| Plasma | ALA | 0.001 | 0.006 | 0.874 | 0.125 | 0.071 | 0.331 | -0.024 | 0.332 | 0.134 | 0.318 | -0.002 | 0.005 | 0.677 | 3 | 135 |
| RBC | ALA | 0.008 | 0.009 | 0.383 | 0.014 | 0.022 | 0.558 | -0.002 | 0.781 | 0.449 | 0.339 | 0.011 | 0.012 | 0.376 | 7 | 487 |
| RBC | LA | -0.003 | 0.003 | 0.406 | -0.011 | 0.008 | 0.226 | 0.003 | 0.301 | 0.514 | 0.564 | -0.005 | 0.004 | 0.200 | 7 | 487 |
| RBC | GLA | -0.006 | 0.006 | 0.350 | 0.014 | 0.030 | 0.650 | -0.006 | 0.483 | 0.976 | 0.985 | -0.009 | 0.016 | 0.592 | 7 | 487 |
| RBC | DGLA | -0.003 | 0.004 | 0.338 | -0.009 | 0.011 | 0.455 | 0.002 | 0.603 | 0.785 | 0.719 | -0.004 | 0.006 | 0.500 | 7 | 487 |
| RBC | AA | 1.46e-04 | 0.001 | 0.918 | 0.004 | 0.005 | 0.479 | -0.001 | 0.446 | 0.887 | 0.895 | 0.001 | 0.003 | 0.792 | 7 | 487 |
| Plasma | DPA-n3 | 0.009 | 0.018 | 0.626 | -0.293 | 0.296 | 0.503 | 0.058 | 0.493 | 0.230 | 0.231 | 0.012 | 0.015 | 0.432 | 3 | 135 |
| RBC | DPA-n3 | 0.005 | 0.002 | 0.068 | 0.001 | 0.008 | 0.894 | 0.001 | 0.663 | 0.851 | 0.785 | 0.003 | 0.005 | 0.462 | 7 | 487 |
| RBC | DTA | 0.004 | 0.004 | 0.340 | -0.007 | 0.012 | 0.587 | 0.004 | 0.346 | 0.699 | 0.738 | -0.001 | 0.007 | 0.847 | 7 | 487 |
| Plasma | DHA | -0.091 | 0.066 | 0.166 | -0.032 | 1.141 | 0.982 | -0.012 | 0.967 | 0.380 | 0.165 | -0.080 | 0.070 | 0.254 | 3 | 135 |

PUFA, polyunsaturated fatty acid; RBC, red blood cell; ALA, α-Linolenic acid; LA: linoleic acid; GLA, γ-Linoleic acid; DGLA, dihomo-γ-linoleic acid; AA, arachidonic acid; DPA-n3, docosapentaenoic acid; DTA, docosatetraenoic acid; DHA, docosahexaenoic acid; β, causal effect size; SE, standard error; IVW_MRE, inverse-variance weighted random-effects model; Egger, MR-Egger; nsnps, number of SNPs retained for this analysis.

**Supplementary Table 23. Reverse Mendelian randomization estimates of associations of genetically predicted COVID-19 severity with polyunsaturated fatty acids based on the release 4 HGI B2 (COVID-19 SNP *P* < 5e-8).**

| | | IVW_MRE | | | MR-Egger | | | Wald ratio | | | MR-Egger intercept | | Heterogeneity test | | Weighted median | | | | |
|---|---|---|---|---|---|---|---|---|---|---|---|---|---|---|---|---|---|---|---|
| **Group name** | **PUFA** | **β** | **SE** | **P** | **β** | **SE** | **P** | **β** | **SE** | **P** | **β** | **P** | **P_IVW** | **P_Egger** | **β** | **SE** | **P** | **nsnps** | **F-statistic** |
| Plasma | ALA | - | - | - | - | - | - | 0.03 | 0.015 | 0.049 | - | - | - | - | - | - | - | 1 | 61 |
| RBC | ALA | 0.005 | 0.013 | 0.688 | 0.022 | 0.03 | 0.501 | - | - | - | -0.005 | 0.558 | 0.197 | 0.155 | 0.013 | 0.014 | 0.333 | 6 | 462 |
| RBC | LA | -0.003 | 0.004 | 0.366 | -0.011 | 0.009 | 0.29 | - | - | - | 0.002 | 0.402 | 0.498 | 0.48 | -0.006 | 0.005 | 0.224 | 6 | 462 |
| RBC | GLA | -0.01 | 0.009 | 0.279 | 0.018 | 0.034 | 0.625 | - | - | - | -0.008 | 0.409 | 0.891 | 0.934 | -0.012 | 0.019 | 0.542 | 6 | 462 |
| RBC | DGLA | -0.005 | 0.005 | 0.349 | -0.004 | 0.012 | 0.737 | - | - | - | -8.21E-05 | 0.98 | 0.548 | 0.405 | -0.004 | 0.007 | 0.546 | 6 | 462 |
| RBC | AA | 0.001 | 0.002 | 0.818 | 0.002 | 0.006 | 0.734 | - | - | - | -4.40E-04 | 0.774 | 0.599 | 0.468 | 3.89E-04 | 0.003 | 0.905 | 6 | 462 |
| Plasma | DPA-n3 | - | - | - | - | - | - | -0.009 | 0.055 | 0.864 | - | - | - | - | - | - | - | 1 | 61 |
| RBC | DPA-n3 | 0.005 | 0.003 | 0.079 | 0.003 | 0.009 | 0.792 | - | - | - | 0.001 | 0.767 | 0.786 | 0.675 | 0.004 | 0.005 | 0.429 | 6 | 462 |
| RBC | DTA | 0.005 | 0.006 | 0.422 | -0.013 | 0.014 | 0.417 | - | - | - | 0.005 | 0.23 | 0.464 | 0.624 | -0.002 | 0.008 | 0.826 | 6 | 462 |
| Plasma | DHA | - | - | - | - | - | - | -0.027 | 0.155 | 0.862 | - | - | - | - | - | - | - | 1 | 61 |

PUFA, polyunsaturated fatty acid; RBC, red blood cell; ALA, α-Linolenic acid; LA: linoleic acid; GLA, γ-Linoleic acid; DGLA, dihomo-γ-linoleic acid; AA, arachidonic acid; DPA-n3, docosapentaenoic acid; DTA, docosatetraenoic acid; DHA, docosahexaenoic acid; β, causal effect size; SE, standard error; IVW_MRE, inverse-variance weighted random-effects model; Egger, MR-Egger; nsnps, number of SNPs retained for this analysis.

**Supplementary Table 24. Reverse Mendelian randomization estimates of associations of genetically predicted COVID-19 severity with polyunsaturated fatty acids based on the release 4 HGI B1 (COVID-19 SNP *P* < 5e-8).**

| | | Wald ratio | | | | |
|---|---|---|---|---|---|---|
| **Group name** | **PUFA** | **β** | **SE** | ***P*** | **nsnps** | ***F*-statistic** |
| RBC | ALA | 0.004 | 0.023 | 0.874 | 1 | 31 |
| RBC | LA | -0.008 | 0.009 | 0.347 | 1 | 31 |
| RBC | GLA | 0.02 | 0.033 | 0.545 | 1 | 31 |
| RBC | DGLA | 0.001 | 0.012 | 0.91 | 1 | 31 |
| RBC | AA | 0.006 | 0.006 | 0.299 | 1 | 31 |
| RBC | DPA-n3 | -0.003 | 0.009 | 0.771 | 1 | 31 |
| RBC | DTA | 0.006 | 0.014 | 0.635 | 1 | 31 |

PUFA, polyunsaturated fatty acid; RBC, red blood cell; ALA, α-Linolenic acid; LA: linoleic acid; GLA, γ-Linoleic acid; DGLA, dihomo-γ-linoleic acid; AA, arachidonic acid; DPA-n3, docosapentaenoic acid; DTA, docosatetraenoic acid; DHA, docosahexaenoic acid; β, causal effect size; SE, standard error; IVW_MRE, inverse-variance weighted random-effects model; Egger, MR-Egger; nsnps, number of SNPs retained for this analysis.

**Supplementary Table 25. Reverse Mendelian randomization estimates of associations of genetically predicted COVID-19 susceptibility with polyunsaturated fatty acids based on the release 4 HGI C2 (COVID-19 SNP *P* < 5e-8).**

| | | IVW_MRE | | | MR-Egger | | | Wald ratio | | | MR-Egger intercept | | Heterogeneity test | | Weighted median | | | | |
|---|---|---|---|---|---|---|---|---|---|---|---|---|---|---|---|---|---|---|---|
| **Group name** | **PUFA** | **β** | **SE** | **P** | **β** | **SE** | **P** | **β** | **SE** | **P** | **β** | **P** | **P_IVW** | **P_Egger** | **β** | **SE** | **P** | **nsnps** | **F-statistic** |
| Plasma | ALA | - | - | - | - | - | - | 0.066 | 0.034 | 0.049 | - | - | - | - | - | - | - | 1 | 31 |
| RBC | ALA | 0.034 | 0.018 | 0.057 | -0.005 | 0.059 | 0.942 | - | - | - | 0.008 | 0.583 | 0.686 | 0.686 | 0.034 | 0.032 | 0.292 | 3 | 183 |
| RBC | LA | -0.014 | 0.013 | 0.289 | -0.020 | 0.037 | 0.687 | - | - | - | 0.001 | 0.885 | 0.238 | 0.096 | -0.018 | 0.012 | 0.129 | 3 | 183 |
| RBC | GLA | -0.022 | 0.024 | 0.352 | 0.001 | 0.087 | 0.995 | - | - | - | -0.004 | 0.810 | 0.727 | 0.461 | -0.017 | 0.046 | 0.709 | 3 | 183 |
| RBC | DGLA | -0.011 | 0.026 | 0.666 | -0.039 | 0.066 | 0.660 | - | - | - | 0.005 | 0.712 | 0.066 | 0.036 | -0.005 | 0.016 | 0.757 | 3 | 183 |
| RBC | AA | 0.001 | 0.007 | 0.881 | 0.010 | 0.018 | 0.682 | - | - | - | -0.002 | 0.675 | 0.388 | 0.230 | 0.002 | 0.008 | 0.756 | 3 | 183 |
| Plasma | DPA-n3 | - | - | - | - | - | - | -0.021 | 0.121 | 0.864 | - | - | - | - | - | - | - | 1 | 31 |
| RBC | DPA-n3 | 0.012 | 0.009 | 0.176 | -0.002 | 0.024 | 0.941 | - | - | - | 0.003 | 0.623 | 0.582 | 0.427 | 0.011 | 0.012 | 0.380 | 3 | 183 |
| RBC | DTA | -0.005 | 0.039 | 0.891 | -0.055 | 0.096 | 0.670 | - | - | - | 0.009 | 0.660 | 0.007 | 0.007 | 0.002 | 0.018 | 0.901 | 3 | 183 |
| Plasma | DHA | - | - | - | - | - | - | -0.059 | 0.337 | 0.862 | - | - | - | - | - | - | - | 1 | 31 |

PUFA, polyunsaturated fatty acid; RBC, red blood cell; ALA, α-Linolenic acid; LA: linoleic acid; GLA, γ-Linoleic acid; DGLA, dihomo-γ-linoleic acid; AA, arachidonic acid; DPA-n3, docosapentaenoic acid; DTA, docosatetraenoic acid; DHA, docosahexaenoic acid; β, causal effect size; SE, standard error; IVW_MRE, inverse-variance weighted random-effects model; Egger, MR-Egger; nsnps, number of SNPs retained for this analysis.

**Supplementary Table 26. Reverse Mendelian randomization estimates of associations of genetically predicted COVID-19 severity with polyunsaturated fatty acids based on the release 4 HGI A2 (COVID-19 SNP *P* < 5e-6).**

| | | IVW_MRE | | | MR-Egger | | | MR-Egger intercept | | Heterogeneity test | | Weighted median | | | | |
|---|---|---|---|---|---|---|---|---|---|---|---|---|---|---|---|---|
| **Group name** | **PUFA** | **β** | **SE** | ***P*** | **β** | **SE** | ***P*** | **β** | ***P*** | ***P*_IVW** | ***P*_Egger** | **β** | **SE** | ***P*** | **nsnps** | ***F*-statistic** |
| Plasma | ALA | 4.31e-04 | 0.004 | 0.909 | 0.019 | 0.024 | 0.464 | -0.003 | 0.468 | 0.120 | 0.110 | -0.001 | 0.004 | 0.685 | 6 | 202 |
| RBC | ALA | 0.012 | 0.007 | 0.080 | -0.007 | 0.014 | 0.634 | 0.005 | 0.153 | 0.431 | 0.498 | 0.014 | 0.011 | 0.184 | 25 | 905 |
| RBC | LA | -0.004 | 0.003 | 0.085 | -0.006 | 0.005 | 0.279 | 4.86e-04 | 0.721 | 0.471 | 0.420 | -0.006 | 0.004 | 0.126 | 25 | 905 |
| RBC | GLA | -0.016 | 0.010 | 0.100 | -0.011 | 0.021 | 0.609 | -0.001 | 0.794 | 0.470 | 0.416 | -0.006 | 0.014 | 0.692 | 25 | 905 |
| RBC | DGLA | -0.001 | 0.004 | 0.809 | 1.97e-04 | 0.008 | 0.981 | -3.03e-04 | 0.880 | 0.346 | 0.296 | -0.004 | 0.006 | 0.456 | 25 | 905 |
| RBC | AA | 0.001 | 0.001 | 0.419 | 0.001 | 0.004 | 0.848 | 1.20e-04 | 0.891 | 0.849 | 0.810 | 0.002 | 0.002 | 0.454 | 25 | 905 |
| Plasma | DPA-n3 | 0.007 | 0.009 | 0.446 | -0.016 | 0.064 | 0.816 | 0.004 | 0.732 | 0.404 | 0.294 | 0.008 | 0.011 | 0.467 | 6 | 202 |
| RBC | DPA-n3 | 0.004 | 0.003 | 0.155 | 0.005 | 0.006 | 0.367 | -4.37e-04 | 0.763 | 0.486 | 0.433 | 0.003 | 0.004 | 0.404 | 25 | 905 |
| RBC | DTA | 0.004 | 0.004 | 0.366 | -0.005 | 0.009 | 0.604 | 0.002 | 0.294 | 0.257 | 0.268 | 0.003 | 0.006 | 0.623 | 25 | 905 |
| Plasma | DHA | -0.081 | 0.064 | 0.204 | -0.155 | 0.403 | 0.720 | 0.012 | 0.860 | 0.067 | 0.037 | -0.087 | 0.060 | 0.146 | 6 | 202 |

PUFA, polyunsaturated fatty acid; RBC, red blood cell; ALA, α-Linolenic acid; LA: linoleic acid; GLA, γ-Linoleic acid; DGLA, dihomo-γ-linoleic acid; AA, arachidonic acid; DPA-n3, docosapentaenoic acid; DTA, docosatetraenoic acid; DHA, docosahexaenoic acid; β, causal effect size; SE, standard error; IVW_MRE, inverse-variance weighted random-effects model; Egger, MR-Egger; nsnps, number of SNPs retained for this analysis.

**Supplementary Table 27. Reverse Mendelian randomization estimates of associations of genetically predicted COVID-19 severity with polyunsaturated fatty acids based on the release 4 HGI B2 (COVID-19 SNP *P* < 5e-6).**

| | | IVW_MRE | | | MR-Egger | | | MR-Egger intercept | | Heterogeneity test | | Weighted median | | | | |
|---|---|---|---|---|---|---|---|---|---|---|---|---|---|---|---|---|
| **Group name** | **PUFA** | **β** | **SE** | ***P*** | **β** | **SE** | ***P*** | **β** | ***P*** | ***P*_IVW** | ***P*_Egger** | **β** | **SE** | ***P*** | **nsnps** | ***F*-statistic** |
| Plasma | ALA | 1.44e-04 | 0.005 | 0.975 | -0.005 | 0.019 | 0.794 | 0.001 | 0.781 | 0.173 | 0.110 | -0.001 | 0.005 | 0.889 | 6 | 180 |
| RBC | ALA | 0.005 | 0.005 | 0.357 | 0.005 | 0.015 | 0.761 | -1.07e-05 | 0.997 | 0.990 | 0.985 | 0.014 | 0.011 | 0.197 | 32 | 1076 |
| RBC | LA | -4.20e-04 | 0.003 | 0.874 | -0.009 | 0.006 | 0.149 | 0.002 | 0.122 | 0.543 | 0.626 | -0.005 | 0.004 | 0.219 | 32 | 1076 |
| RBC | GLA | -0.016 | 0.008 | 0.043 | 0.014 | 0.023 | 0.529 | -0.007 | 0.135 | 0.965 | 0.983 | -0.019 | 0.015 | 0.208 | 32 | 1076 |
| RBC | DGLA | -1.03e-04 | 0.004 | 0.977 | 0.004 | 0.008 | 0.640 | -0.001 | 0.588 | 0.648 | 0.614 | -0.003 | 0.006 | 0.569 | 32 | 1076 |
| RBC | AA | 3.42e-04 | 0.002 | 0.840 | 0.003 | 0.004 | 0.393 | -0.001 | 0.387 | 0.605 | 0.595 | -0.002 | 0.003 | 0.406 | 32 | 1076 |
| Plasma | DPA-n3 | 0.005 | 0.013 | 0.726 | 0.019 | 0.056 | 0.757 | -0.002 | 0.810 | 0.275 | 0.183 | -0.011 | 0.015 | 0.482 | 6 | 180 |
| RBC | DPA-n3 | 0.002 | 0.003 | 0.501 | -2.94e-04 | 0.006 | 0.963 | 5.00e-04 | 0.696 | 0.585 | 0.542 | 0.002 | 0.004 | 0.571 | 32 | 1076 |
| RBC | DTA | -2.57e-04 | 0.004 | 0.952 | 0.002 | 0.009 | 0.814 | -0.001 | 0.767 | 0.477 | 0.430 | -0.002 | 0.006 | 0.701 | 32 | 1076 |
| Plasma | DHA | -0.058 | 0.037 | 0.120 | -0.098 | 0.200 | 0.648 | 0.006 | 0.842 | 0.807 | 0.690 | -0.046 | 0.065 | 0.480 | 6 | 180 |

PUFA, polyunsaturated fatty acid; RBC, red blood cell; ALA, α-Linolenic acid; LA: linoleic acid; GLA, γ-Linoleic acid; DGLA, dihomo-γ-linoleic acid; AA, arachidonic acid; DPA-n3, docosapentaenoic acid; DTA, docosatetraenoic acid; DHA, docosahexaenoic acid; β, causal effect size; SE, standard error; IVW_MRE, inverse-variance weighted random-effects model; Egger, MR-Egger; nsnps, number of SNPs retained for this analysis.

**Supplementary Table 28. Reverse Mendelian randomization estimates of associations of genetically predicted COVID-19 severity with polyunsaturated fatty acids based on the release 4 HGI B1 (COVID-19 SNP *P* < 5e-6).**

| | | IVW_MRE | | | MR-Egger | | | Wald ratio | | | MR-Egger intercept | | Heterogeneity test | | Weighted median | | | | |
|---|---|---|---|---|---|---|---|---|---|---|---|---|---|---|---|---|---|---|---|
| **Group name** | **PUFA** | **β** | **SE** | **P** | **β** | **SE** | **P** | **β** | **SE** | **P** | **β** | **P** | **P_IVW** | **P_Egger** | **β** | **SE** | **P** | **nsnps** | **F-statistic** |
| Plasma | ALA | - | - | - | - | - | - | 0.006 | 0.006 | 0.317 | - | - | - | - | - | - | - | 1 | 22 |
| RBC | ALA | 0.002 | 0.009 | 0.834 | -0.008 | 0.021 | 0.696 | - | - | - | 0.004 | 0.588 | 0.471 | 0.408 | 0.004 | 0.013 | 0.778 | 11 | 262 |
| RBC | LA | 0.001 | 0.003 | 0.807 | -0.001 | 0.008 | 0.886 | - | - | - | 0.001 | 0.784 | 0.632 | 0.546 | -0.001 | 0.005 | 0.771 | 11 | 262 |
| RBC | GLA | 0.011 | 0.014 | 0.420 | 0.072 | 0.030 | 0.037 | - | - | - | -0.026 | 0.045 | 0.406 | 0.837 | 0.031 | 0.019 | 0.099 | 11 | 262 |
| RBC | DGLA | 0.004 | 0.006 | 0.492 | 0.001 | 0.013 | 0.960 | - | - | - | 0.001 | 0.778 | 0.180 | 0.133 | 0.001 | 0.006 | 0.842 | 11 | 262 |
| RBC | AA | 0.001 | 0.002 | 0.470 | 0.004 | 0.005 | 0.428 | - | - | - | -0.001 | 0.528 | 0.863 | 0.837 | 0.003 | 0.003 | 0.359 | 11 | 262 |
| Plasma | DPA-n3 | - | - | - | - | - | - | 0.001 | 0.021 | 0.976 | - | - | - | - | - | - | - | 1 | 22 |
| RBC | DPA-n3 | -0.003 | 0.004 | 0.409 | -0.007 | 0.008 | 0.408 | - | - | - | 0.002 | 0.588 | 0.510 | 0.445 | -0.002 | 0.005 | 0.716 | 11 | 262 |
| RBC | DTA | 0.009 | 0.004 | 0.016 | 0.019 | 0.012 | 0.152 | - | - | - | -0.004 | 0.378 | 0.929 | 0.941 | 0.004 | 0.007 | 0.589 | 11 | 262 |
| Plasma | DHA | - | - | - | - | - | - | 0.109 | 0.095 | 0.249 | - | - | - | - | - | - | - | 1 | 22 |

PUFA, polyunsaturated fatty acid; RBC, red blood cell; ALA, α-Linolenic acid; LA: linoleic acid; GLA, γ-Linoleic acid; DGLA, dihomo-γ-linoleic acid; AA, arachidonic acid; DPA-n3, docosapentaenoic acid; DTA, docosatetraenoic acid; DHA, docosahexaenoic acid; β, causal effect size; SE, standard error; IVW_MRE, inverse-variance weighted random-effects model; Egger, MR-Egger; nsnps, number of SNPs retained for this analysis.

**Supplementary Table 29. Reverse Mendelian randomization estimates of associations of genetically predicted COVID-19 susceptibility with polyunsaturated fatty acids based on the release 4 HGI C2 (COVID-19 SNP *P* < 5e-6).**

| | | IVW_MRE | | | MR-Egger | | | MR-Egger intercept | | Heterogeneity test | | Weighted median | | | | |
|---|---|---|---|---|---|---|---|---|---|---|---|---|---|---|---|---|
| **Group name** | **PUFA** | **β** | **SE** | ***P*** | **β** | **SE** | ***P*** | **β** | ***P*** | ***P*_IVW** | ***P*_Egger** | **β** | **SE** | ***P*** | **nsnps** | ***F*-statistic** |
| Plasma | ALA | -0.001 | 0.006 | 0.891 | 0.021 | 0.053 | 0.705 | -0.002 | 0.693 | 0.349 | 0.262 | -0.002 | 0.007 | 0.756 | 7 | 172 |
| RBC | ALA | 0.013 | 0.011 | 0.206 | -0.004 | 0.037 | 0.909 | 0.002 | 0.607 | 0.982 | 0.977 | 0.018 | 0.022 | 0.412 | 19 | 556 |
| RBC | LA | 0.001 | 0.006 | 0.881 | -0.014 | 0.014 | 0.328 | 0.002 | 0.252 | 0.476 | 0.504 | -1.91e-04 | 0.009 | 0.983 | 19 | 556 |
| RBC | GLA | -0.030 | 0.015 | 0.043 | -0.015 | 0.055 | 0.781 | -0.002 | 0.776 | 0.990 | 0.985 | -0.015 | 0.032 | 0.638 | 19 | 556 |
| RBC | DGLA | -0.013 | 0.010 | 0.171 | -0.020 | 0.023 | 0.401 | 0.001 | 0.754 | 0.166 | 0.133 | -0.008 | 0.013 | 0.543 | 19 | 556 |
| RBC | AA | -0.003 | 0.005 | 0.518 | 0.001 | 0.011 | 0.894 | -0.001 | 0.658 | 0.183 | 0.152 | -0.002 | 0.006 | 0.733 | 19 | 556 |
| Plasma | DPA-n3 | 0.006 | 0.018 | 0.732 | -0.178 | 0.151 | 0.293 | 0.014 | 0.275 | 0.341 | 0.390 | 0.019 | 0.023 | 0.417 | 7 | 172 |
| RBC | DPA-n3 | 0.008 | 0.007 | 0.286 | 0.005 | 0.017 | 0.759 | 2.77e-04 | 0.883 | 0.212 | 0.168 | 0.005 | 0.010 | 0.587 | 19 | 556 |
| RBC | DTA | -0.003 | 0.012 | 0.818 | -0.019 | 0.029 | 0.515 | 0.002 | 0.538 | 0.050 | 0.043 | -0.004 | 0.014 | 0.769 | 19 | 556 |
| Plasma | DHA | -0.137 | 0.082 | 0.095 | -0.840 | 0.710 | 0.290 | 0.055 | 0.364 | 0.467 | 0.463 | -0.120 | 0.116 | 0.301 | 7 | 172 |

PUFA, polyunsaturated fatty acid; RBC, red blood cell; ALA, α-Linolenic acid; LA: linoleic acid; GLA, γ-Linoleic acid; DGLA, dihomo-γ-linoleic acid; AA, arachidonic acid; DPA-n3, docosapentaenoic acid; DTA, docosatetraenoic acid; DHA, docosahexaenoic acid; β, causal effect size; SE, standard error; IVW_MRE, inverse-variance weighted random-effects model; Egger, MR-Egger; nsnps, number of SNPs retained for this analysis.
